# Mapping the comorbid landscape of Parkinson’s disease and Crohn’s disease along the gut-blood-brain axis

**DOI:** 10.1101/2025.10.01.25337087

**Authors:** Yanshi Hu, Bentao She, Zhounan Yin, Xinjian Yu, Wenyi Wu, Ming Chen

## Abstract

Parkinson’s disease (PD) and Crohn’s disease (CD) are primarily localized to the brain and gut, respectively. Nevertheless, epidemiological evidence increasingly links these two seemingly unrelated disorders. Although genomic or transcriptomic efforts have been dedicated to understanding this phenomenon, the precise landscape underlying this comorbidity remains elusive. Here, a systematic multi-omics approach is employed to panoramically map this pathogenic nexus for the first time. By curating a comprehensive genetic corpus related to PD and CD from extensive publications, we uncovered a shared genetic architecture converging on biological functions governing host-pathogen interactions and barrier integrity maintenance. Further, multi-tissue transcriptomic datasets were meta-analyzed to validate genomic insights in transcriptional circumstances, which identified pervasive transcriptional synergies of PD and CD pathways within the blood context, indicating in blood CD pathological milieu could create a permissive environment for PD pathogenesis. Finally, delineating the aberrant gut-blood-brain axis through the sequential compromise of gut epithelial barrier, gut-vascular barrier and blood-brain barrier, we revealed a directional cascade where CD intestinal pathology facilitates PD substantia nigra degeneration via blood circulation, establishing a theoretical foundation for preventive and therapeutic interventions for PD and CD comorbidity. Crucially, this study provides a blueprint for dissecting the molecular etiology of comorbidities in other complex diseases affecting disparate anatomical sites.

## 1. Introduction

As one of the devastating neurodegenerative diseases, Parkinson’s disease (PD) is characterized by both progressive motor and non-motor symptoms which affect daily life, and is reported by The Global Burden of Disease study to have the fastest increase in global prevalence and mortality among neurological diseases [1]. Though how PD occurs and evolves remains elusive, mounting evidence indicates vital roles of genetics in its sporadic form [2,3]. Crohn’s disease (CD), a chronic and recurrent type of inflammatory bowel disease (IBD), often occurs in the ileocolic and colonic parts of the intestine [4,5]. For the United States only, there are more than 700,000 CD patients [6]. It is widely accepted that CD pathogenesis is comprised of genetic, microbiomic and other environmental elements [4,7]. Growing knowledge has led to the hypothesis that chronic intestinal inflammation or IBD might elicit PD [8,9]. While controversies on the impact of CD on PD exist [10-12], more and more epidemiological [13-17], genomic [18-21], transcriptomic [22,23], and biochemical studies [24] have strengthened the connections between these two diseases.

Numerous investigations have endeavored to dissect the shared molecular mechanisms driving PD and CD comorbidity through diverse perspectives. Kang *et al*. [20] unraveled the intricate genetic interplay between PD and IBD by identifying novel pleiotropic loci that exhibit a mixture of synergistic and antagonistic effects, thereby highlighting the pivotal role of immune-mediated mechanisms and post-translational modifications in their shared etiology. By leveraging whole-genome data from cohorts with comorbid IBD and PD, Kars *et al*. [21] characterizes the landscape of shared high-impact rare variants, confirming *LRRK2* pleiotropy while identifying novel candidate genes involved in inflammation and autophagy, such as *IL10RA*, through network-based heterogeneity clustering and phenome-wide association studies. However, these two studies are restricted to the genomic level, which lacks subsequent functional validation (e.g., transcriptional activity) of these genetic findings. Zheng *et al*. [22] and Sun *et al*. [23] separately sought to uncover pathogenic pathways enriched in common differentially expressed genes (DEGs) from transcriptomes in the same tissue, i.e., peripheral blood of PD and CD patients, and from those in the different tissues, i.e., PD substantia nigra and IBD colonic mucosa. Reliance on single transcriptomic dataset is frequently plagued by small-sample bias, potentially compromising the robustness and reproducibility of study conclusions [25]. Furthermore, juxtaposing transcriptomic alterations across spatially distinct tissues—such as the substantia nigra in PD versus the colon in CD—offers limited insight into the intrinsic etiology of this comorbidity. Given that both pathologies possess substantial heritable components [2,7], exclusive reliance on transcriptomic profiling may prove insufficient to pinpoint the definitive drivers governing their co-occurrence. Given systemic insights into the pairwise relation between PD and CD are still lacking [26], proposing a multi-omics framework, which synergizes genomics and transcriptomics in a systems biology architecture to identify bona fide pathogenic drivers for PD-CD comorbidity in a holistic resolution, is imperative.

In the current study, a systematic approach is employed to panoramically decipher the molecular mechanisms underpinning the comorbidity of PD and CD for the first time. We initiated our investigation by compiling a comprehensive catalog of genetic variants and genes relevant to PD and CD from extensive literature and databases, unveiling a shared genetic architecture that converges on biological functions governing host-pathogen interactions and barrier integrity maintenance. To ensure robustness and refine these genomic insights in transcriptional contexts, transcriptomic datasets in various tissues were integrated through meta-analysis technique, which identified critical driver factors and uncovered profound transcriptional synergies of PD and CD within the blood compartment. Notably, this analysis revealed a suggestive transcriptional resemblance between the CD blood milieu and the substantia nigra pathology of PD. We further substantiated a physical basis for this connection by delineating a sequential breach of the gut epithelial barrier, gut-vascular barrier and blood-brain barrier. These findings culminate in a cohesive gut-blood-brain axis model, positing a directional pathogenic cascade where intestinal pathology in CD promotes PD neurodegeneration via blood circulation, thereby establishing a theoretical foundation for future preventive and therapeutic interventions for PD and CD comorbidity.

## 2. Materials and Methods

### 2.1. Genetic association data curation

We utilized a tailored pipeline (Figure 1a) to retrieve, aggregate and normalize genetic association data for PD and CD. Specifically, in the light of the unbiased nature of genome-wide association study (GWAS) to unearth genetic variants across whole genome [27], PheGenI [28] was leveraged to obtain GWAS data of PD and CD. Only variants which surpass genome-wide significant threshold were kept. Considering complex disorder phenotypes, such as PD and CD, are typically featured by synergistic variants with very small or mild individual effects [29], genetic factors not satisfying stringent cutoff but with suggestive or nominal associations might virtually matter. So, candidate gene-based genetic association studies as complements were also taken into consideration. This kind of studies has two-(or three-) fold evidence: pre-study biological hints, within-study conclusions (and post-study validations).

**Figure 1.**
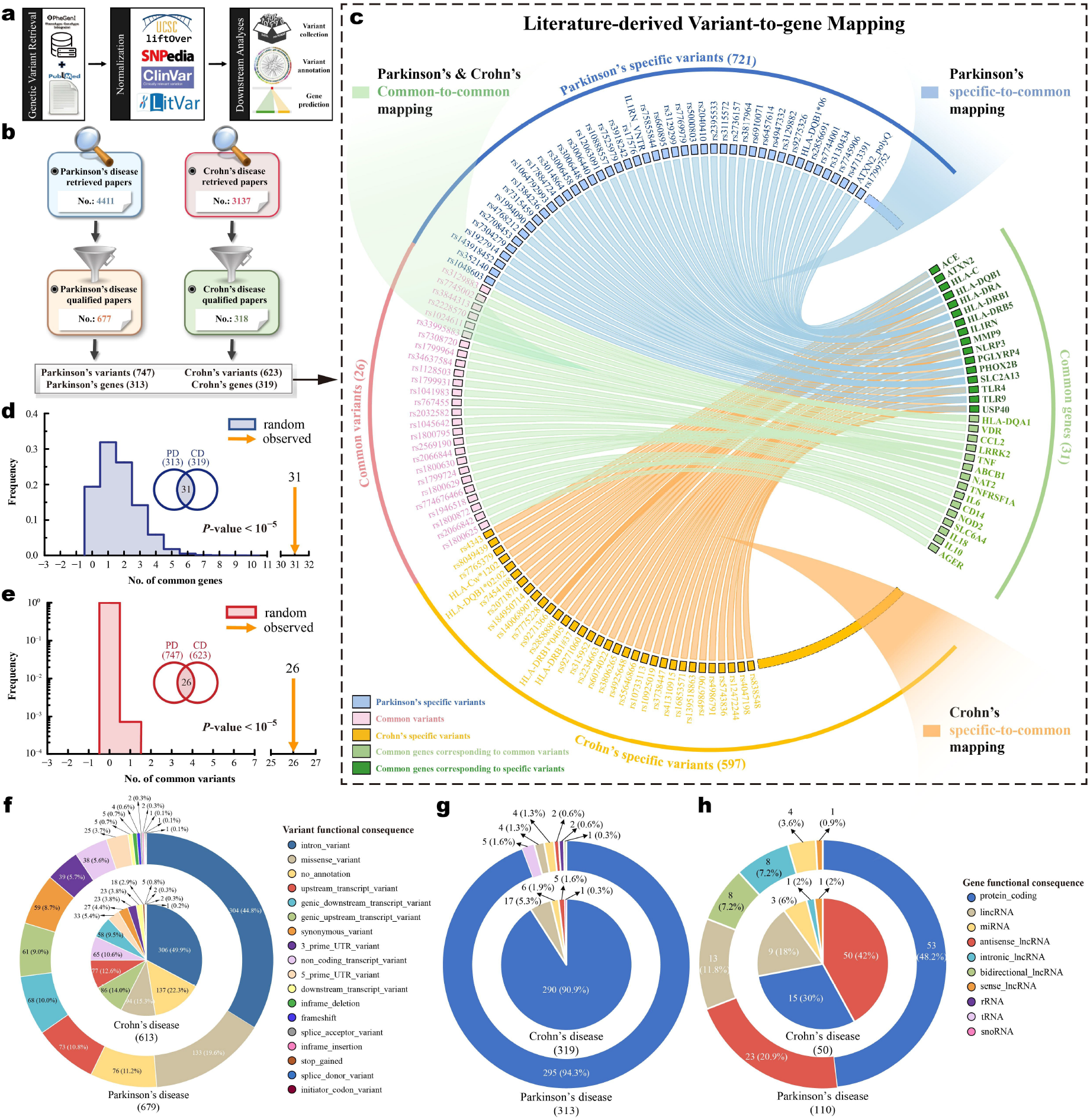
Variant-centric dissection reveals a shared genetic architecture of PD and CD. **(a)** Retrieval, integration, and standardization process of PD and CD genetic variants. **(b)** Statistics of initial PubMed retrieval, filtration, and final genetic variants and genes relevant to PD and CD. **(c)** Literature-derived variant-to-gene mapping showcases the gene intersection between PD and CD can be derived from common variants (Parkinson’s & Crohn’s common-to-common mapping) or disease-specific variants (Parkinson’s or Crohn’s specific-to-common mapping). **(d, e)** The gene and variant intersections between PD and CD are statistically significant, with empirical P-value determined by 100,000 Monte Carlo experiments. **(f)** Functional consequences of PD and CD genetic variants. **(g)** Functional consequences of PD and CD genetic association genes. (h) Functional consequences of PD and CD dbSNP-predicted genes.

Due to the fact that MeSH term labelling is time-consuming and lagging [30,31], PubMed search using MeSH terms would undoubtedly omit quite a lot of related literature. Thus, in order to fetch as many relevant publications indexed in PubMed as possible, we revised our previous literature retrieval terms [32] as “Parkinson* AND (Polymorphism OR Genotype OR Alleles) NOT Neoplasms” and “Crohn* AND (Polymorphism OR Genotype OR Alleles) NOT Neoplasms” for PD and CD, respectively. All abstracts were scanned and only genetic association studies reporting significant associations with PD and CD from retrieved publication corpus were included for in-detailed selection. Further, full contents of all qualified papers were reviewed to ensure the consistency of those drawn conclusions. To converge irregular variants into a uniform corpus, several tools were adopted for final variant standardization. UCSC liftOver [33] was utilized to calibrate PD and CD-related variants from various genome coordinate versions to GRCh38 assembly. SNPedia [34], LitVar [35] and ClinVar [36] were used to normalize polymorphic sites into standard formats.

Monte Carlo experiment was implemented to testify if variant- and gene-level commonalities between PD and CD are biologically meaningful. Specifically, let *G* designate whole human variants (667,501,404 live Ref SNPs in NCBI dbSNP database) / genes (61,197 in NCBI Entrez Gene database). Let *P* and *C* denote the variant/gene sizes related to PD and CD separately. We randomly chose variants with the same number as PD and CD ones from *G* 100,000 timesdistinctively, and the empirical *P*-value was calculated as the number of scenarios with no fewer than the observed *N* shared variants / genes divided by 100,000. The final *P*-value is given by the following formula:
154 cl:1036

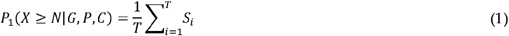

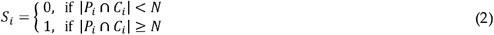

where *T* is constant 100,000, *S*_i_ is a Boolean variable indicating the status in the *i*-th experiment, in which *P*_i_ and *C*_i_ are variant / gene sets of PD and CD correspondingly.

### 2.2. Bio-functional aggregation of genetic association data

Capitalizing on latest Gene Ontology (GO) and Kyoto Encyclopedia of Genes and Genomes (KEGG), clusterProfiler [37] was leveraged to detect biological meanings hidden behind PD and CD genetic association genes with over-representation analysis (ORA) method. GO provides the world’s largest information multiplicity source on gene function, saturated with Biological Process (BP), Cellular Component (CC) and Molecular Function (MF) facets. Described as a directed acyclic graph, GO possesses loose hierarchies in which child (leaf) items are more specialized and concrete than corresponding parent (root) terms. With the notion that leaf GO terms are more informative than root terms, a customized strategy was used in the current study: leaf enriched GO terms were picked as agents for biology-level interpretations. All overrepresented GO / pathway terms were further refined by multiple testing correction with false discovery rate (FDR) value 0.05 as significance threshold.

### 2.3. Human biological pathway network construction

Distinctive biological pathways act in a concerted manner to organize micro- and macro-phenotypes. However, current biological pathways are largely discrete and need to be weaved together. Our previous work attempted solving this problem using network separation distance metric [38]. In present study, this framework was implemented in the context of a high-quality human protein interactome [39]. Albeit lots of pathway resources have been available, such as KEGG and reactome [40], pathway definitions and criteria of these databases differ from each other, and merging or comparing various pathway resources is a pretty arduous task. As a widely used pathway source, only KEGG pathway system was used to construct human biological pathway network. KEGG pathways belonging to the category “Human Diseases” were filtered out for further analysis, because these specific ones are meta-pathways, i.e., merged version of many involved pathways, not reflecting the original physiological profiles inside human cells. Members of a pathway tend to converge into one or more module(s) in the context of human interactome. Pathways with significant network modularity were deemed real biological pathways and retained. As indicated by our previous work [38], network distance between two pathways should be negative if this pathway pair has connections, thus pathways with network separation distance less than zero were kept.

### 2.4. Transcriptomic meta-analysis of PD and CD

Transcriptome is able to figure out how the genetic factors virtually function. PD and CD transcriptomic data were fetched from GEO, SRA, and ArrayExpress. As the most affected sites, substantia nigra transcriptome data were used for PD, and colonic and ileal mucosa ones for CD. Owing to currently no public transcriptomic dataset(s) of individual patients with both PD and CD and the fact that blood transcriptomic profiling has proved to be unbiased and shown efficacy in idiopathic PD and CD diagnosis [41-44], as a compromise, peripheral blood datasets in either PD or CD were used for preliminary dissection of etiological linkages between these two disorders. The detailed information of transcriptomic data collection and processing used in this study can be found in our recent work [45], where a novel transcriptomic meta-analysis method, AWmeta, was proposed to aggregate multiple PD or CD transcriptomic data in these five tissues for robust DEGs with reliable fold change (FC) and corrected *P*-value, respectively.

### 2.5. Transcriptomic GO and pathway enrichment of PD and CD

To elucidate the functional manifestation of the genetic architecture in relevant tissues, we performed a multi-level transcriptomic analysis. First, we identified DEGs from the transcriptomic meta-analyses across five disease-relevant contexts: the substantia nigra and blood in PD, and the ileal and colonic mucosa and blood in CD (see “*Transcriptomic meta-analysis of PD and CD*” section for details). A stringent statistical threshold of |log_2_FC| > log_2_1.5 and *P* < 0.05 was applied to define the DEG sets. Subsequently, to comprehensively map the functional landscape, we conducted enrichment analyses for both GO terms and biological pathways using two complementary algorithms: ORA on the DEG sets and gene set enrichment analysis (GSEA) on the complete ranked gene lists [46]. To leverage the unique strengths of both methods, we adopted a consolidated approach, assigning the minimum *P*-value obtained from either ORA or GSEA as the definitive significance score for each item. Finally, to quantify the functional activity of key biological processes identified at the genetic level, we focused on the previously identified GO hotspots and the genetically-implicated biological pathways. We utilized normalized enrichment score (NES) derived from GSEA as a robust metric to represent the activation or suppression status of these specific pathways and GO terms within each of the five disease-tissue profiles.

### 2.6. Transcriptomic genes and pathway correlation calculation of PD and CD using signed maximal information coefficient

To interrogate the associative relationships between transcriptional signatures in PD and CD at both the gene and pathway levels, we sought a metric capable of transcending the limitations of conventional linear correlation measures. Recognizing that biological systems are governed by complex and often non-linear dynamics, we employed a robust approach based on the maximal information coefficient (MIC), a novel measure of dependence that excels at detecting a wide spectrum of both linear and non-linear associations with high statistical power [47]. However, as an unsigned metric, MIC quantifies the strength but not the directionality of an association. To preserve this critical information, we implemented a signed version, the signed maximal information coefficient (SMIC), by integrating the MIC value with the directionality provided by the Spearman’s rank correlation coefficient (*ρ*). The SMIC between two variables, X and Y, is formally defined as:

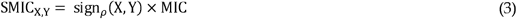

where sign_*ρ*_ (X,Y) denotes the sign (+1 or −1) of the Spearman correlation. MIC values were computed using the minerva R package [48]. This hybrid approach allows us to capture the full complexity of transcriptional relationships while retaining an intuitive understanding of the crucial directional context.

2.7. Transcriptomic pathway synergy measure of PD and CD based on acting-in-concert score

Recognizing that simple correlation metrics are often insufficient to capture the intricate interplay between complex diseases, we moved beyond simple pairwise associations to quantify functional cooperativity at the pathway level. The overall activity of a shared biological pathway can be misleading, as it may obscure critical synergistic or antagonistic behaviors among its constituent members. To dissect these sub-pathway dynamics, we conceptualized and implemented a novel metric, the acting-in-concert score (ACS), designed to quantify the degree of concordant regulation within a given pathway across two disease states. A gene is defined as “acting in concert” if it is directionally consistent in its dysregulation, i.e., up-or down-regulated in both diseases. The ACS for a given pathway is calculated as:

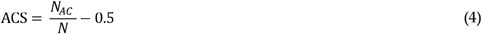

where is the number of constituent genes acting in concert, and is the total number of genes in the pathway. A positive ACS signifies a net synergistic propensity within the pathway, suggesting that the pathological states of the two diseases reinforce each other, while a negative ACS indicates a predominantly antagonistic relationship.

### 2.8. Intestinal and brain barrier permeability biomarker collection

To evaluate the integrity of key biological interfaces implicated in the gut-brain axis, we curated comprehensive panels of molecular biomarkers based on an extensive literature review. Recognizing that compromised barrier function is a pivotal event in initiating and perpetuating pathological inflammatory and neurodegenerative cascades, we focused on three critical barriers. First, for gut epithelial barrier (GEB), which delineates the intestinal lumen from the lamina propria and serves as the primary sentinel against the translocation of luminal pathogens and antigens [49], we compiled a list of established permeability markers (Table S1). Subsequently, we collated a set of indicators reflective of the integrity of gut-vascular barrier (GVB) (Table S2), a crucial checkpoint that restricts the passage of microbial derivatives from the intestinal interstitium into systemic circulation, thereby preventing systemic dissemination and end-organ damage [50]. Finally, to probe the status of the central nervous system interface, we assembled a panel of well-characterized biomarkers for blood-brain barrier (BBB) (Table S3), a highly selective physiological boundary that meticulously regulates molecular traffic between the periphery and the brain parenchyma to safeguard neural homeostasis against blood-borne insults [51].

## 3. Results

### 3.1. Variant-centric dissection reveals a shared genetic architecture of PD and CD

Following a stringent processing strategy (Figure 1a and “Genetic association data curation” in Materials and Methods), our keyword-based search on PubMed initially yielded 4,411 and 3,137 publications for PD and CD, respectively. A meticulous literature filtration process subsequently narrowed this corpus to 677 and 318 publications with genuine relevance to genetic variants in PD and CD. Finally, a comprehensive full-text review allowed us to curate a high-confidence dataset comprising 747 genetic variants in 313 genes for PD, and 623 variants in 319 genes for CD (Figure 1b; Table S4 and S5).

A comparative analysis of these datasets revealed a significant genetic intersection, with 26 variants and 31 genes shared between PD and CD (Figure 1c). Intriguingly, 16 of these 31 shared genes were exclusively linked to disease-specific variants. This finding points toward a compelling comorbid mechanism wherein distinct genetic perturbations converge upon a common set of genes, thereby orchestrating shared downstream pathological events. Among these shared genes, *MMP9* [52] and *ABCB1* [53] are known to modulate blood-brain barrier permeability, providing a potential conduit for CD peripheral pathologies to influence central nervous system processes in PD. The presence of *PGLYRP4*, a peptidoglycan recognition protein [54], hints at a shared role for bacterial sensing and immune response in both disorders. *ACE* links the shared genetic risk to dysregulated blood pressure, a non-motor feature associated with PD pathology [55]. The inflammasome component *NLRP3* [56,57], along with downstream NF-κB signaling cascade [58,59], points to a shared inflammatory axis, which is pathologically active in PD and CD. Mutations of *LRRK2*—a multi-domain protein—confer significant risk for both conditions [60,61]. Collectively, these findings establish a substantive genetic nexus between PD and CD.

To ascertain that the observed genetic overlap was statistically significant and not a product of random chance, we employed a Monte Carlo simulation approach with 100,000 iterations to calculate empirical *P*-values. The simulation robustly confirmed the profound statistical significance of the overlap for both the genetic variants and their associated genes (*P* < 10^−5^ for both), underscoring the authenticity of this shared genetic architecture (Figure 1d, e).

We further delved into the genomic distribution and functional annotation of the variant datasets, which revealed strikingly similar patterns between the two diseases (Figure S1a, b). Variants for both were distributed across the autosomes. However, several key distinctions emerged. Notably, CD variants were identified on the Y chromosome while PD variants were not, an observation that aligns perfectly with recent reports negating a strong link between PD and the Y chromosome [62]. Conversely, mitochondrial variants were unique to PD (Figure S1c), suggesting that mitochondrial dysfunction, while a hallmark of PD, may not be a primary genetic driver in CD. The pathogenic contribution of mitochondrial variants in PD does not appear to be dominated by any single functional class. Instead, synonymous, missense and unannotated variants are present in commensurate frequencies and uniformly scattered across the mitochondrial genome, raising the possibility that all three types contribute significantly to the disease genetic risk. A comparison of variant functional annotations further solidified this disease similarity, with variants predominantly located in intronic regions, followed by missense and unannotated variants (Figure 1f). This highlights that the functional consequences of many variants implicated in PD and CD comorbidity remain to be elucidated.

Expanding our analysis to the gene level, we observed a continued trend of functional similarity. In both diseases, over 90% of the genetically-implicated genes were protein-coding (Figure 1g). This starkly contrasts with the vastness of the non-coding genome, highlighting a significant knowledge gap and potential bias in current genetic studies. To address this deficit, we leveraged variant annotations from the dbSNP database to predict novel disease-related genes, identifying 110 and 50 potential candidates for PD and CD, respectively (Table S6 and S7). The credibility of these predictions was substantiated through both extensive literature mining and quantitative evidence from AWmeta [45]. An examination of these expanded gene sets revealed a dramatic shift: non-coding genes now constitute over 50% of the total in both diseases (Figure 1h). This effort effectively supplements the initial protein-coding-centric view, providing a more comprehensive and balanced genetic landscape for exploring the comorbidity between PD and CD.

### 3.2. Variant-enriched disease pathways implicate PD and CD comorbidity

Given that KEGG provides a well-curated repository of biological pathways linked to human diseases [63], it serves as a powerful platform for investigating inter-disease connections at a functional enrichment level. We therefore performed a disease pathway enrichment analysis on the PD and CD genetic variants (see “*Bio-functional aggregation of genetic association data*” in Materials and Methods). Our objective was to map the broader disease landscape associated with each disorder, thereby gaining novel perspectives on the mechanistic underpinnings of their comorbidity.

This enrichment analysis revealed that PD and CD share 38 human disease pathways (Figure S2; Table S8 and S9). Notably, these commonly enriched pathways consistently demonstrated greater statistical significance than the disease-specific pathways for either condition, suggesting they represent core pathological processes of central importance to both PD and CD. As a critical internal validation of our gene sets, the top-enriched pathway for PD was “Parkinson disease” itself, with the three most significant hits all being neurodegenerative disorders; similarly, the premier hit for CD was “Inflammatory bowel disease”.

A striking asymmetry emerged from this analysis. While “Inflammatory bowel disease” ranked as the eighth most significant pathway for the PD genes, the reciprocal was not true: “Parkinson disease”, or indeed any neuro-related pathway, was conspicuously absent from the CD enrichment results. This non-reciprocal relationship suggests a directional influence, where the genetic architecture of PD encompasses susceptibility to Inflammatory bowel disease-like processes, whereas the genetic basis of CD does not inherently predispose to parkinsonism. This finding lends a genetic support to the clinical hypothesis that inflammatory processes originating in conditions like CD can act as a catalyst for PD initiation or progression, while the reverse is not established [64,65].

Further exploration of the shared pathways provided deeper mechanistic insights. The enrichment of “Lipid and atherosclerosis” and “Fluid shear stress and atherosclerosis” implicates dysregulated lipid metabolism as a common pathological feature [66,67]. Similarly, the shared enrichment of “Type I diabetes mellitus” and “Insulin resistance” points toward aberrant insulin signaling as a convergent metabolic vulnerability in both diseases [68,69].

Intriguingly, an examination of the disease-specific pathways unveiled a potential etiological link. Pathways for “Hepatitis B” and “Hepatitis C” were uniquely enriched in the CD gene set; concurrently, “Hepatocellular carcinoma” was a specific enrichment item for PD. Given that chronic viral hepatitis is a primary driver of hepatocellular carcinoma [70], this constellation of findings hints at a possible pathogenic trajectory where systemic inflammation and hepatic stress associated with CD genetic background may contribute to a cellular environment conducive to pathologies seen in PD.

### 3.3. PD and CD genetic variants converge on biological functions involving host-pathogen interactions and barrier integrity maintenance

To obtain a panoramic view of the biological functions encoded by the shared genetic architecture of PD and CD, we first conducted a broad-stroke GO enrichment analysis using WebGestalt [71]. This revealed a remarkably congruent functional landscape for both diseases at a high level (Figure 2a). Within GO-BP category, terms such as “biological regulation”, “response to stimulus”, and “metabolic process” were top hits for both, implicating shared roles for regulatory homeostasis, environmental stress responses, and metabolism. Similarly, in GO-MF and GO-CC domains, terms like “protein binding”, “ion binding”, and “membrane” were commonly enriched, highlighting the importance of molecular interactions at cellular membranes in the etiology of both disorders.

**Figure 2.**
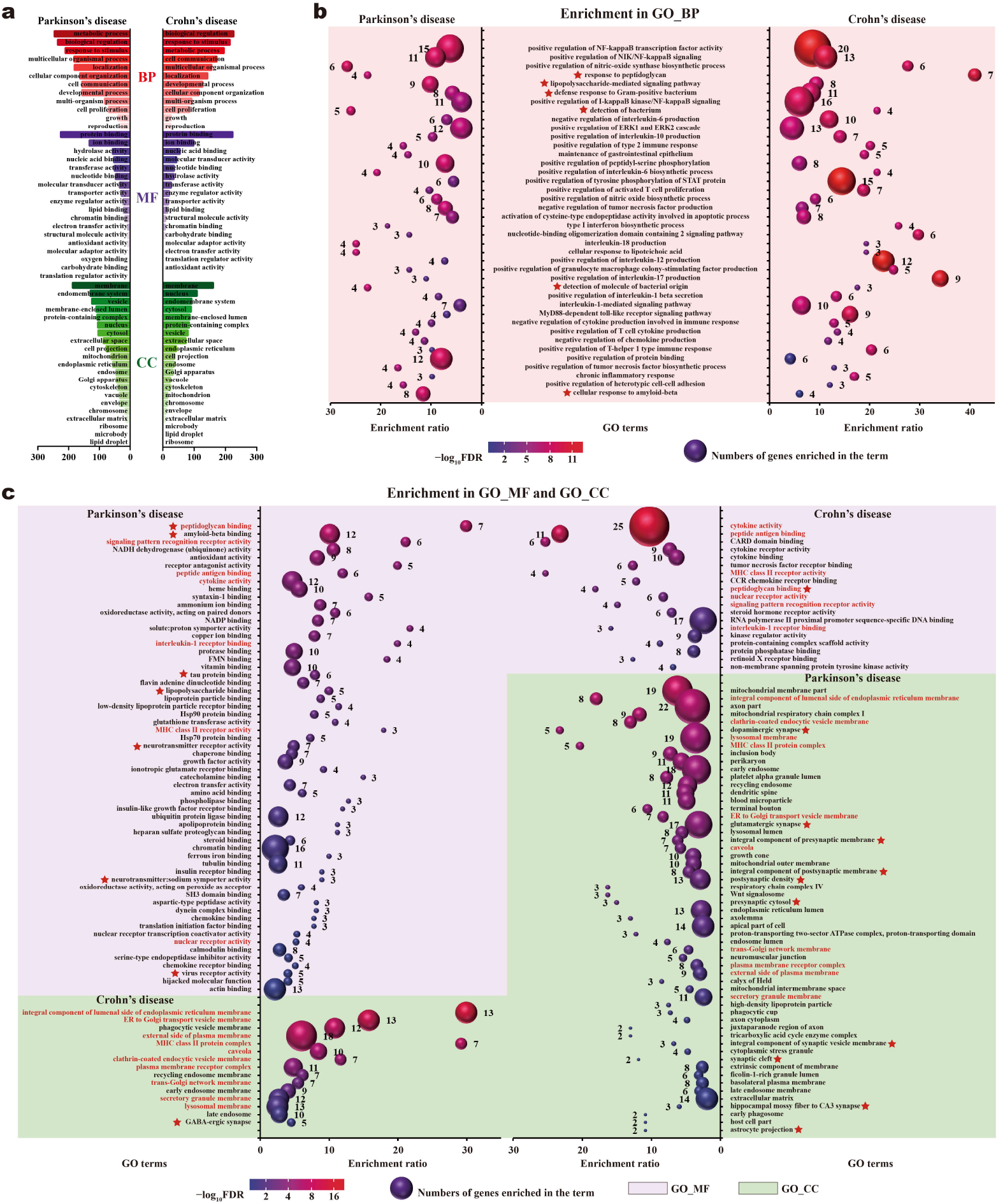
PD and CD genetic variants converge on biological functions involving pathogen- and derivative-induced infection, recognition and resistance and barrier integrity maintenance. **(a)** Broad GO terms enriched by genetic association genes in PD and CD. The x-axis represents the number of genes included in the enriched GO terms (y-axis). Red, purple, and green indicate GO-BP, GO-MF, and GO-CC, respectively. **(b)** The same significantly enriched GO-BP terms (top 40) for genetic association genes in both PD and CD. **(c)** Significantly enriched GO-MF and GO-CC terms for genetic association genes in PD and CD. The x-axis represents the enrichment ratio of the enriched GO terms (y-axis). The sphere size indicates the number of genes enriched in each GO term, and the sphere color the enrichment significance. GO term name in red denotes the same GO terms enriched by genetic association genes in both PD and CD within the GO-MF and GO-CC categories. GO terms marked with red asterisks are GO hotspots, whose transcriptional activities in multiple disease tissues were quantified in Figure 3.

To achieve greater functional resolution, we performed a more detailed GO enrichment analysis. Examination of the top 40 shared significantly-enriched GO-BP terms unveiled a striking thematic convergence on pathogen recognition and host defense (Figure 2b; Table S10). A substantial proportion of these terms, including “response to peptidoglycan”, “lipopolysaccharide-mediated signaling pathway”, and “detection of molecule of bacterial origin”, strongly implicated host-pathogen interactions as a central element of the shared genetic risk. Two particularly compelling findings emerged from this analysis. First, the enrichment of “positive regulation of nitric-oxide biosynthetic process” was surprising, as nitric oxide is a potent modulator of blood-brain barrier integrity [72]. This finding provides a direct genetic link to potential blood-brain barrier dysfunction as a component of the comorbidity. Second, the enrichment of “maintenance of gastrointestinal epithelium” pointed directly to the intestinal epithelium’s homeostasis as a shared biological vulnerability. The GO-MF and GO-CC results further substantiated the pathogen response theme (Figure 2c; Table S11–S14), solidifying the hypothesis that processes governing the recognition of and resistance to pathogens and their derivatives play a pivotal role in the shared PD and CD mechanism. We designated these potentially functional terms as “GO hotspots” (indicated by red asterisks in Figure 2).

To further determine if these genetically-defined GO hotspots were functionally active in disease-relevant tissues, we analyzed their transcriptional status across the five tissue datasets, i.e., the substantia nigra and blood in PD, and the ileal and colonic mucosa and blood in CD (Figure 3). Remarkably, the pathogen recognition and resistance processes were significantly and broadly activated (NES > 0) across all five contexts. This system-wide activation signature strongly suggests a persistent state of heightened immune surveillance, likely driven by an elevated presence of pathogens or their molecular patterns in these tissues compared to healthy controls.

**Figure 3.**
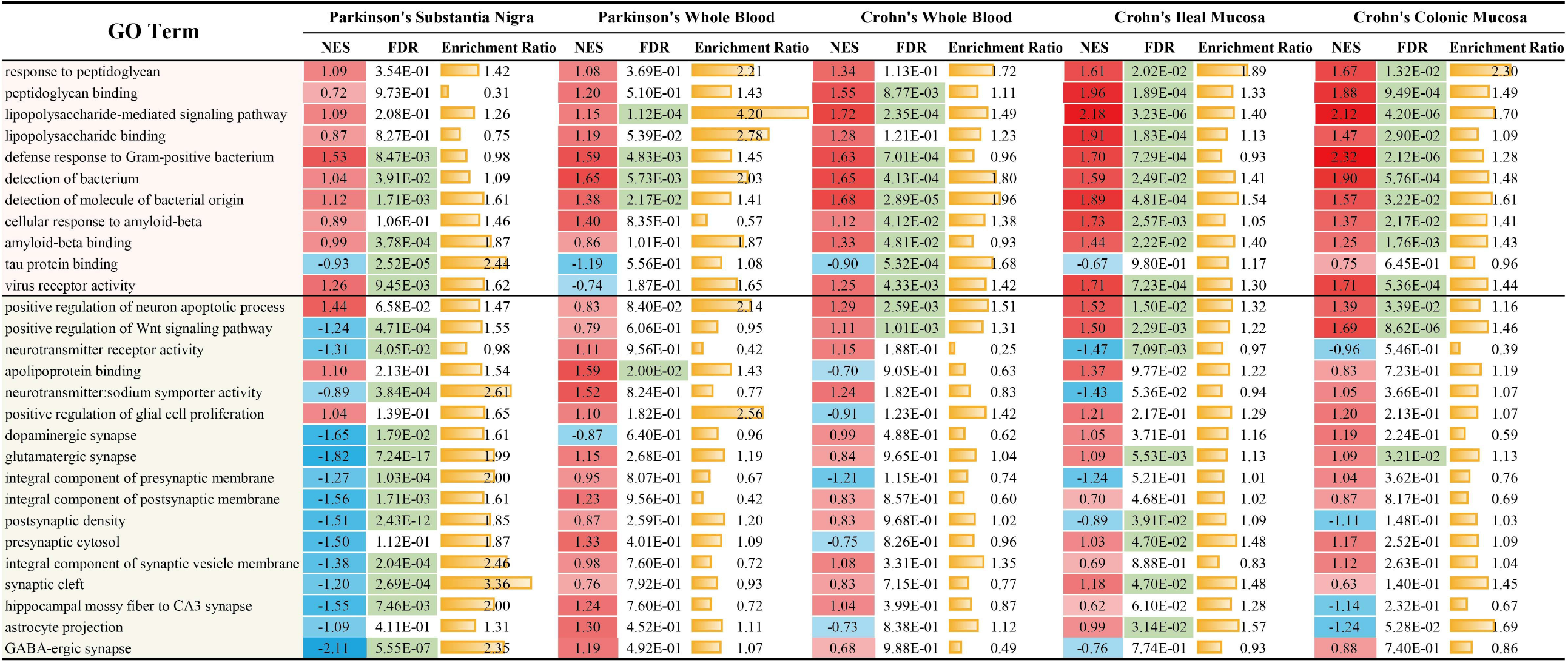
Tissue-wise transcriptional activities of GO hotspots in PD and CD. Pink shading indicates GO hotspots related to the recognition and resistance of pathogens and their relevant derivatives, and gray shading those related to neural activity. NES is used to quantify transcriptional activities of GO hotspots in PD and CD, with red shading representing activation and blue shading repression. FDR with green shading indicates corresponding activity quantification is statistically significant.

As expected, processes related to neuronal activity were globally downregulated (NES < 0) in PD substantia nigra, reflecting its neurodegenerative pathology. These same processes showed no consistent dysregulation in the other four tissues, highlighting their tissue-specific nature. Notably, “positive regulation of neuron apoptotic process” was significantly upregulated across all CD tissues, revealing a previously underappreciated pro-apoptotic process in CD that could impact neuronal health. Furthermore, we observed a significant activation of the “positive regulation of Wnt signaling pathway” in both the ileal and colonic mucosa of CD patients. Given that aberrant Wnt activation is a recognized hallmark of compromised intestinal barrier integrity and increased permeability [73,74], this result provides direct transcriptomic evidence of a defective gut barrier in CD, lending mechanistic support to the gut-origin hypothesis of the comorbidity.

Finally, to assess the relative importance of different biological processes at the genetic level, we compared the top 40 enriched GO-BP terms for each disease (Figure S3; Table S15 and S16). This revealed that the shared functional space was almost exclusively dominated by immune-related processes, cementing the immune system’s role as the central arena for the PD-CD interaction. Moreover, for CD, nearly all of its top 40 enriched terms were immune-related, confirming that immune dysregulation is the preeminent feature of its genetic etiology.

### 3.4. Genetically-informed genes and pathways exhibit coordinated transcriptional activities within and across tissues in PD and CD

To translate the genetic findings into functional contexts, we next determined the biological pathways enriched within the PD and CD variants. This investigation uncovered 18 shared pathways whose enrichment consistently reached a higher statistical significance than pathways specific to either disease alone, underscoring their central importance to the shared pathophysiology (Figure 4a; Table S17 and S18). Among these, the co-enrichments of the “IL-17 signaling pathway” and “Th17 cell differentiation” resonate with our GO findings, as hyperactive Th17 cells are potent producers of the cytokine IL-17, a key mediator of antimicrobial peptide production [75], thus reinforcing the theme of host-pathogen interactions. Intriguingly, the “Intestinal immune network for IgA production” was also a shared pathway, implying that immune processes within the gut may be a common feature and thus pointing to a potential nexus between the two diseases. The “Hematopoietic cell lineage” pathway emerged as one of the most statistically significant shared pathways. This observation aligns with clinical evidence where hematopoietic stem cell transplantation has proven effective in ameliorating symptoms of both PD and CD [76,77], highlighting the critical role of hematopoietic differentiation in their common mechanisms. Furthermore, the presence of the “TNF signaling pathway” is particularly salient, given that anti-TNF therapies for CD are associated with a substantially reduced risk of developing PD [78]. The enrichment of the “Adipocytokine signaling pathway” lends further support to the role of aberrant lipid metabolism as a convergent process in both disorders, while the most statistically significant shared pathway, “HIF-1 signaling pathway”, implicates hypoxia—a condition known to be pivotal in both PD [79] and CD [80-82]—as a critical nexus linking mitochondrial dysfunction, oxidative stress, and impaired protein degradation [83].

**Figure 4.**
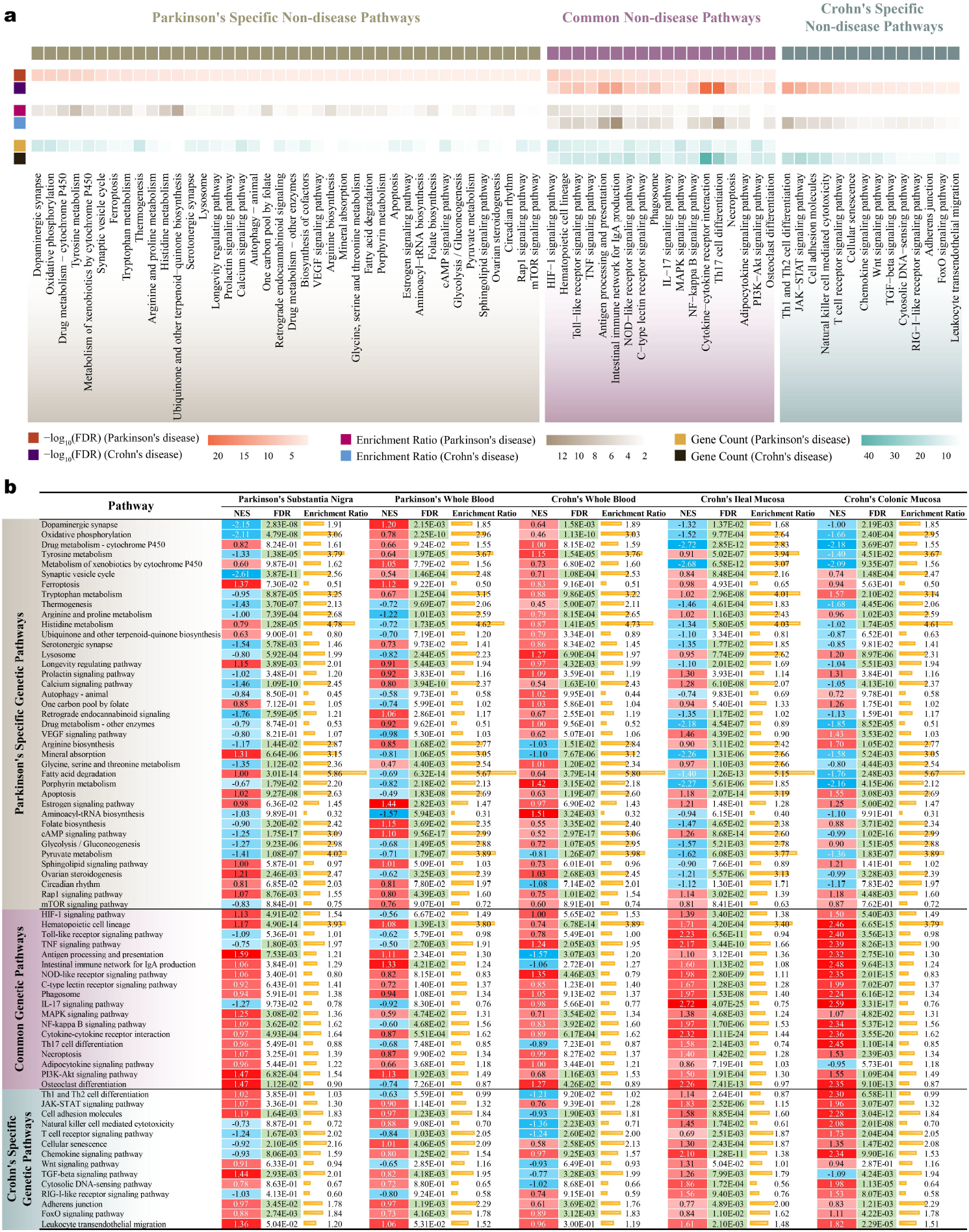
Genetic variant-enriched pathways are transcriptionally active across tissues in PD and CD. **(a)** Enriched non-disease pathways for genetic variants in PD and CD. This pathway enrichment was done against biological pathways within KEGG non-disease categories by ORA approach, with FDR, enrichment ratio and gene counts shown in PD and CD specific and common pathway classes. **(b)** Tissue-wise transcriptional activities of genetic variant-enriched pathways in PD and CD. NES is used to quantify transcriptional activities of genetic pathways in PD and CD tissues, with red shading representing activation and blue shading repression. FDR with green shading indicates corresponding activity quantification is statistically significant.

An exploration of the disease-specific pathways also yielded profound insights into both distinct and convergent mechanisms. For PD, the unique enrichment of “Ferroptosis” aligns with its known role in dopaminergic neuron death and glial activation [84], while “Thermogenesis” provides a molecular correlate for the clinical observation of temperature sensitivity in PD patients [85-87]. Intriguingly, longevity-regulating pathways were enriched in both diseases, incl. “Longevity regulating pathway” and “mTOR signaling pathway” for PD, and “FoxO signaling pathway” for CD, which suggests a deeper shared connection to cellular aging processes that warrants further investigation. The PD specific enrichment of “Circadian rhythm” supports the widespread prevalence of sleep and circadian disruptions in these patients [88,89]. Crucially, several pathways converged on the theme of cell adhesion. The PD specific “Rap1 signaling pathway”, which governs cell-cell junction formation via integrin regulation, alongside the CD specific “Cell adhesion molecules” and “Adherens junction” pathways, collectively point to compromised barrier function as a fundamental, albeit genetically distinct feature. This is further reinforced by the PD specific “VEGF signaling pathway”, a known mediator of blood-brain barrier damage [72]. Together, these results illustrate a complex etiological landscape where both shared and disease-specific pathways ultimately converge upon common pathological themes, suggesting a multifaceted basis for the comorbidity.

To elucidate if these genetically-implicated pathways were functionally active *in vivo*, we assessed their transcriptional activities across five disease-relevant tissues. A striking pattern emerged where PD and CD genetic pathways were significantly engaged in the pathological processes of all five tissues (Figure 4b). PD specific pathways were pervasively downregulated in both substantia nigra and blood, indicative of a systemic degenerative state in PD. Notably, “Dopaminergic synapse” and “Serotonergic synapse” were suppressed not only in the PD substantia nigra but also in the CD ileal and colonic mucosa. Given the direct communication between the brain and gut via the vagus nerve [90], this concurrent downregulation suggests a potential trans-synaptic or neuro-inflammatory link between these distant tissues. PD specific “VEGF signaling pathway” was significantly upregulated in the CD ileal and colonic mucosa. As a non-pathogenic pathway in these contexts, its activation may represent a compensatory response to repair the compromised mucosal barrier [91,92]. CD specific pathways were generally upregulated in the CD intestinal mucosa but displayed divergent activity patterns in CD blood, suggesting distinct functional roles in systemic versus local compartments. Interestingly, CD specific “Cell adhesion molecules” and “Adherens junction” pathways were significantly activated in the PD substantia nigra and blood. Since their activation can be a reparative response [93], this signature hints at a pre-existing or ongoing compromise of the blood-brain barrier in PD. Finally, the shared genetic pathways were consistently activated across all five tissues, strongly suggesting a synergistic interplay that underpins the comorbidity.

To move beyond isolated analyses and investigate the coordinated behavior of the genetic association genes (313 genes for PD, 319 for CD, and 31 for common), we calculated the transcriptional correlation of four gene sets, i.e., shared genes, PD specific genes, CD specific genes, and all combined genes, within and across tissues using the SMIC algorithm. Within the blood, a shared tissue, all four gene sets displayed positive correlation, with the shared gene set reaching statistical significance (SMIC = 0.33, *P* < 0.05), indicating a functional mechanistic link within the circulatory system (Figure S4). As expected, correlations between the CD ileal and colonic mucosa were exceptionally high across all gene sets (SMIC > 0.3, *P* < 0.001), confirming their profound molecular similarity. An unexpected finding emerged from the cross-disease analysis: a consistent, positive correlation was observed between the CD blood and the PD substantia nigra for all four gene sets, reaching statistical significance for the CD specific and combined gene sets (*P* < 0.05 and *P* < 0.01). In contrast, the correlation between PD blood and substantia nigra was inconsistent. This critical asymmetry suggests that the pathological state of the blood in CD, more so than in PD itself, is transcriptionally aligned with the pathological processes occurring in the Parkinsonian brain.

We then extended this correlational analysis to the pathway level, applying SMIC to the transcriptional activities of four corresponding sets of genetic pathways. While this again confirmed the strong positive correlation between the CD ileal and colonic mucosa, other intra- and inter-tissue comparisons failed to yield consistent or significant results (Figure S5), suggesting that simple correlation may be insufficient to capture the complexity of disease linkage. We therefore deployed the ACS method to quantify synergy, a more nuanced measure of synergistic activity. The ACS results revealed widespread synergistic relationships among all five disease tissues (Figure S6), implying a high degree of functional influence.

Recognizing that pathway-level activity scores (i.e., NES in this study) can obscure the individual contributions of member genes, we performed a more granular analysis by calculating both SMIC-based correlation and ACS-based synergy for each genetic pathway based on the differential expression of its constituent genes. The correlation analysis again highlighted the strong, consistent relationship between the CD ileum and colon but yielded few other significant findings (Figure S7a). However, the synergy analysis produced several compelling insights (Figure S7b). Beyond the robust synergy between the CD ileal and colonic mucosa, we observed significant synergy for shared genetic pathways between PD and CD blood, indicating a direct cooperative interaction within the circulation. More critically, a high degree of synergy for shared pathways was detected between PD substantia nigra and CD blood, reinforcing the hypothesis that CD systemic factors directly and synergistically impact central nervous system pathology in PD. Finally, we observed heightened synergistic scores for CD specific pathways between PD substantia nigra and CD colonic mucosa, providing further evidence for a direct gut-brain axis of interaction that contributes to the shared disease landscape.

### 3.5. Increased GEB permeability facilitates ileal and colonic mucosal pathologies in CD

Building upon the above discovery of intricate multi-tissue functional associations between PD and CD, we next sought to establish a plausible physical basis for this pathological crosstalk. Given that PD and CD are canonically defined by pathology primarily localized to the brain and intestine, respectively [94,95], any molecular dialogue between them must navigate the gut-brain axis. This axis is anatomically fortified by a series of three sequential biological barriers: GEB, GVB, and BBB. We therefore posited a sequential-breach hypothesis, wherein molecular pathology originating in the CD gut must progressively traverse these three checkpoints to exert a tangible influence on neuropathogenesis in the PD brain. The compromise of these barriers would thus represent the fundamental physical conduit for the observed comorbidity. Accordingly, our investigation first focused on interrogating the integrity of the initial line of defense, GEB, within the context of active CD pathology.

To empirically test this, we explored the transcriptomic profiles of the ileal and colonic mucosa from CD patients, focusing on the curated GEB permeability biomarker panel. Our analysis revealed that a striking majority of these markers exhibited expression changes that were not only statistically significant but also directionally consistent with the states indicative of impaired barrier function (Figure 5a). This provides compelling molecular evidence for the structural disruption of intercellular tight and adherens junctions in the CD gut, leading to heightened epithelial permeability. This breach effectively dismantles the first physical safeguard, permitting the translocation of luminal contents into the lamina propria (Figure 5b). While the expression of most markers aligned with this conclusion, three biomarkers—claudin 1, claudin 2, and JAM3—displayed a consistent and significant transcriptional upregulation, a finding seemingly at odds with their protein-level functions in maintaining barrier integrity. As these are not CD susceptibility genes, we propose this transcriptional induction may represent a compensatory, albeit insufficient, response to protein-level degradation or mislocalization, a complex regulatory feedback loop that warrants further mechanistic elucidation.

**Figure 5.**
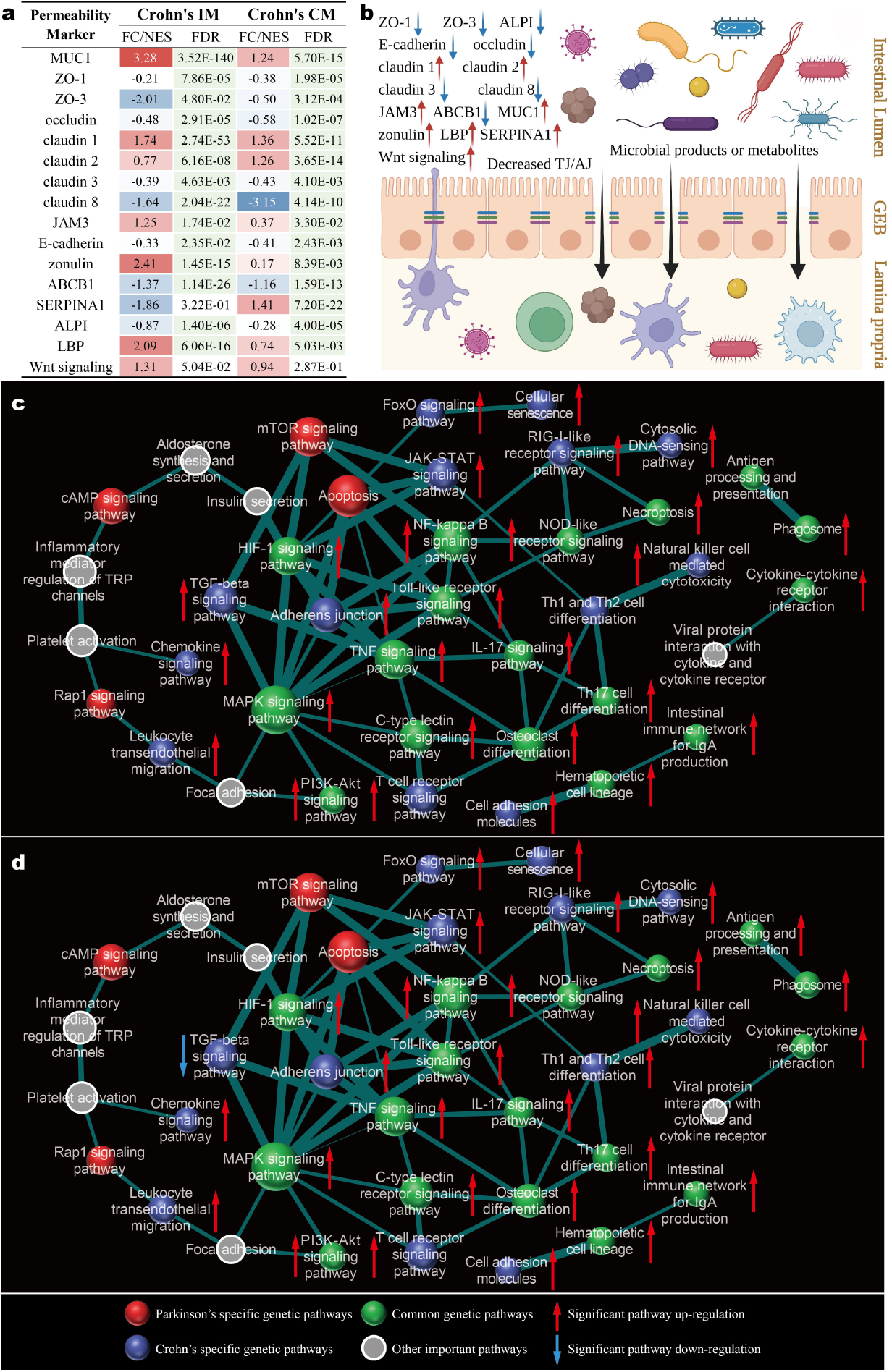
Increased GEB permeability facilitates ileal and colonic mucosal pathologies in CD. **(a)** Transcriptional change profile of gut epithelial barrier permeability biomarkers in ileal and colonic mucosa of CD. **(b)** Schematic diagram of the mechanism underlying increased gut epithelial barrier permeability in the ileum and colon of CD. **(c)** Biological pathway network in ileal mucosa of CD. **(d)** Biological pathway network in colonic mucosa of CD. The thickness of the edges between pathway nodes represents the pathway crosstalk intensity: the thicker the edge, the greater the crosstalk intensity. The pathway node size denotes the network node degree: the larger the node, the higher the node degree. IM, ileal mucosa. CM, colonic mucosa. TJ, tight junction. AJ, adherens junction.

The consequence of this compromised GEB is an influx of microbial products and other luminal antigens into the intestinal wall, a perturbation expected to profoundly remodel the local biological network. To visualize these down-stream effects, we integrated our transcriptomic activity data (pathway NES and *P*-values) with the foundational biological pathway network (“*Human biological pathway network construction*” in Materials and Methods). By filtering for significantly perturbed pathways while preserving network connectivity, we constructed context-specific pathway networks for both the CD ileum and colon (Figure 5c, d). The resultant networks were remarkably congruent, corroborating our prior correlational analyses and underscoring the profound mechanistic similarity between the two intestinal sites. Intriguingly, these networks retained several pathways without direct genetic links to CD, including some with PD specific genetic underpinnings. Their topological persistence reveals their critical role as indispensable functional bridges that connect otherwise disconnected hubs of significantly altered CD pathways. The specific inclusion of PD genetic pathways as such conduits within the CD intestinal milieu provides compelling evidence for a latent molecular comorbidity at the gut level. While direct investigation of PD-related pathology in the ileum and colon remains a nascent field, our findings lay a novel conceptual groundwork and provide a strong impetus for future research into this unexplored facet of the gut-brain axis.

### 3.6. GVB dysfunction underlies the blood-borne comorbidity of PD and CD

We next assessed the integrity of GVB, the second critical checkpoint in the gut-brain axis. To this end, we analyzed blood transcriptomes from both PD and CD cohorts for permeability biomarker signatures. This analysis revealed a compelling transcriptomic profile of GVB impairment in CD, with most biomarkers showing statistically robust expression changes indicative of increased permeability; conversely, these changes were largely non-significant in PD, suggesting the GVB remains substantially intact in this condition (Figure 6a). The paradoxical upregulation of claudin 5 in CD blood, a potential compensatory response, was a notable exception. These transcriptomic signatures strongly suggest a compromised GVB in CD, characterized by the disruption of its intercellular junctions, which facilitates the infiltration of gut-derived molecules into the bloodstream (Figure 6b).

**Figure 6.**
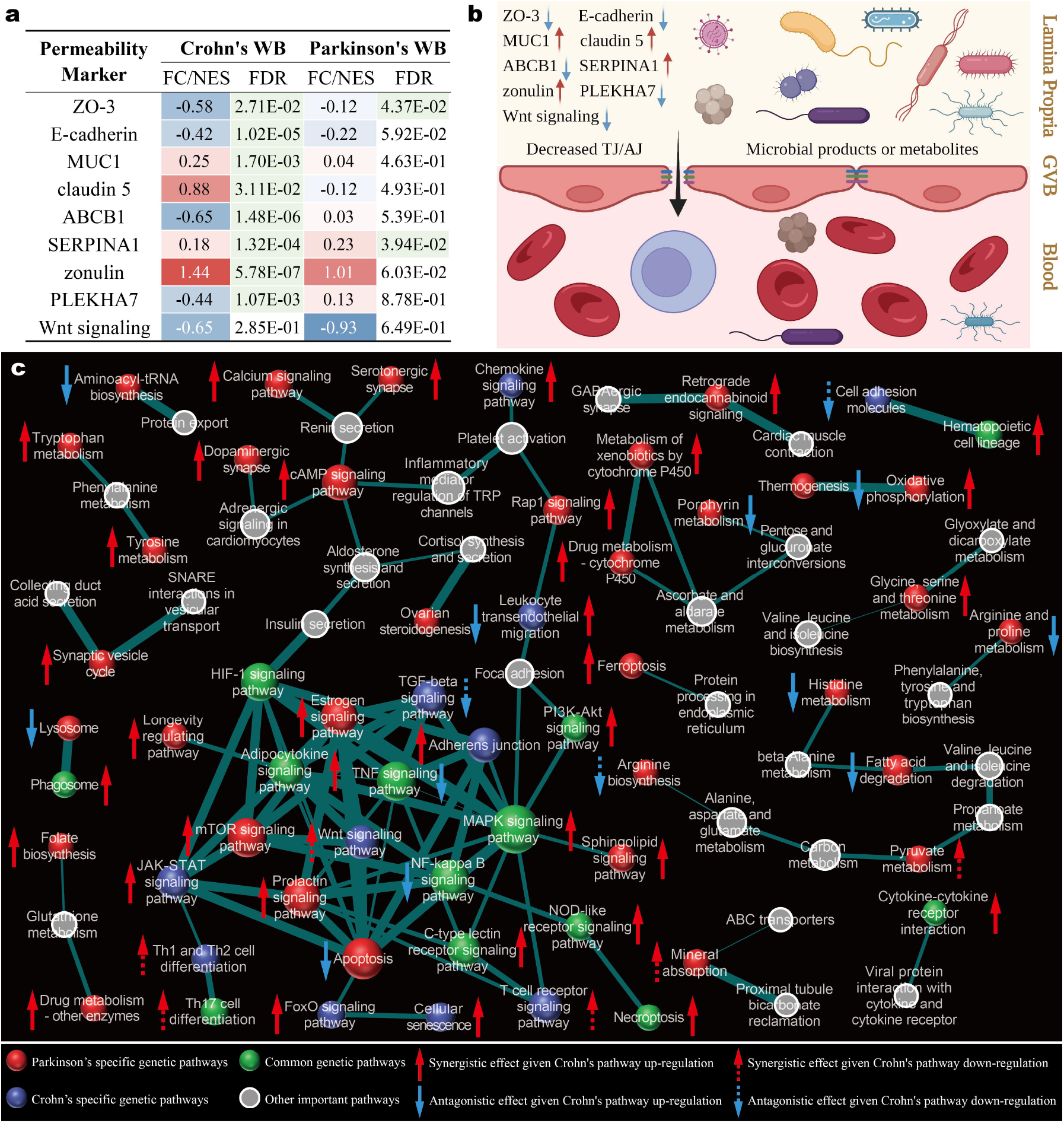
GVB dysfunction underlies the blood-borne comorbidity of PD and CD. **(a)** Transcriptional change profile of gut-vascular barrier permeability biomarkers in blood of PD and CD. **(b)** Schematic diagram of the mechanism underlying increased gut-vascular barrier permeability in PD and CD. **(c)** Biological pathway network in blood of PD and CD. The thickness of the edges between pathway nodes represents the pathway crosstalk intensity: the thicker the edge, the greater the crosstalk intensity. The pathway node size denotes the network node degree: the larger the node, the higher the node degree. WB, whole blood. TJ, tight junction. AJ, adherens junction.

To model the systemic consequences of GVB breach in CD, we constructed the blood-specific biological landscape for both PD and CD (Figure 6c) by integrating transcriptomic activity onto the foundational pathway network. We then classified interaction for any same blood-borne pathway in PD and CD as either synergistic (both up-or down-regulated) or antagonistic (divergently regulated). The synergistic mode would theoretically promote comorbidity, while the antagonistic mode would counteract it. The landscape was overwhelmingly dominated by synergistic effects, revealing a powerful systemic synergy. Antagonistic interactions were rare and confined to the network’s periphery, exerting negligible global influence. This systemic synergy provides a molecular basis for how the pathological milieu in CD blood could create a permissive environment for PD pathogenesis. Notably, the network’s structural integrity relied on several non-genetically associated “bridge” pathways, highlighting their critical role in mediating this pathological crosstalk and marking them as targets for future investigation.

### 3.7. CD blood signature promotes substantia nigra degeneration in PD through BBB disruption

Our investigation culminated at the final and most critical gatekeeper of the gut-brain axis, BBB. We sought to determine if this terminal interface is breached under the pathological conditions of CD or PD, thereby permitting the influx of peripheral molecules into the brain parenchyma.

A transcriptomic interrogation of permeability biomarkers yielded unambiguous evidence (Figure 7a). In the blood of patients with CD, the entire panel of BBB markers exhibited expression changes that were both statistically robust and directionally consistent with compromised barrier integrity. In contrast, these markers remained largely quiescent in PD blood, suggesting that the systemic milieu in PD does not induce a BBB breach. However, a direct analysis of the PD substantia nigra painted a different picture, revealing a significant local disruption of BBB. This dual-front assault on BBB—a systemic challenge from the CD periphery and a localized compromise from intrinsic PD pathology—creates a permissive gateway for pathological crosstalk (Figure 7b). Notably, the paradoxical transcriptional upregulation of claudin 5 and JAM3 within the PD substantia nigra suggests a compensatory response to protein-level junctional instability.

**Figure 7.**
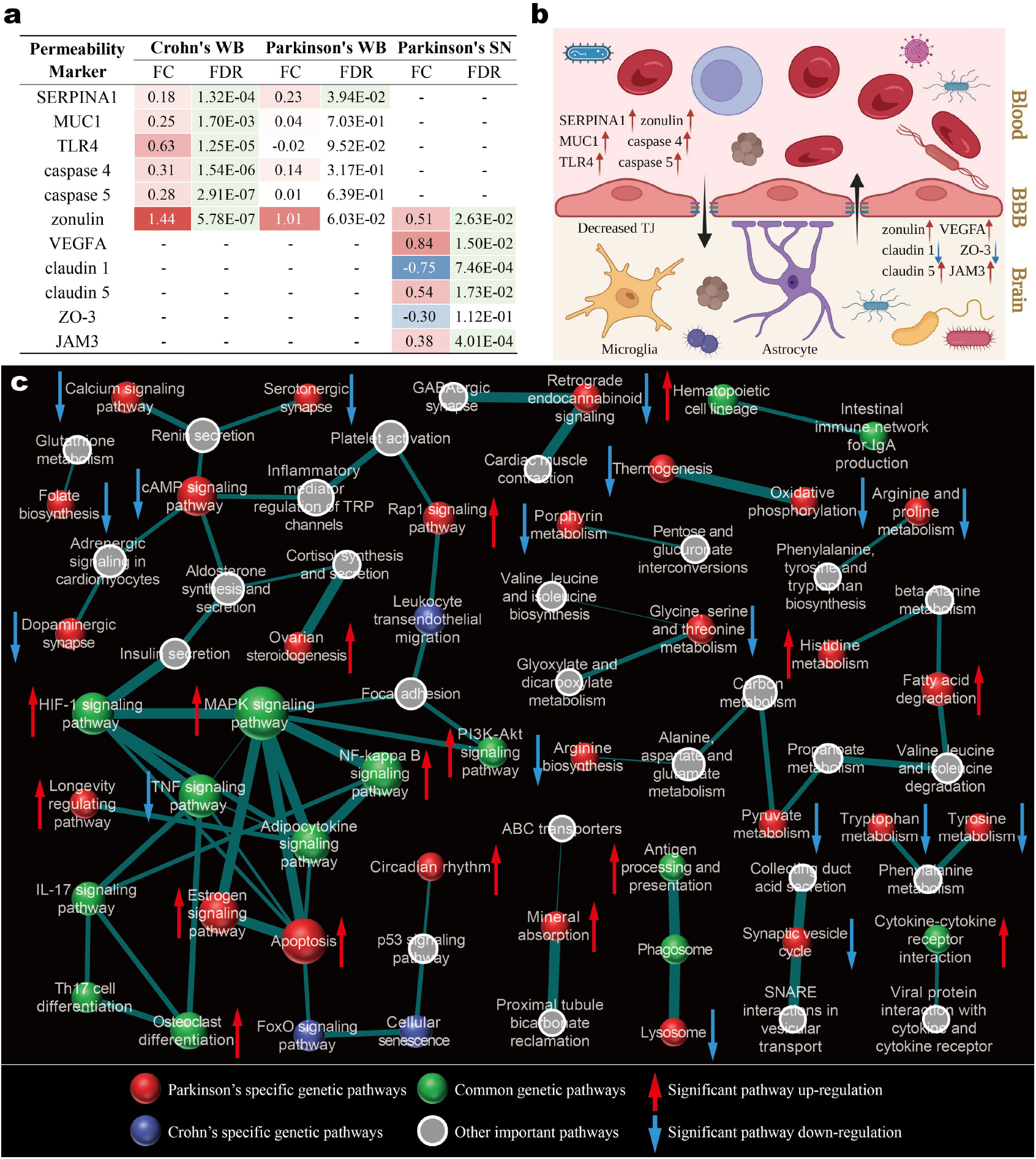
CD blood signature promotes substantia nigra degeneration in PD through BBB disruption. **(a)** Transcriptional change profile of blood-brain barrier permeability biomarkers in blood of PD and CD, and PD substantia nigra. **(b)** Schematic diagram of the mechanism underlying increased blood-brain barrier permeability in PD and CD. **(c)** Biological pathway network in PD substantia nigra. The thickness of the edges between pathway nodes represents the pathway crosstalk intensity: the thicker the edge, the greater the crosstalk intensity. The pathway node size denotes the network node degree: the larger the node, the higher the node degree. WB, whole blood. SN, substantia nigra. TJ, tight junction.

To delineate the functional architecture of the diseased brain following BBB breach, we constructed a substantia nigra-specific pathological network (Figure 7c) by integrating transcriptomic activity data onto the foundational pathway landscape. The resulting network was dominated by pervasive pathway downregulations, a molecular portrait of the degenerative state characteristic of the PD brain. Critically, the network’s structural integrity was maintained by several topological keystones, including pathways with specific genetic links to CD. These pathways, though not directly implicated in PD genetics, serve as indispensable functional conduits connecting significantly altered PD-centric hubs. The presence of these CD specific pathways within the brain pathological network in PD provides a tantalizing glimpse into a latent molecular comorbidity within the brain itself. This finding posits a novel mechanism for the comorbidity, suggesting that systemic factors in CD may directly engage and exacerbate the intrinsic pathological cascades of PD, a hypothesis that opens a new frontier for research into the gut-brain connection in neurodegeneration.

## 4. Discussion and Conclusions

In this study, the multi-omics systems biology investigation, spanning from genetic variants to tissue-specific transcriptional networks, converges on a cohesive and compelling mechanistic interpretation for the comorbidity between PD and CD. Based on our findings, a gut-blood-brain axis model is proposed to elucidate a plausible, directional cascade through which peripheral intestinal pathology in CD may promote PD neurodegeneration (Figure 8). This model posits a sequential breach of three critical biological barriers. The process is initiated by a compromised GEB in CD ileum and colon (Figure 8a), which permits the translocation of luminal antigens and microbial products, thereby remodeling the local mucosal pathobiology (Figure 8b, c). This is followed by the permeabilization of GVB, leading to the systemic dissemination of these pro-inflammatory molecules into the bloodstream (Figure 8d). Our analysis reveals that this creates a systemic milieu in CD blood that not only acts synergistically with that in PD blood (Figure 8e) but also is transcriptionally aligned with the pathological state of PD substantia nigra. The cascade culminates in the compromise of BBB, a dual-front assault driven by both systemic factors from CD periphery and intrinsic pathology within the PD substantia nigra (Figure 8f), ultimately allowing peripheral insults to engage and exacerbate central nervous system degeneration (Figure 8g). This model provides, for the first time, a tangible physical and molecular basis for the well-documented epidemiological link between these two seemingly disparate disorders.

**Figure 8.**
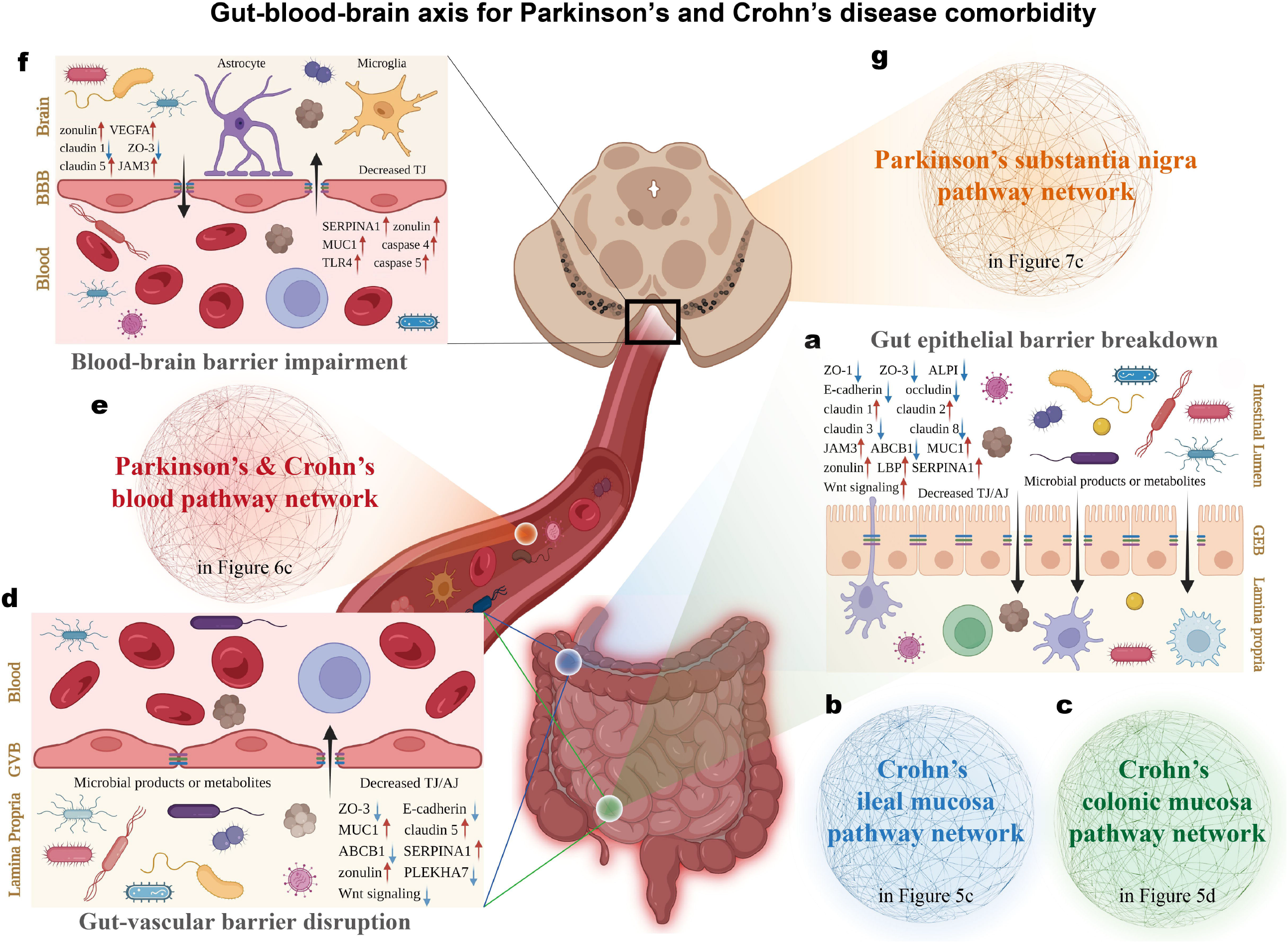
Gut-blood-brain axis model for the comorbidity between PD and CD. **(a)** GEBs of the ileal and colonic mucosa are impaired in the molecular pathological context of CD. **(b, c)** The biological pathway networks in CD ileal and colonic mucosa are established following GEB breakdown. **(d)** GVB disruption causes microorganisms and their derivatives to cross the barrier and enter the blood circulation. **(e)** Blood pathway network synergy of PD and CD after GVB disruption. **(f)** BBB impairment against the background of CD related blood pathology, enables blood contents to cross the blockade and enter the brain parenchyma. **(g)** The biological pathway network in PD substantia nigra is influenced following BBB impairment. TJ, tight junction. AJ, adherens junction.

A key strength of our study lies in its integrative, systems-level design, which bridges the gap between static genetic risk and dynamic, tissue-specific functional consequences. By employing novel computational approaches such as SMIC and newly proposed ACS, we were able to move beyond simple correlation to uncover complex, non-linear relationships and synergistic interplay between pathways and tissues. This methodology provides a powerful blueprint for investigating other complex comorbidities, particularly those spanning the gut-brain axis. Our findings redefine the etiological landscape of PD-CD comorbidity by positioning peripheral inflammatory conditions not merely as risk factors, but as active participants in neuropathogenesis, offering a paradigm shift that could inform new diagnostic and therapeutic strategies focused on maintaining gut barrier integrity and mitigating systemic inflammation to preserve neurological health.

While our data-driven discoveries provide valuable insights, we acknowledge several limitations that chart a clear course for future research. Foremost, our findings are computationally derived and constitute a rich portfolio of data-driven hypotheses that await rigorous experimental validation. Methodologically, our ACS metric represents a prototype approach; future iterations could incorporate the magnitude of gene expression changes, not just the direction, to achieve a more quantitative measure of synergy. Furthermore, validation across multi-omics layers—including proteomics and metabolomics—is imperative to capture post-transcriptional nuances and confirm that the observed transcriptional synergies translate to the functional protein and metabolite levels. The current analysis is also constrained by the availability of matched tissue datasets. Acquiring transcriptomic data from the substantia nigra in CD and the PD intestinal mucosa represents a critical next step to complete the mechanistic puzzle. Finally, leveraging the power of single-cell and spatial transcriptomics and proteomics will be crucial for dissecting the cell type- and niche-specific contributions to barrier dysfunction, and intra- and inter-tissue crosstalk, elevating the resolution of our proposed comorbidity model from the tissue to the cellular level.

In conclusion, this study transitions the understanding of PD and CD comorbidity from epidemiological correlation to a mechanistically defined gut-blood-brain axis. By delineating the trajectory from shared genetic defects in barrier maintenance to a sequential collapse of physiological boundaries, we demonstrate how peripheral intestinal pathology can transcriptionally imprint upon the central nervous system in the context of PD and CD comorbidity. Ultimately, beyond deciphering this specific pathogenic nexus, the analytical framework established herein stands as a scalable paradigm for unraveling the molecular routes bridging anatomically distinct diseases.

## Data Availability Statement

The data that support the findings of this study are available in the supplementary material of this article.

## Author Contributions

**Yanshi Hu**: Conceptualization, Investigation, Methodology, Data curation, Formal analysis, Writing - original draft, Writing - review & editing. **Bentao She**: Data curation, Investigation, Writing - review & editing. **Zhounan Yin**: Data curation, Writing - review & editing. **Xinjian Yu**: Data curation, Writing - review & editing. **Wenyi Wu**: Data curation, Writing - review & editing. **Ming Chen**: Conceptualization, Supervision, Funding acquisition, Project administration, Writing - review & editing.

## Acknowledgment

The authors are grateful to the editors for handling this manuscript, the domain experts for insightful reviews, and the members of Ming Chen’s laboratory for helpful discussions and valuable comments.

## Abbreviations

The following abbreviations are used in this manuscript:

PD: Parkinson’s disease
CD: Crohn’s disease
IBD: Inflammatory bowel disease
DEG: Differentially expressed gene
GWAS: Genome-wide association study
GO: Gene Ontology
KEGG: Kyoto Encyclopedia of Genes and Genomes
ORA: Over-representation analysis
BP: Biological Process
CC: Cellular Component
MF: Molecular Function
FDR: False discovery rate
FC: Fold change
GSEA: Gene set enrichment analysis
NES: Normalized enrichment score
MIC: Maximal information coefficient
SMIC: Signed maximal information coefficient
ACS: Acting-in-concert score
GEB: Gut epithelial barrier
GVB: Gut-vascular barrier
BBB: Blood-brain barrier
IM: Ileal mucosa
CM: Colonic mucosa
TJ: Tight junction
AJ: Adherens junction
WB: Whole blood
SN: Substantia nigra

## References

1. Feigin, V.L.; Abajobir, A.A.; Abate, K.H.; Abd-Allah, F.; Abdulle, A.M.; Abera, S.F.; Abyu, G.Y.; Ahmed, M.B.; Aichour, A.N.; Aichour, I. Global, regional, and national burden of neurological disorders during 1990–2015: A systematic analysis for the global burden of disease study 2015. Lancet Neurol. 2017, 16, 877–897.

2. Nalls, M.A.; Blauwendraat, C.; Vallerga, C.L.; Heilbron, K.; Bandres-Ciga, S.; Chang, D.; Tan, M.; Kia, D.A.; Noyce, A.J.; Xue, A., et al. Identification of novel risk loci, causal insights, and heritable risk for parkinson’s disease: A meta-analysis of genome-wide association studies. Lancet Neurol. 2019, 18, 1091–1102.

3. Chang, D.; Nalls, M.A.; Hallgrímsdóttir, I.B.; Hunkapiller, J.; van der Brug, M.; Cai, F.; Kerchner, G.A.; Ayalon, G.; Bingol, B.; Sheng, M., et al. A meta-analysis of genome-wide association studies identifies 17 new parkinson’s disease risk loci. Nat. Genet. 2017, 49, 1511–1516.

4. Baumgart, D.C.; Sandborn, W.J. Crohn’s disease. The Lancet 2012, 380, 1590–1605.

5. Roda, G.; Chien Ng, S.; Kotze, P.G.; Argollo, M.; Panaccione, R.; Spinelli, A.; Kaser, A.; Peyrin-Biroulet, L.; Danese, S. Crohn’s disease. Nature reviews Disease primers 2020, 6, 22.

6. Shivashankar, R.; Tremaine, W.J.; Harmsen, W.S.; Loftus, E.V. Incidence and prevalence of crohn’s disease and ulcerative colitis in olmsted county, minnesota from 1970 through 2010. Clin. Gastroenterol. Hepatol. 2017, 15, 857–863.

7. Torres, J.; Mehandru, S.; Colombel, J.-F.; Peyrin-Biroulet, L. Crohn’s disease. The Lancet 2017, 389, 1741–1755.

8. Houser, M.C.; Tansey, M.G. The gut-brain axis: Is intestinal inflammation a silent driver of parkinson’s disease pathogenesis? npj Parkinsons Dis. 2017, 3, 3.

9. Liddle, R.A. Parkinson’s disease from the gut. Brain Res. 2018, 1693, 201–206.

10. Weimers, P.; Halfvarson, J.; Sachs, M.C.; Ludvigsson, J.F.; Peter, I.; Olén, O.; Burisch, J. Association between inflammatory bowel disease and parkinson’s disease: Seek and you shall find? Gut 2019, 68, 175–176.

11. Rolli-Derkinderen, M.; Leclair-Visonneau, L.; Bourreille, A.; Coron, E.; Neunlist, M.; Derkinderen, P. Is parkinson’s disease a chronic low-grade inflammatory bowel disease? J. Neurol. 2019, 1–7.

12. Hutfless, S.; Wenning, G.K. Which way does the axis tip? Ibd increases the risk of parkinson’s disease. Gut 2019, 68, 3.

13. Lin, J.-C.; Lin, C.-S.; Hsu, C.-W.; Lin, C.-L.; Kao, C.-H. Association between parkinson’s disease and inflammatory bowel disease: A nationwide taiwanese retrospective cohort study. Inflamm. Bowel Dis. 2016, 22, 1049–1055.

14. Weimers, P.; Halfvarson, J.; Sachs, M.C.; Saunders-Pullman, R.; Ludvigsson, J.F.; Peter, I.; Burisch, J.; Olén, O. Inflammatory bowel disease and parkinson’s disease: A nationwide swedish cohort study. Inflamm. Bowel Dis. 2019, 25, 111–123.

15. Villumsen, M.; Aznar, S.; Pakkenberg, B.; Jess, T.; Brudek, T. Inflammatory bowel disease increases the risk of parkinson’s disease: A danish nationwide cohort study 1977–2014. Gut 2019, 68, 18–24.

16. Peter, I.; Dubinsky, M.; Bressman, S.; Park, A.; Lu, C.; Chen, N.; Wang, A. Anti–tumor necrosis factor therapy and incidence of parkinson disease among patients with inflammatory bowel disease. JAMA Neurol. 2018, 75, 939–946.

17. Park, S.; Kim, J.; Chun, J.; Han, K.; Soh, H.; Kang, E.A.; Lee, H.J.; Im, J.P.; Kim, J.S. Patients with inflammatory bowel disease are at an increased risk of parkinson’s disease: A south korean nationwide population-based study. J Clin Med. 2019, 8, 1191.

18. Nalls, M.A.; Saad, M.; Noyce, A.J.; Keller, M.F.; Schrag, A.; Bestwick, J.P.; Traynor, B.J.; Gibbs, J.R.; Hernandez, D.G.; Cookson, M.R., et al. Genetic comorbidities in parkinson’s disease. Hum. Mol. Genet. 2014, 23, 831–841.

19. Witoelar, A.; Jansen, I.E.; Wang, Y.; Desikan, R.S.; Gibbs, J.R.; Blauwendraat, C.; Thompson, W.K.; Hernandez, D.G.; Djurovic, S.; Schork, A.J., et al. Genome-wide pleiotropy between parkinson disease and autoimmune diseases. JAMA Neurol. 2017, 74, 780–792.

20. Kang, X.; Ploner, A.; Wang, Y.; Ludvigsson, J.F.; Williams, D.M.; Pedersen, N.L.; Wirdefeldt, K. Genetic overlap between parkinson’s disease and inflammatory bowel disease. Brain Communications 2023, 5, fcad002.

21. Kars, M.E.; Wu, Y.; Stenson, P.D.; Cooper, D.N.; Burisch, J.; Peter, I.; Itan, Y. The landscape of rare genetic variation associated with inflammatory bowel disease and parkinson’s disease comorbidity. Genome Med. 2024, 16, 66.

22. Zheng, H.; Qian, X.; Tian, W.; Cao, L. Exploration of the common gene characteristics and molecular mechanism of parkinson’s disease and crohn’s disease from transcriptome data. Brain sciences 2022, 12, 774.

23. Sun, Q.; Li, Y.-J.; Ning, S.-B. Investigating the molecular mechanisms underlying the co-occurrence of parkinson’s disease and inflammatory bowel disease through the integration of multiple datasets. Sci. Rep. 2024, 14, 17028.

24. Hui, K.Y.; Fernandez-Hernandez, H.; Hu, J.; Schaffner, A.; Pankratz, N.; Hsu, N.-Y.; Chuang, L.-S.; Carmi, S.; Villaverde, N.; Li, X. Functional variants in the lrrk2 gene confer shared effects on risk for crohn’s disease and parkinson’s disease. Sci. Transl. Med. 2018, 10, eaai7795.

25. Toro-Domínguez, D.; Villatoro-Garciá, J.A.; Martorell-Marugán, J.; Román-Montoya, Y.; Alarcón-Riquelme, M.E.; Carmona-Saéz, P. A survey of gene expression meta-analysis: Methods and applications. Briefings in Bioinformatics 2021, 22, 1694–1705.

26. Lee, H.-S.; Lobbestael, E.; Vermeire, S.; Sabino, J.; Cleynen, I. Inflammatory bowel disease and parkinson’s disease: Common pathophysiological links. Gut 2021, 70, gutjnl-2020-322429.

27. Karczewski, K.J.; Snyder, M.P. Integrative omics for health and disease. Nat. Rev. Genet. 2018, 19, 299–310.

28. Ramos, E.M.; Hoffman, D.; Junkins, H.A.; Maglott, D.; Phan, L.; Sherry, S.T.; Feolo, M.; Hindorff, L.A. Phenotype–genotype integrator (phegeni): Synthesizing genome-wide association study (gwas) data with existing genomic resources. European Journal of Human Genetics 2013, 22, 144–147.

29. Tam, V.; Patel, N.; Turcotte, M.; Bossé, Y.; Paré, G.; Meyre, D. Benefits and limitations of genome-wide association studies. Nat. Rev. Genet. 2019, 20, 467–484.

30. Dai, S.; You, R.; Lu, Z.; Huang, X.; Mamitsuka, H.; Zhu, S.; Murphy, R. Fullmesh: Improving large-scale mesh indexing with full text. Bioinformatics 2019, 36, 1533–1541.

31. Xun, G.; Jha, K.; Yuan, Y.; Wang, Y.; Zhang, A.; Wren, J. Meshprobenet: A self-attentive probe net for mesh indexing. Bioinformatics 2019, 35, 3794–3802.

32. Hu, Y.; Pan, Z.; Hu, Y.; Zhang, L.; Wang, J. Network and pathway-based analyses of genes associated with parkinson’s disease. Mol. Neurobiol. 2017, 54, 4452–4465.

33. Fujita, P.A.; Rhead, B.; Zweig, A.S.; Hinrichs, A.S.; Karolchik, D.; Cline, M.S.; Goldman, M.; Barber, G.P.; Clawson, H.; Coelho, A., et al. The ucsc genome browser database: Update 2011. Nucleic Acids Res. 2011, 39, D876–D882.

34. Cariaso, M.; Lennon, G. Snpedia: A wiki supporting personal genome annotation, interpretation and analysis. Nucleic Acids Res. 2012, 40, D1308–D1312.

35. Allot, A.; Peng, Y.; Wei, C.-H.; Lee, K.; Phan, L.; Lu, Z. Litvar: A semantic search engine for linking genomic variant data in pubmed and pmc. Nucleic Acids Res. 2018, 46, W530–W536.

36. Landrum, M.J.; Lee, J.M.; Benson, M.; Brown, G.R.; Chao, C.; Chitipiralla, S.; Gu, B.; Hart, J.; Hoffman, D.; Jang, W., et al. Clinvar: Improving access to variant interpretations and supporting evidence. Nucleic Acids Res. 2018, 46, D1062–D1067.

37. Yu, G.; Wang, L.-G.; Han, Y.; He, Q.-Y. Clusterprofiler: An r package for comparing biological themes among gene clusters. OMICS: J. Integrative Biol. 2012, 16, 284–287.

38. Hu, Y.; Yang, Y.; Fang, Z.; Hu, Y.-S.; Zhang, L.; Wang, J. Detecting pathway relationship in the context of human protein-protein interaction network and its application to parkinson’s disease. Methods 2017, 131, 93–103.

39. Hu, Y.-S.; Xin, J.; Hu, Y.; Zhang, L.; Wang, J. Analyzing the genes related to alzheimer’s disease via a network and pathway-based approach. Alzheimers Res. Ther. 2017, 9, 29.

40. Jassal, B.; Matthews, L.; Viteri, G.; Gong, C.; Lorente, P.; Fabregat, A.; Sidiropoulos, K.; Cook, J.; Gillespie, M.; Haw, R., et al. The reactome pathway knowledgebase. Nucleic Acids Res. 2019, 48, D498–D503.

41. Shamir, R.; Klein, C.; Amar, D.; Vollstedt, E.-J.; Bonin, M.; Usenovic, M.; Wong, Y.C.; Maver, A.; Poths, S.; Safer, H., et al. Analysis of blood-based gene expression in idiopathic parkinson disease. Neurology 2017, 89, 1676–1683.

42. Chen, P.; Zhou, G.; Lin, J.; Li, L.; Zeng, Z.; Chen, M.; Zhang, S. Serum biomarkers for inflammatory bowel disease. Frontiers in Medicine 2020, 7, 123.

43. Iturria-Medina, Y.; Khan, A.F.; Adewale, Q.; Shirazi, A.H.; Initiative, t.A.s.D.N. Blood and brain gene expression trajectories mirror neuropathology and clinical deterioration in neurodegeneration. Brain 2020, 143, 661–673.

44. Pai, N.; Palmer, N.P.; Silvester, J.A.; Lee, J.J.; Beam, A.L.; Fried, I.; Valtchinov, V.I.; Rahimov, F.; Kong, S.W.; Ghodoussipour, S., et al. Concordance between gene expression in peripheral whole blood and colonic tissue in children with inflammatory bowel disease. PLoS One 2019, 14, e0222952.

45. Hu, Y.; Wang, Z.; Hu, Y.; Feng, C.; Fang, Q.; Chen, M. Awmeta empowers adaptively-weighted transcriptomic meta-analysis. bioRxiv 2025, 2025.2005.2006.650408.

46. Subramanian, A.; Tamayo, P.; Mootha, V.K.; Mukherjee, S.; Ebert, B.L.; Gillette, M.A.; Paulovich, A.; Pomeroy, S.L.; Golub, T.R.; Lander, E.S., et al. Gene set enrichment analysis: A knowledge-based approach for interpreting genome-wide expression profiles. Proceedings of the National Academy of Sciences 2005, 102, 15545–15550.

47. Reshef, D.N.; Reshef, Y.A.; Finucane, H.K.; Grossman, S.R.; McVean, G.; Turnbaugh, P.J.; Lander, E.S.; Mitzenmacher, M.; Sabeti, P.C. Detecting novel associations in large data sets. Science 2011, 334, 1518–1524.

48. Albanese, D.; Filosi, M.; Visintainer, R.; Riccadonna, S.; Jurman, G.; Furlanello, C. Minerva and minepy: A c engine for the mine suite and its r, python and matlab wrappers. Bioinformatics 2013, 29, 407–408.

49. Sharkey, K.A.; Beck, P.L.; McKay, D.M. Neuroimmunophysiology of the gut: Advances and emerging concepts focusing on the epithelium. Nature Reviews Gastroenterology & Hepatology 2018, 15, 765–784.

50. Brescia, P.; Rescigno, M. The gut vascular barrier: A new player in the gut–liver–brain axis. Trends in Molecular Medicine 2021, 27, 844–855.

51. Kadry, H.; Noorani, B.; Cucullo, L. A blood–brain barrier overview on structure, function, impairment, and biomarkers of integrity. Fluids and Barriers of the CNS 2020, 17, 69.

52. Bell, R.D.; Winkler, E.A.; Singh, I.; Sagare, A.P.; Deane, R.; Wu, Z.; Holtzman, D.M.; Betsholtz, C.; Armulik, A.; Sallstrom, J., et al. Apolipoprotein e controls cerebrovascular integrity via cyclophilin a. Nature 2012, 485, 512–516.

53. Poller, B.; Drewe, J.; Krähenbühl, S.; Huwyler, J.; Gutmann, H. Regulation of bcrp (abcg2) and p-glycoprotein (abcb1) by cytokines in a model of the human blood–brain barrier. Cellular and Molecular Neurobiology 2009, 30, 63–70.

54. Lu, X.; Wang, M.; Qi, J.; Wang, H.; Li, X.; Gupta, D.; Dziarski, R. Peptidoglycan recognition proteins are a new class of human bactericidal proteins. Journal of Biological Chemistry 2006, 281, 5895–5907.

55. Chen, J.; Zhang, C.; Wu, Y.; Zhang, D. Association between hypertension and the risk of parkinson’s disease: A meta-analysis of analytical studies. Neuroepidemiology 2019, 52, 181–192.

56. Mao, Z.; Liu, C.; Ji, S.; Yang, Q.; Ye, H.; Han, H.; Xue, Z. The nlrp3 inflammasome is involved in the pathogenesis of parkinson’s disease in rats. Neurochemical Research 2017, 42, 1104–1115.

57. Villani, A.-C.; Lemire, M.; Fortin, G.; Louis, E.; Silverberg, M.S.; Collette, C.; Baba, N.; Libioulle, C.; Belaiche, J.; Bitton, A., et al. Common variants in the nlrp3 region contribute to crohn’s disease susceptibility. Nature Genetics 2008, 41, 71–76.

58. Stavrou, E.F.; Chatzopoulou, F.; Antonatos, C.; Pappa, P.; Makridou, E.; Oikonomou, K.; Kapsoritakis, A.; Potamianos, P.S.; Karmiris, K.; Tzathas, C., et al. Pharmacogenetic analysis of canonical versus noncanonical pathway of nf-kb in crohn’s disease patients under anti-tumor necrosis factor-α treatment. Pharmacogenetics and Genomics 2022, 32, 235–241.

59. Patel, M.; Singh, S. Apigenin attenuates functional and structural alterations via targeting nf-kb/nrf2 signaling pathway in lps-induced parkinsonism in experimental rats. Neurotoxicity Research 2022, 40, 941–960.

60. Hui, K.Y.; Fernandez-Hernandez, H.; Hu, J.; Schaffner, A.; Pankratz, N.; Hsu, N.Y.; Chuang, L.S.; Carmi, S.; Villaverde, N.; Li, X., et al. Functional variants in the lrrk2 gene confer shared effects on risk for crohn’s disease and parkinson’s disease. Science Translational Medicine 2018, 10, eaai7795.

61. Derkinderen, P.; Neunlist, M. Crohn’s and parkinson disease: Is lrrk2 lurking around the corner? Nature Reviews Gastroenterology & Hepatology 2018, 15, 330–331.

62. Grenn, F.P.; Makarious, M.B.; Bandres-Ciga, S.; Iwaki, H.; Singleton, A.B.; Nalls, M.A.; Blauwendraat, C. Analysis of y chromosome haplogroups in parkinson’s disease. Brain Communications 2022, 4, fcac277.

63. Kanehisa, M.; Sato, Y.; Furumichi, M.; Morishima, K.; Tanabe, M. New approach for understanding genome variations in kegg. Nucleic Acids Res. 2019, 47, D590–D595.

64. Camacho-Soto, A.; Gross, A.; Searles Nielsen, S.; Dey, N.; Racette, B.A. Inflammatory bowel disease and risk of parkinson’s disease in medicare beneficiaries. Parkinsonism & Related Disorders 2018, 50, 23–28.

65. Lee, H.S.; Lobbestael, E.; Vermeire, S.; Sabino, J.; Cleynen, I. Inflammatory bowel disease and parkinson’s disease: Common pathophysiological links. Gut 2021, 70, 408–417.

66. Alecu, I.; Bennett, S.A.L. Dysregulated lipid metabolism and its role in α-synucleinopathy in parkinson’s disease. Frontiers in Neuroscience 2019, 13, 328.

67. Scoville, E.A.; Allaman, M.M.; Brown, C.T.; Motley, A.K.; Horst, S.N.; Williams, C.S.; Koyama, T.; Zhao, Z.; Adams, D.W.; Beaulieu, D.B., et al. Alterations in lipid, amino acid, and energy metabolism distinguish crohn’s disease from ulcerative colitis and control subjects by serum metabolomic profiling. Metabolomics 2017, 14, 17.

68. Davies, A.; Nixon, A.; Tsintzas, K.; Stephens, F.B.; Moran, G.W. Skeletal muscle anabolic and insulin sensitivity responses to a mixed meal in adult patients with active crohn’s disease. Clinical Nutrition ESPEN 2021, 41, 305–313.

69. Athauda, D.; Foltynie, T. Insulin resistance and parkinson’s disease: A new target for disease modification? Progress in Neurobiology 2016, 145-146, 98–120.

70. Anzola, M. Hepatocellular carcinoma: Role of hepatitis b and hepatitis c viruses proteins in hepatocarcinogenesis. Journal of Viral Hepatitis 2004, 11, 383–393.

71. Liao, Y.; Wang, J.; Jaehnig, E.J.; Shi, Z.; Zhang, B. Webgestalt 2019: Gene set analysis toolkit with revamped uis and apis. Nucleic Acids Research 2019, 47, W199–W205.

72. Lan, G.; Wang, P.; Chan, R.B.; Liu, Z.; Yu, Z.; Liu, X.; Yang, Y.; Zhang, J. Astrocytic vegfa: An essential mediator in blood-brain-barrier disruption in parkinson’s disease. Glia 2021, 70, 337–353.

73. Bouziat, R.; Jabri, B. Breaching the gut-vascular barrier. Science 2015, 350, 742–743.

74. Spadoni, I.; Zagato, E.; Bertocchi, A.; Paolinelli, R.; Hot, E.; Di Sabatino, A.; Caprioli, F.; Bottiglieri, L.; Oldani, A.; Viale, G., et al. A gut-vascular barrier controls the systemic dissemination of bacteria. Science 2015, 350, 830–834.

75. Archer, N.K.; Adappa, N.D.; Palmer, J.N.; Cohen, N.A.; Harro, J.M.; Lee, S.K.; Miller, L.S.; Shirtliff, M.E.; Freitag, N.E. Interleukin-17a (il-17a) and il-17f are critical for antimicrobial peptide production and clearance of staphylococcus aureus nasal colonization. Infection and Immunity 2016, 84, 3575–3583.

76. DiNicola, C.A.; Zand, A.; Hommes, D.W. Autologous hematopoietic stem cells for refractory crohn’s disease. Expert Opinion on Biological Therapy 2017, 17, 555–564.

77. Altarche-Xifro, W.; di Vicino, U.; Muñoz-Martin, M.I.; Bortolozzi, A.; Bové, J.; Vila, M.; Cosma, M.P. Functional rescue of dopaminergic neuron loss in parkinson’s disease mice after transplantation of hematopoietic stem and progenitor cells. EBioMedicine 2016, 8, 83–95.

78. Peter, I.; Dubinsky, M.; Bressman, S.; Park, A.; Lu, C.; Chen, N.; Wang, A. Anti–tumor necrosis factor therapy and incidence of parkinson disease among patients with inflammatory bowel disease. JAMA Neurology 2018, 75, 939.

79. Rey, F.; Messa, L.; Maghraby, E.; Casili, G.; Ottolenghi, S.; Barzaghini, B.; Raimondi, M.T.; Cereda, C.; Cuzzocrea, S.; Zuccotti, G., et al. Oxygen sensing in neurodegenerative diseases: Current mechanisms, implication of transcriptional response, and pharmacological modulation. Antioxidants & Redox Signaling 2023, 38, 160–182.

80. Kerber, E.L.; Padberg, C.; Koll, N.; Schuetzhold, V.; Fandrey, J.; Winning, S. The importance of hypoxia-inducible factors (hif-1 and hif-2) for the pathophysiology of inflammatory bowel disease. International Journal of Molecular Sciences 2020, 21, 8551.

81. Sawyer, R.G.; Spengler, M.D.; Adams, R.B.; Pruett, T.L. The peritoneal environment during infection. Annals of Surgery 1991, 213, 253–260.

82. Manresa, M.C.; Taylor, C.T. Hypoxia inducible factor (hif) hydroxylases as regulators of intestinal epithelial barrier function. Cellular and Molecular Gastroenterology and Hepatology 2017, 3, 303–315.

83. Leston Pinilla, L.; Ugun-Klusek, A.; Rutella, S.; De Girolamo, L.A. Hypoxia signaling in parkinson’s disease: There is use in asking “what hif?”. Biology 2021, 10, 723.

84. Wang, Z.-L.; Yuan, L.; Li, W.; Li, J.-Y. Ferroptosis in parkinson’s disease: Glia–neuron crosstalk. Trends in Molecular Medicine 2022, 28, 258–269.

85. Zhong, G.; Bolitho, S.; Grunstein, R.; Naismith, S.L.; Lewis, S.J.G. The relationship between thermoregulation and rem sleep behaviour disorder in parkinson’s disease. PLoS One 2013, 8, e72661.

86. Jost, W.H. Autonomic dysfunction in parkinson’s disease: Cardiovascular symptoms, thermoregulation, and urogenital symptoms. International Review of Neurobiology 2017, 134, 771–785.

87. Coon, E.A.; Low, P.A. Thermoregulation in parkinson disease. Handbook of Clinical Neurology 2018, 157, 715–725.

88. Cheng, W.-Y.; Ho, Y.-S.; Chang, R.C.-C. Linking circadian rhythms to microbiome-gut-brain axis in aging-associated neurodegenerative diseases. Ageing Research Reviews 2022, 78, 101620.

89. Shen, Y.; Lv, Q.-k.; Xie, W.-y.; Gong, S.-y.; Zhuang, S.; Liu, J.-y.; Mao, C.-j.; Liu, C.-f. Circadian disruption and sleep disorders in neurodegeneration. Translational Neurodegeneration 2023, 12, 8.

90. Kaelberer, M.M.; Buchanan, K.L.; Klein, M.E.; Barth, B.B.; Montoya, M.M.; Shen, X.; Bohórquez, D.V. A gut-brain neural circuit for nutrient sensory transduction. Science 2018, 361, eaat5236.

91. Martin-Rodriguez, O.; Gauthier, T.; Bonnefoy, F.; Couturier, M.; Daoui, A.; Chagué, C.; Valmary-Degano, S.; Gay, C.; Saas, P.; Perruche, S. Pro-resolving factors released by macrophages after efferocytosis promote mucosal wound healing in inflammatory bowel disease. Frontiers in Immunology 2021, 12, 754475.

92. Abramov, V.M.; Kosarev, I.V.; Priputnevich, T.V.; Machulin, A.V.; Abashina, T.N.; Chikileva, I.O.; Donetskova, A.D.; Takada, K.; Melnikov, V.G.; Vasilenko, R.N., et al. S-layer protein 2 of vaginal lactobacillus crispatus 2029 enhances growth, differentiation, vegf production and barrier functions in intestinal epithelial cell line caco-2. International Journal of Biological Macromolecules 2021, 189, 410–419.

93. Aggarwal, A.; Singh, I.; Sandhir, R. Protective effect of s-nitrosoglutathione administration against hyperglycemia induced disruption of blood brain barrier is mediated by modulation of tight junction proteins and cell adhesion molecules. Neurochemistry International 2018, 118, 205–216.

94. Poewe, W.; Seppi, K.; Tanner, C.M.; Halliday, G.M.; Brundin, P.; Volkmann, J.; Schrag, A.-E.; Lang, A.E. Parkinson disease. Nature Reviews Disease Primers 2017, 3, 17013.

95. Roda, G.; Chien Ng, S.; Kotze, P.G.; Argollo, M.; Panaccione, R.; Spinelli, A.; Kaser, A.; Peyrin-Biroulet, L.; Danese, S. Crohn’s disease. Nature Reviews Disease Primers 2020, 6, 22.

