## Supplementary Materials for "Mapping the comorbid landscape of Parkinson’s disease and Crohn’s disease along the gut-blood-brain axis"

Table of Contents in Supplementary Material

Supplementary Figures:

Figure S1. Chromosomal distributions and functional consequences of genetic variants in PD and CD.

Figure S2. Enriched disease pathways for genetic variants in PD and CD.

Figure S3. Top 40 enriched GO_BP terms for PD and CD genetic association genes.

Figure S4. Tissue-wise transcriptional correlation of genetically-informed genes in PD and CD.

Figure S5. Tissue-wise transcriptional correlation of genetically-informed pathways in PD and CD.

Figure S6. Tissue-wise transcriptional synergy of genetically-informed pathways in PD and CD.

Figure S7. Genetically-informed-pathway-wise transcriptional correlation and synergy within and across tissues in PD and CD.

Supplementary Tables:

**Table S1.** Detailed information of biomarkers related to GEB permeability.

**Table S2.** Detailed information of biomarkers related to GVB permeability.

**Table S3.** Detailed information of biomarkers related to BBB permeability.

**Table S4.** List of literature-derived variants and genes in PD.

**Table S5.** List of literature-derived variants and genes in CD.

**Table S6.** Detailed information of dbSNP-predicted genes in PD, evidenced by literature and/or AWmeta.

**Table S7.** Detailed information of dbSNP-predicted genes in CD, evidenced by literature and/or AWmeta.

**Table S8**. KEGG disease pathway enrichment of PD genetic association genes.

**Table S9**. KEGG disease pathway enrichment of CD genetic association genes.

**Table S10.** Common GO_BP enrichment for PD and CD genetic association genes.

**Table S11.** GO_MF enrichment of PD genetic association genes.

**Table S12.** GO_MF enrichment of CD genetic association genes.

**Table S13.** GO_CC enrichment of PD genetic association genes.

**Table S14.** GO_CC enrichment of CD genetic association genes.

**Table S15.** GO_BP enrichment of PD genetic association genes.

**Table S16.** GO_BP enrichment of CD genetic association genes.

**Table S17.** KEGG non-disease pathway enrichment of PD genetic association genes.

**Table S18.** KEGG non-disease pathway enrichment of CD genetic association genes.

Note: Table S8–S18 are in Supplementary_Table_S8-S18.xlsx file and not shown here.

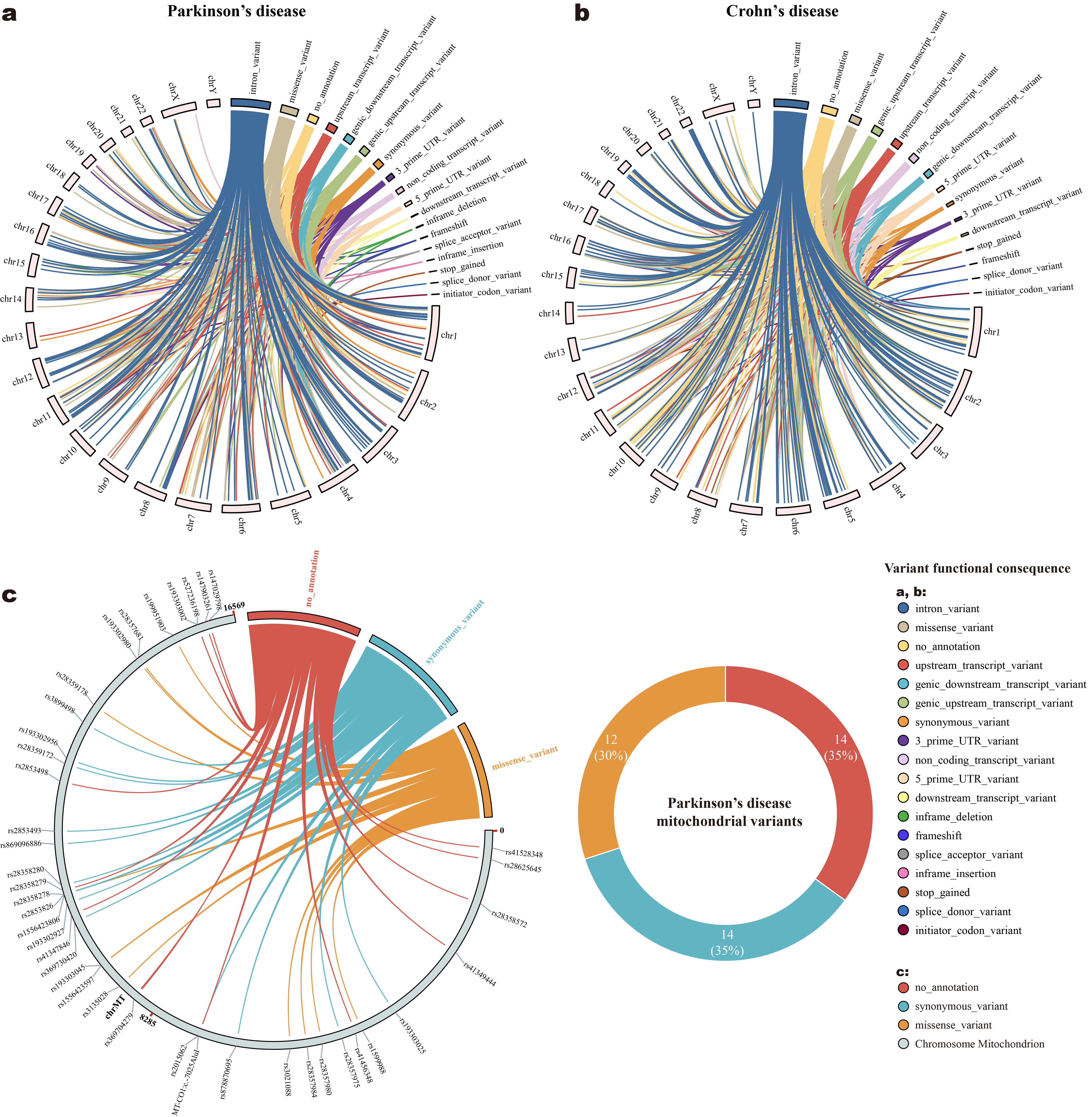

Figure S1. Chromosomal distributions and functional consequences of genetic variants in PD and CD. (a, b) Genetic variant functional consequences in PD and CD are mapped to corresponding chromosomal positions (mitochondrial chromosome excluded). (c) PD mitochondrial variant functional consequences and their respective genomic distributions (left), with three variant functional consequence classes in commensurate frequencies (right).

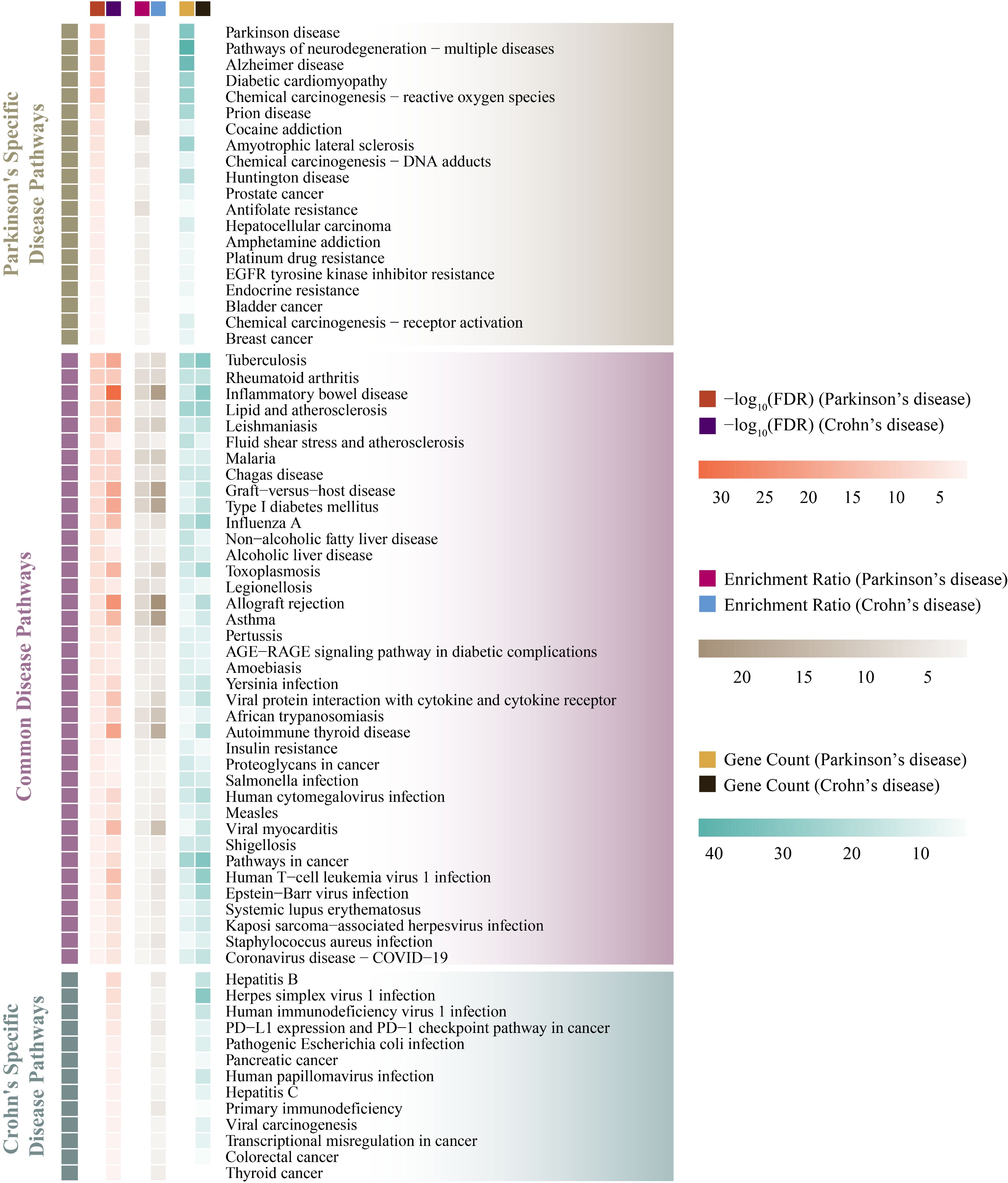

**Figure S2**. Enriched disease pathways for genetic variants in PD and CD. This pathway enrichment was done against biological pathways within KEGG “Human Diseases” category by ORA approach.

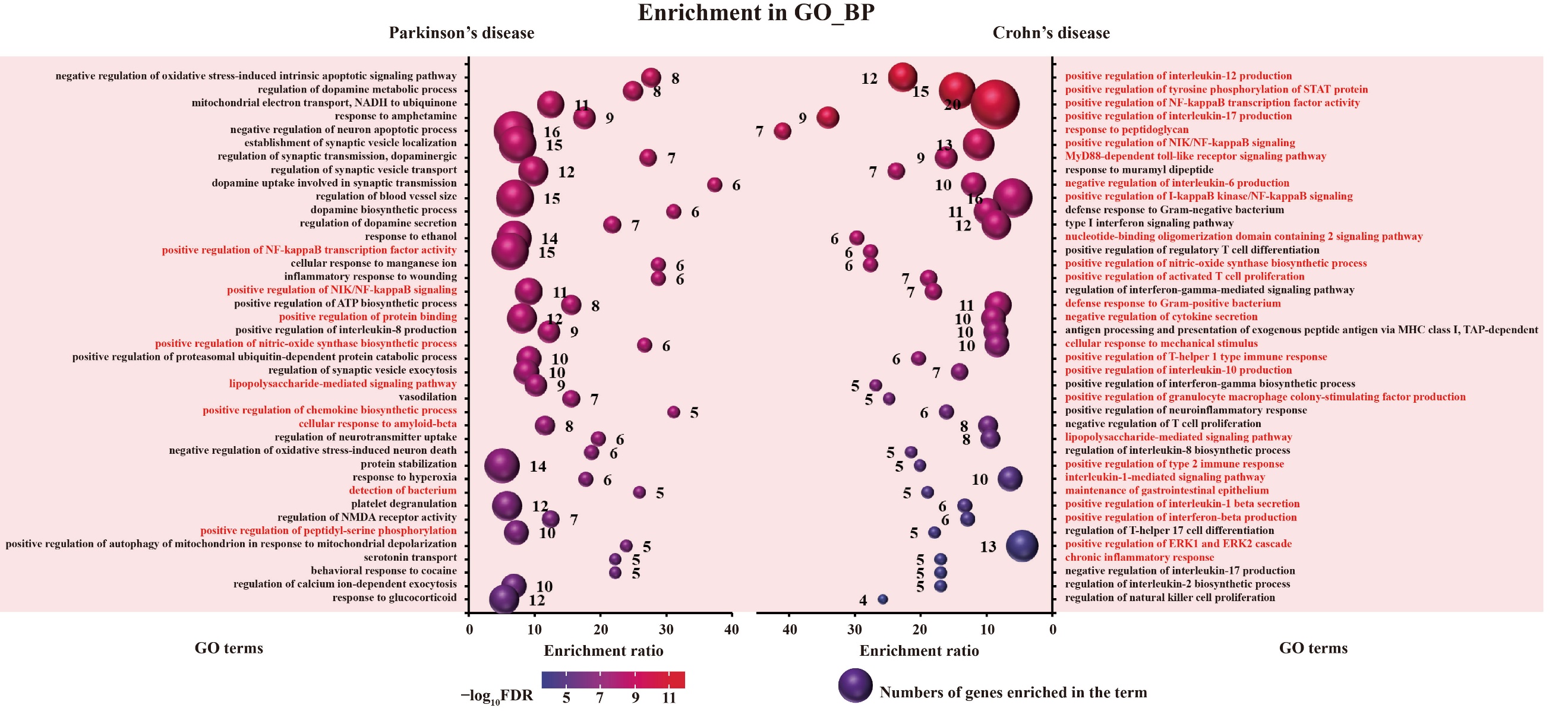

**Figure S3**. Top 40 enriched GO_BP terms for PD and CD genetic association genes. GO term name in red indicates the significantly enriched GO-BP terms shared by PD and CD. The complete results are provided in Tables S15 and S16.

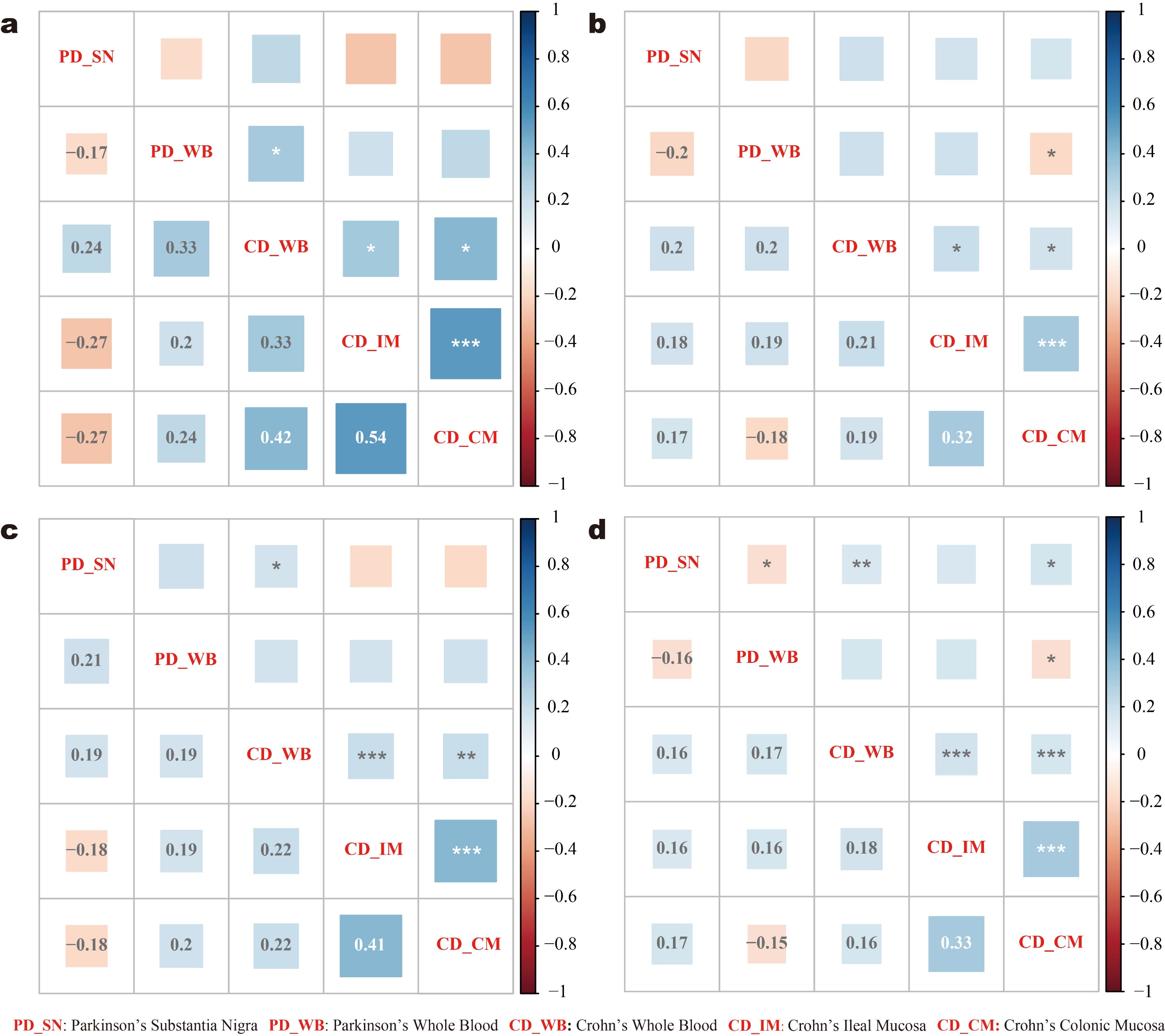

**Figure S4**. Tissue-wise transcriptional correlation of genetically-informed genes in PD and CD. Transcriptional correlation analyses within and across tissues for four gene sets were conducted separately: (**a**) genes genetically associated with both diseases, (**b**) genes genetically associated with PD, (**c**) genes genetically associated with CD, and (**d**) all genes genetically associated with the two diseases. In each correlation result plot, the bottom-left cells below diagonal red texts represent SMIC scores, and the top-right cells above statistical significance, with no asterisk indicating *P* > 0.05, * *P* < 0.05, ** *P* < 0.01, and *** *P* < 0.001.

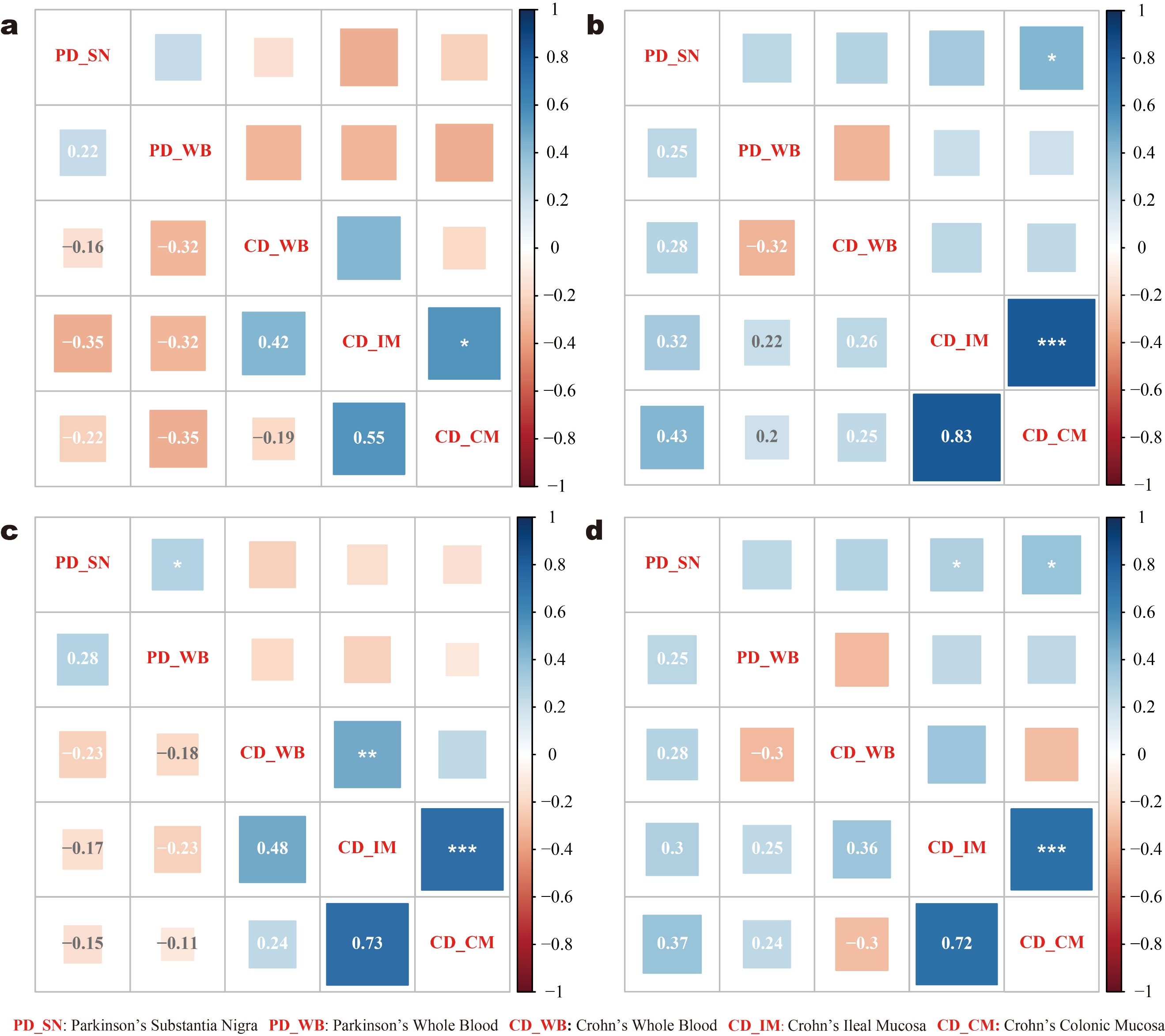

Figure S5. Tissue-wise transcriptional correlation of genetically-informed pathways in PD and CD. Transcriptional correlation analyses within and across tissues for four pathway sets were conducted separately: (a) pathways genetically associated with both diseases, (b) pathways genetically associated with PD, (c) pathways genetically associated with CD, and (d) all pathways genetically associated with the two diseases. In each correlation result plot, the bottom-left cells below diagonal red texts represent SMIC scores, and the top-right cells above statistical significance, with no asterisk indicating *P* > 0.05, * *P* < 0.05, ** *P* < 0.01, and *** *P* < 0.001.

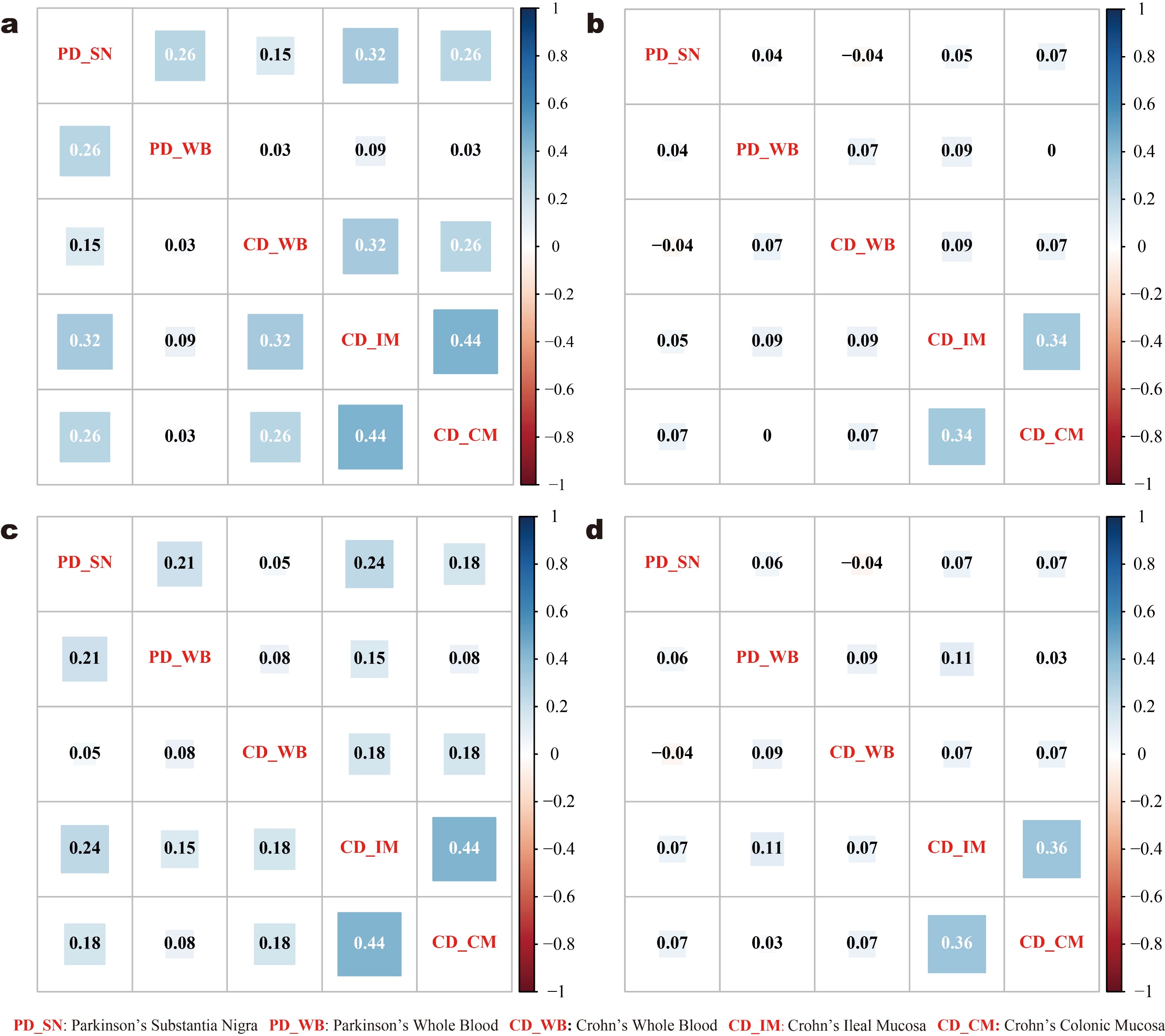

Figure S6. Tissue-wise transcriptional synergy of genetically-informed pathways in PD and CD. Transcriptional synergy analyses within and across tissues for four pathway sets were conducted separately: (a) pathways genetically associated with both diseases, (b) pathways genetically associated with PD, (c) pathways genetically associated with CD, and (d) all pathways genetically associated with the two diseases. In each correlation result plot, every cell represents a ACS score, distributed symmetrically along the diagonal red texts.

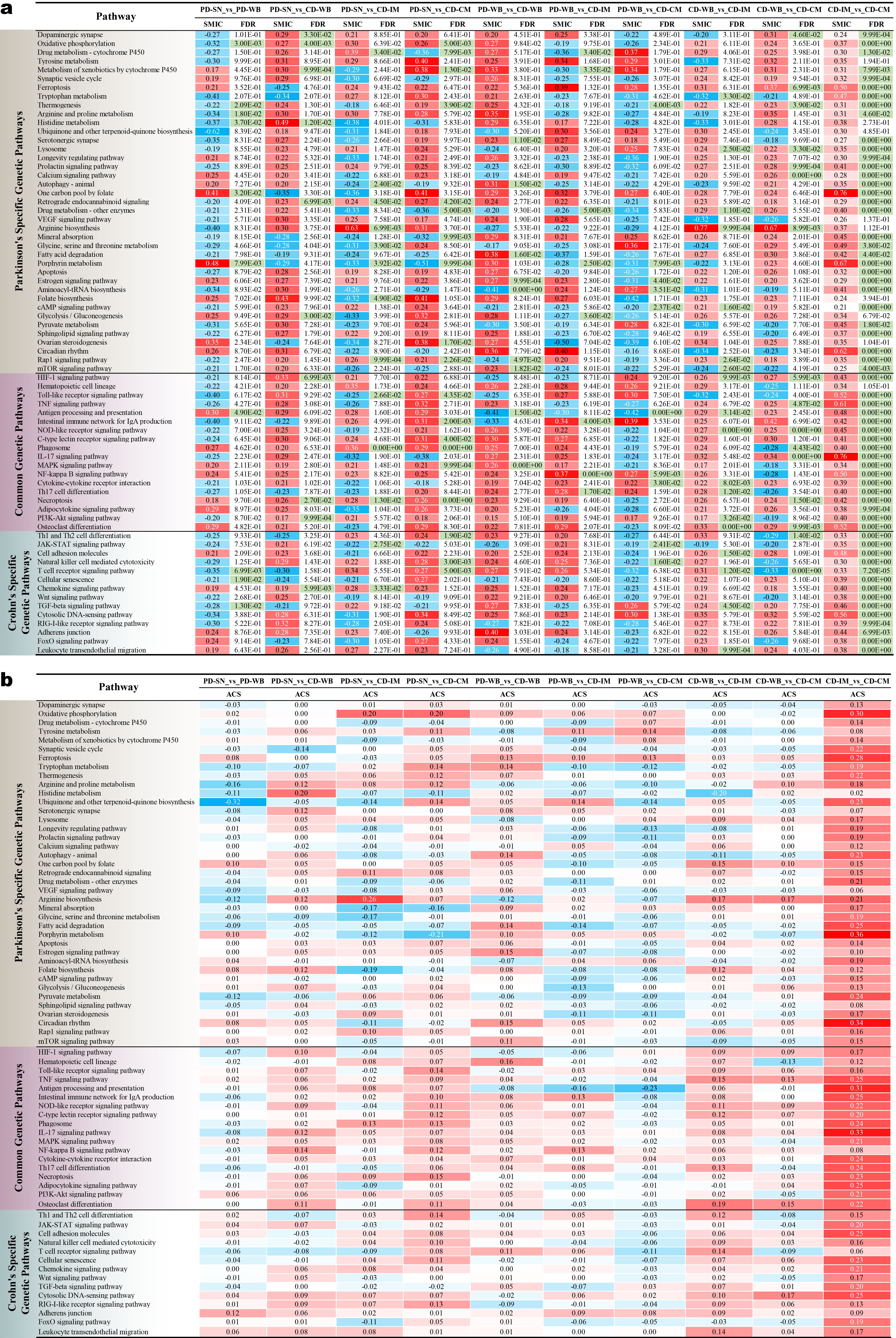

**Figure S7**. Genetically-informed-pathway-wise transcriptional correlation and synergy within and across tissues in PD and CD. (**a**, **b**) Both SMIC-based correlation and ACS-based synergy for each genetic pathway were calculated based on the differential expression of its constituent genes, within and across five disease tissues. Red shading represents positive correlation or synergistic effect, and blue shading negative correlation or antagonistic effect. FDR with green shading indicates corresponding correlation is statistically significant. PD-SN, PD substantia nigra. PD-WB, PD whole blood. CD-WB, CD whole blood. CD-IM, CD ileal mucosa. CD-CM, CD colonic mucosa.

**Table S1.** Detailed information of biomarkers related to GEB permeability.

| **Biomarker type** | **Biomarker** | **Active site** | **Permeability state^a)^** | **Reference^b)^** |
| --- | --- | --- | --- | --- |
| Mucosal layer | MUC1 | Gut mucosa | U | 28885228 |
| Tight junction | ZO-1 | Gut mucosa | D | 26294173 |
| Tight junction | ZO-3 | Gut mucosa | D | 26294173 |
| Tight junction | occludin | Gut mucosa | D | 27848962 |
| Tight junction | claudin 1 | Gut mucosa | D | 23589827 |
| Tight junction | claudin 2 | Gut mucosa | D | 23589827 |
| Tight junction | claudin 3 | Gut mucosa | D | 23589827 |
| Tight junction | claudin 8 | Gut mucosa | D | 23589827 |
| Tight junction | JAM3 | Gut mucosa | D | 27848962 |
| Adherens junction | E-cadherin | Gut mucosa | D | 17934504 |
| Other type | zonulin | Gut mucosa | U | 21248165 |
| Other type | SERPINA1 | Gut mucosa | U | 29454662 |
| Other type | ALPI | Gut mucosa | D | 27106638 |
| Other type | LBP | Gut mucosa | U | 34009040 |
| Other type | ABCB1 | Gut mucosa | D | 28321153 |
| Other type | Wnt signaling | Gut mucosa | U | 26564856 |

^a)^ U represents increased permeability and D decreased permeability.

^b)^ References are shown using PubMed ID (PMID).

**Table S2.** Detailed information of biomarkers related to GVB permeability.

| **Biomarker type** | **Biomarker** | **Active site** | **Permeability state^a)^** | **Reference^b)^** |
| --- | --- | --- | --- | --- |
| Tight junction | ZO-3 | Blood | D | 34229973 |
| Tight junction | claudin 5 | Blood | D | 23589827 |
| Adherens junction | E-cadherin | Blood | D | 17934504 |
| Adherens junction | PLEKHA7 | Blood | D | 17934504 |
| Other type | ABCB1 | Blood | D | 28321153 |
| Other type | SERPINA1 | Blood | U | 29454662 |
| Other type | zonulin | Blood | U | 21248165 |
| Other type | MUC1 | Blood | U | 28885228 |
| Other type | Wnt signaling | Blood | D | 26564856 |

^a)^ U represents increased permeability and D decreased permeability.

^b)^ References are shown using PubMed ID (PMID).

**Table S3.** Detailed information of biomarkers related to BBB permeability.

| **Biomarker type** | **Biomarker** | **Active site** | **Permeability state^a)^** | **Reference^b)^** |
| --- | --- | --- | --- | --- |
| Tight junction | ZO-3 | Substantia nigra | D | 17934504 |
| Tight junction | claudin 1 | Substantia nigra | D | 23589827 |
| Tight junction | claudin 5 | Substantia nigra | D | 23589827 |
| Tight junction | JAM3 | Substantia nigra | D | 27848962 |
| Other type | VEGFA | Substantia nigra | U | 34713920 |
| Other type | zonulin | Substantia nigra  Blood | U | 30449598  32564127 |
| Other type | zonulin |  | U |  |
| Other type | SERPINA1 | Blood | U | 36089165 |
| Other type | MUC1 | Blood | U | 19556244 |
| Other type | TLR4 | Blood | U | 29328946 |
| Other type | caspase 4 | Blood | U | 32581219 |
| Other type | caspase 5 | Blood | U | 29328946 |

^a)^ U represents increased permeability and D decreased permeability.

^b)^ References are shown using PubMed ID (PMID).

**Table S4.** List of literature-derived variants and genes in PD.

| **Genetic variant** | **Corresp. genes** | **Genetic variant** | **Corresp. genes** | **Genetic variant** | **Corresp. genes** | **Genetic variant** | **Corresp. genes** | **Genetic variant** | **Corresp. genes** |
| --- | --- | --- | --- | --- | --- | --- | --- | --- | --- |
| rs823114 | NUCKS1 | rs6721961 | NFE2L2 | rs3130434 | HLA-C | rs193922936 | FMR1 | rs356168 | SNCA |
| rs1721100 | FGF20 | rs9938550 | HSD3B7 | rs3129883 | HLA-DQA1 | rs7525979 | NLRP3 | rs1229984 | ADH1B |
| rs12720208 | FGF20 | rs429358 | APOE | rs7745002 | HLA-DQA1 | rs1051312 | SNAP25 | rs11868035 | SREBF1 |
| rs713598 | TAS2R38 | rs3818361 | CR1 | rs3844313 | HLA-DQA1 | rs3746544 | SNAP25 | rs660895 | HLA-DRB1 |
| rs1726866 | TAS2R38 | rs6265 | BDNF | HLA-DQA1*03:03 | HLA-DQA1 | rs363043 | SNAP25 | rs2230806 | ABCA1 |
| rs10246939 | TAS2R38 | rs2279590 | CLU | rs7744001 | HLA-DQB1 | rs2296481 | DNAJC6 | rs1801474 | PRKN |
| rs7041 | GC | rs8126696 | DYRK1A | rs2856691 | HLA-DQB1 | rs6588142 | DNAJC6 | ACHE_TGTT_DEL | ACHE |
| rs7975232 | VDR | rs9296559 | CD2AP | HLA-DQB1*06 | HLA-DQB1 | rs6588144 | DNAJC6 | rs705379 | PON1 |
| rs33949390 | LRRK2 | rs6356 | TH | rs4947332 | HLA-DRB1 | rs4325172 | DNAJC6 | rs72470544 | HTRA2 |
| rs9614 | FBXO2 | rs387906315 | GBA | rs6457614 | HLA-DRB1 | rs11208644 | DNAJC6 | rs1800547 | MAPT |
| rs6013897 | CYP24A1 | rs76763715 | GBA | rs6910071 | HLA-DRB1 | rs12077111 | DNAJC6 | rs334558 | GSK3B |
| rs10839976 | RIC3 | rs13388259 | downstream of BCYRN1 | rs3817964 | HLA-DRB1 | rs4582839 | DNAJC6 | rs6438552 | GSK3B |
| rs55990541 | RIC3 | SNCA-REP1 | SNCA | rs2736157 | HLA-DRB1 | rs4592284 | DNAJC6 | rs7669 | MTIF3 |
| rs73411617 | RIC3 | SNCA_D4S3474 | SNCA | rs3115572 | HLA-DRB1 | rs4915691 | DNAJC6 | rs9726 | FBXO7 |
| rs11826236 | RIC3 | SNCA_D4S3479 | SNCA | rs2395533 | HLA-DRB1 | rs3818513 | DNAJC6 | rs1695 | GSTP1 |
| rs823128 | NUCKS1 | SNCA_D4S3487 | SNCA | rs2040410 | HLA-DRB1 | rs1014290 | SLC2A9 | rs143561967 | C9orf72 |
| rs1572931 | RAB29 | rs1136666 | GAPDH | rs5000803 | HLA-DRB1 | rs33995883 | LRRK2 | rs2652510 | SLC6A3 |
| rs823156 | SLC41A1 | rs375705174 | NOD2 | rs7769979 | HLA-DRB1 | rs7308720 | LRRK2 | rs11248051 | GAK |
| rs35749011 | GBA;SYT11 | rs7412 | APOE | rs3129299 | HLA-DRB1 | rs3756063 | SNCA | rs3129882 | HLA-DRA |
| rs2230288 | GBA | rs449647 | APOE | rs1143623 | IL1B | rs6280 | DRD3 | rs356165 | SNCA |
| rs2424932 | DNMT3B | rs405509 | APOE | rs529974 | MUL1 | rs1801334 | PRKN | rs2134655 | DRD3 |
| rs998382 | DNMT3B | rs11931074 | SNCA | rs2228570 | VDR | rs12456492 | RIT2 | rs1799836 | MAOB |
| rs2424913 | DNMT3B | rs11564148 | LRRK2 | rs1024611 | CCL2 | rs1800497 | DRD2 | rs66737902 | LRRK2 |
| rs41550117 | POLG | rs356186 | SNCA | rs4680 | COMT | rs5848 | GRN | rs74315359 | PINK1 |
| rs421016 | GBA | rs34778348 | LRRK2 | rs9468199 | ZNF184 | rs3793947 | DLG2 | rs1544410 | VDR |
| rs10797576 | SIPA1L2 | rs356219 | SNCA | rs2241703 | SIRT2 | rs2853826 | MT-ND3 | rs1799964 | TNF |
| rs2414739 | VPS13C | rs4713391 | HLA-C | rs2737024 | SNCA | rs1801133 | MTHFR | GSTT1*0 | GSTT1 |
| rs11158026 | GCH1 | rs7745906 | HLA-C | rs2583959 | SNCA | rs1801131 | MTHFR | rs2736990 | SNCA |
| rs356168 | SNCA | MAPT_del-In9 | MAPT | rs823144 | RAB29 | rs3021088 | MT-ND2 | rs363324 | SLC18A2 |
| rs1229984 | ADH1B | rs2071746 | HMOX1 | rs352140 | TLR9 | rs41456348 | MT-TQ | rs1611115 | DBH |
| rs11868035 | SREBF1;RAI1 | HMOX1_VNTR | HMOX1 | rs1801582 | PRKN | rs897984 | MIR4519 | rs3794087 | SLC1A2 |
| rs660895 | HLA-DRB1 | rs669 | A2M | rs148156462 | COQ2 | rs11651671 | MIR548AT | rs3857059 | SNCA |
| rs2230806 | ABCA1 | rs28357984 | MT-ND2 | rs369704279 | - | rs2645425 | CTSB | rs1130233 | AKT1 |
| rs1801474 | PRKN | rs708723 | RAB29 | rs28358278 | MT-ND3 | rs1128402 | SPPL2B | rs3025039 | VEGFA |
| ACHE_TGTT_DEL | ACHE | rs156429 | GPNMB | rs193302956 | - | rs689466 | PTGS2 | rs356182 | SNCA |
| rs705379 | PON1 | rs4889603 | STX1B | rs4767944 | ALDH2 | rs689465 | PTGS2 | rs1128503 | ABCB1 |
| rs72470544 | HTRA2 | rs140304729 | APP | rs441 | ALDH2 | rs20417 | PTGS2 | rs1805874 | CALB1 |
| rs1800547 | MAPT | rs4698412 | BST1 | rs671 | ALDH2 | rs2338971 | HUSEYO | rs1491942 | LRRK2 |
| rs334558 | GSK3B | rs1564282 | GAK | rs116074753 | GIGYF2 | rs638405 | BACE1 | rs2301135 | SNCA |
| rs6438552 | GSK3B | rs80356769 | GBA | rs34637584 | LRRK2 | rs329648 | MIR4697 | rs1060826 | NOS2 |
| rs7669 | MTIF3 | rs1064651 | GBA | rs387906289 | SMPD1 | rs4898 | TIMP1 | rs2682826 | NOS1 |
| rs9726 | FBXO7 | rs368060 | GBA | rs3822086 | SNCA | rs17576 | MMP9 | rs45478900 | PINK1 |
| rs1695 | GSTP1 | rs1135675 | GBA | rs3775444 | SNCA | rs1469602964 | NFE2L2 | rs2280788 | CCL5 |
| rs143561967 | C9orf72 | rs104886460 | GBA | rs363371 | SLC18A2 | NFE2L2:c.423G>T | NFE2L2 | rs4073 | CXCL8 |
| rs2652510 | SLC6A3 | rs181489 | SNCA | rs11856808 | LINGO1 | rs823118 | RAB29;NUCKS1 | rs1799864 | CCR2 |
| rs4950 | CHRNB3 | rs1388388884 | MMP16 | rs3014864 | PGLYRP4 | rs41347846 | - | rs708730 | SLC41A1 |
| A hyphen (-) indicates that no gene(s) was (or were) specified in the original literature. | | | | | | | | | |

**Table S4.** List of literature-derived variants and genes in PD. (cont.)

| **Genetic variant** | **Corresp. genes** | **Genetic variant** | **Corresp. genes** | **Genetic variant** | **Corresp. genes** | **Genetic variant** | **Corresp. genes** | **Genetic variant** | **Corresp. genes** |
| --- | --- | --- | --- | --- | --- | --- | --- | --- | --- |
| rs11248051 | GAK | rs894278 | SNCA | rs3741918 | GAPDH | rs12637471 | MCCC1 | CCR5_delta32 | CCR5 |
| rs3129882 | HLA-DRA | rs2583988 | SNCA | rs1060619 | GAPDH | rs16856139 | SLC45A3 | rs10794536 | CPLX1 |
| rs356165 | SNCA | rs2619364 | SNCA | rs11240569 | SLC41A1 | rs11240572 | PM20D1 | rs1051308 | HMOX2 |
| rs2134655 | DRD3 | rs10005233 | SNCA | rs120074124 | SMPD1 | rs2435207 | MAPT | rs972936 | IGF1 |
| rs1799836 | MAOB | rs10043 | CHCHD2 | rs75932628 | TREM2 | rs729022 | SYT11 | rs35479735 | NR4A2 |
| rs66737902 | LRRK2 | rs142444896 | CHCHD2 | rs1047735 | NOS1 | rs34372695 | SYT11;RAB25 | rs1467967 | MAPT |
| rs74315359 | PINK1 | rs11558538 | HNMT | rs816353 | NOS1 | rs12118033 | MACF1 | rs242557 | MAPT |
| rs1544410 | VDR | rs1048230 | SLC11A2 | rs3741480 | NOS1 | rs1052553 | MAPT | rs3785883 | MAPT |
| rs1799964 | TNF | rs12817488 | CCDC62;HIP1R | rs1955337 | STK39 | rs1724425 | MAPT | rs2471738 | MAPT |
| GSTT1*0 | GSTT1 | rs6812193 | SCARB2;STBD1;FAM47E | rs900147 | ARNTL | rs8070723 | MAPT | rs7521 | MAPT |
| rs2736990 | SNCA | rs1800562 | HFE | rs2253820 | PER1 | rs17587 | PSMB9 | rs356220 | SNCA |
| rs62063857 | STH;MAPT | rs4025935 | GSTM1 | rs591323 | FGF20 | rs11981883 | SEPTIN14 | rs35652124 | NFE2L2 |
| rs7563724 | GIGYF2 | rs731236 | VDR | rs75548401 | GBA | rs10241628 | SEPTIN14 | rs3851179 | PICALM |
| rs72554080 | GIGYF2 | rs1927914 | TLR4 | rs28359178 | MT-ND5 | rs77231105 | SEPTIN14 | rs652438 | MMP12 |
| rs3775439 | SNCA | rs193303002 | MT-TT | rs767455 | TNFRSF1A | rs193922937 | AFF2 | rs1876487 | SPR |
| rs6532194 | SNCA | rs527236198 | MT-TT | rs4149570 | TNFRSF1A | rs1372525 | SNCA | rs2421095 | SPR |
| rs2270968 | MCCC1 | rs3060174 | DRD2 | rs854560 | PON1 | rs2619363 | SNCA | rs1876491 | SPR |
| rs1384236 | SLC2A13 | rs4934 | SERPINA3 | SNCA:c.-770C>A | SNCA | rs2258689 | MAPT | rs1567229 | SPR |
| rs7315459 | SLC2A13 | rs28363170 | SLC6A3 | SNCA:c.-116C>G | SNCA | rs110402 | CRHR1 | rs2048170 | - |
| rs3135028 | - | rs1799929 | NAT2 | rs3216147 | SNCA | rs1800795 | IL6 | rs4852902 | - |
| rs369730420 | - | rs1799930 | NAT2 | rs1366309839 | NQO2 | rs4986938 | ESR2 | rs2135985 | - |
| rs8093 | GAK | rs1799931 | NAT2 | MAPT:IVS3+9A>G | MAPT | rs283413 | ADH1C | rs1567230 | - |
| rs2306242 | GAK | rs45478696 | DLST | MAPT:IVS11+34G>A | MAPT | rs1018564318 | ADH7 | rs1508060 | - |
| rs1177343555 | GAK | CYP2D6*5 | CYP2D6 | rs3744456 | MAPT | rs356203 | SNCA | rs1396107 | - |
| rs537577117 | STK39 | rs5030655 | CYP2D6 | rs17652121 | MAPT | rs1556423806 | MT-ND3 | rs1561244 | - |
| rs2102808 | STK39 | rs906807 | NDUFV2 | rs11568305 | MAPT | rs1556423597 | MT-ATP6 | rs6730083 | SPR |
| rs2364725 | NFE2L2 | rs6323 | MAOA | rs1169360425 | NOS1 | rs648178 | HIVEP3 | rs1150500 | SPR |
| rs3736282 | CP | MAOA_INTRON_1_VNTR | MAOA | rs1805054 | HTR6 | rs2038978 | HIVEP3 | rs1161265 | SPR |
| rs66508328 | CP | DRD4_EXON_3_VNTR | DRD4 | NR4A2_7048-7049insG | NR4A2 | rs1039997 | HIVEP3 | rs1161267 | SPR |
| rs67870152 | CP | rs662 | PON1 | rs16944 | IL1B | rs661225 | HIVEP3 | rs7684318 | SNCA |
| rs16861642 | CP | rs1041983 | NAT2 | rs1799899 | TF | rs7554964 | HIVEP3 | rs3798097 | SEMA5A |
| rs73020328 | CP | rs1801280 | NAT2 | rs9347683 | PRKN | rs17884724 | PHOX2B | DRD4_dup120bp | DRD4 |
| rs11708215 | CP | rs61018201 | MAOB | rs199951903 | MT-CYB | rs1064792993 | PHOX2B | rs1800955 | DRD4 |
| rs73166855 | CP | rs56092260 | PRKN | NR4A2:c.-291delT | NR4A2 | rs3832799 | ASCL1 | rs905568 | DRD3 |
| rs66953613 | CP | rs35742686 | CYP2D6 | NR4A2:c.-245T>G | NR4A2 | rs682705 | CDCP2 | rs1503670 | DRD3 |
| rs9917256 | STK39 | rs5030732 | UCHL1 | rs2853498 | MT-TL2 | rs7702187 | SEMA5A | rs324026 | DRD3 |
| rs2197120 | SNCA | rs1079597 | DRD2 | rs1599988 | MT-ND1 | ss23418103 |  | rs2234689 | DRD2 |
| rs12603319 | FBXW10 | MAPT_TG | MAPT | rs17571 | CTSD | rs10506151 | LRRK2 | rs1801028 | DRD2 |
| rs393152 | LINC02210 | CCK:c.-45C>T | CCK | rs1008438 | HSPA1A | rs1388598 | LRRK2 | rs1800498 | DRD2 |
| rs1423670702 | MAOB | rs1373006440 | CCK | rs1043618 | HSPA1A | rs1491941 | LRRK2 | rs1799732 | DRD2 |
| rs16947 | CYP2D6 | rs1051740 | EPHX1 | rs56164415 | BDNF | rs1491938 | LRRK2 | rs2255929 | NOS2 |
| rs550610502 | FBXO7 | rs242559 | MAPT | rs1994090 | LRRK2 | rs28358280 | - | rs3899498 | - |
| rs4998386 | GRIN2A | rs7077361 | ITGA8 | rs1630500 | GBA | rs2853493 | - | rs12645693 | BST1 |

A hyphen (-) indicates that no gene(s) was (or were) specified in the original literature.

**Table S4.** List of literature-derived variants and genes in PD. (cont.)

| **Genetic variant** | **Corresp. genes** | **Genetic variant** | **Corresp. genes** | **Genetic variant** | **Corresp. genes** | **Genetic variant** | **Corresp. genes** | **Genetic variant** | **Corresp. genes** |
| --- | --- | --- | --- | --- | --- | --- | --- | --- | --- |
| rs1137070 | MAOA | rs6347 | SLC6A3 | rs2032582 | ABCB1 | rs10784486 | LRRK2 | rs6311 | HTR2A |
| rs761762083 | MAOA | ADH7:c.-94T>C | ADH7 | rs1045642 | ABCB1 | rs1365763 | LRRK2 | rs1801211 | WFS1 |
| rs4646903 | CYP1A1 | ADH7:p.Gly79Ala | ADH7 | rs12718379 | FGF20 | rs17651213 | MAPT | rs28930368 | POMC |
| rs1048943 | CYP1A1 | rs971074 | ADH7 | rs1989756 | FGF20 | rs6438553 | GSK3B | rs2071345 | POMC |
| rs4880 | SOD2 | rs1799752 | ACE | rs1989754 | FGF20 | rs2569190 | CD14 | rs1165228 | USP24 |
| rs287235 | USP24 | rs2494743 | AKT1 | rs1559085 | CAST | rs7094179 | CDNF | rs9991821 | SCARB2 |
| rs487230 | USP24 | rs2498788 | AKT1 | rs1048603 | USP40 | rs2744690 | PRDM2 | rs17234715 | SCARB2 |
| rs555687 | USP24 | rs2494746 | AKT1 | rs1065852 | CYP2D6 | rs2744687 | PRDM2 | rs1130409 | APEX1 |
| rs6671533 | USP24 | rs1130214 | AKT1 | rs10968280 | LINGO2 | rs13306221 | BDNF | rs25487 | XRCC1 |
| rs7512894 | - | rs4919621 | PITX3 | rs1108580 | DBH | rs6821591 | PPARGC1A | rs861539 | XRCC3 |
| rs3738136 | PINK1 | rs1078997 | KANSL1 | rs5320 | DBH | rs2970848 | PPARGC1A | rs1772153 | RAB29 |
| rs2066844 | NOD2 | rs2737029 | SNCA | rs129882 | DBH | rs387906942 | HTRA2 | rs1775151 | RAB29 |
| rs1800630 | TNF | rs356204 | SNCA | RAB29:c.379-12insT | RAB29 | rs2270363 | HMOX2 | IL1RN_VNTR | IL1RN |
| rs1799724 | TNF | rs1549758 | NOS3 | rs2572324 | SNCA | rs224589 | SLC11A2 | rs1800872 | IL10 |
| rs967582 | ELAVL4 | rs3741475 | NOS1 | rs17563965 | MAPT | rs760077 | MTX1 | rs751473385 | PITX3 |
| rs3902720 | ELAVL4 | rs12944039 | NOS2 | rs17649641 | MAPT | rs14235 | BCKDK;STX1B | ATXN8:c.-62G>A | ATXN8 |
| rs3138526 | PARP1 | rs2297516 | NOS2 | rs17650417 | MAPT | rs34594498 | LRRK2 | rs854571 | PON1 |
| rs2793378 | PARP1 | rs6269 | COMT | rs17571857 | MAPT | rs11175964 | LRRK2 | rs797906 | GLIS1 |
| rs2146315 | HIVEP3 | rs4633 | COMT | rs17573266 | MAPT | rs35658131 | LRRK2 | rs10878226 | LRRK2 |
| rs1105414 | HIVEP3 | rs4818 | COMT | rs162036 | MTRR | rs11136000 | CLU | rs11176013 | LRRK2 |
| rs10493099 | HIVEP3 | rs147903261 | - | rs1805087 | MTR | rs1800566 | NQO1 | rs2301134 | SNCA |
| rs783309 | HIVEP3 | rs41528348 | - | rs10380 | MTRR | rs4925 | GSTO1 | rs356221 | SNCA |
| rs1800587 | IL1A | rs28625645 | - | rs17468382 | DCC | rs9268515 | - | rs769931280 | POLG |
| rs9468 | MAPT | rs13312 | USP24 | rs2030737 | EPHB1 | rs356180 | SNCA | rs2307439 | POLG |
| rs439898 | GBA | rs147277743 | ATP13A2 | rs6492998 | CHP1 | rs3775423 | SNCA | rs2302084 | POLG |
| rs3758549 | PITX3 | rs1345514 | EN2 | rs2970332 | RRAS2 | rs356192 | SNCA | rs2246900 | POLG |
| Hp2 | HP | rs1438852 | EN1 | rs735555 | CDK5R1 | rs2737020 | SNCA | rs2066842 | NOD2 |
| rs137853233 | HP | rs2070676 | CYP2E1 | rs7133914 | LRRK2 | rs141051364 | GIGYF2 | rs151107579 | FBXO41 |
| rs760678 | NEDD9 | rs2281983 | PITX3 | rs1946518 | IL18 | rs17690703 | - | rs526106 | FBXO41 |
| rs680 | IGF2 | rs11155313 | PHACTR2 | rs71651683 | ADORA2A | rs17769552 | - | rs61733550 | FBXO41 |
| rs689 | INS | rs7557529 | NFE2L2 | rs5996696 | ADORA2A | rs7215239 | MAPT | rs187317553 | FBXO41 |
| HUMTH01 | TH | rs6706649 | NFE2L2 | rs7033345 | LINGO2 | rs12373139 | - | rs34363861 | FBXO41 |
| rs1721082 | FGF20 | rs2886161 | NFE2L2 | rs4334089 | VDR | rs1635291 | - | rs12233759 | SNCA |
| rs11203822 | FGF20 | rs76708041 | ATG7 | rs10083198 | VDR | rs1981997 | - | rs4698120 | BST1 |
| rs10888125 | FGF20 | rs1402392667 | ATG7 | rs7299460 | VDR | rs6824953 | SCARB2 | rs11724635 | BST1 |
| rs1800629 | TNF | rs2606729 | ATG7 | rs10875708 | VDR | rs4241591 | SCARB2 | rs10878245 | LRRK2 |
| rs2031920 | CYP2E1 | rs577245589 | ATG7 | rs7976091 | VDR | rs6825004 | SCARB2 | rs375570126 | SMPD1 |
| rs2083482 | FIGN | rs4538475 | BST1 | rs1806649 | NFE2L2 | rs3892097 | CYP2D6 | rs9275326 | HLA-DQB1 |
| MT-CO1:c.-7025AluI | - | rs9290751 | MCCC1;LAMP3 | rs11711441 | MCCC1;LAMP3 | rs11012 | LINC02210;PLEKHM1;MAPT | LRPAP1_37bp-I/D | LRPAP1 |
| rs774676466 | SLC6A4 | rs4768212 | LRRK2 | rs10183914 | NFE2L2 | rs3804099 | TLR2 | rs117896735 | INPP5F |
| rs9652490 | LINGO1 | rs2708453 | LRRK2 | rs7311174 | NCAPD2 | rs1801260 | CLOCK | rs76904798 | LRRK2 |
| rs1880753 | - | rs2046932 | LRRK2 | rs2072374 | NCAPD2 | rs1461618417 | GIGYF2 | rs11060180 | CCDC62 |
| rs1880756 | - | rs2306604 | TFAM | rs3751143 | P2RX7 | rs4653767 | ITPKB | rs35303786 | LRRK2 |
| rs33958906 | LRRK2 | rs767770365 | PARK7 | rs11248060 | DGKQ | rs34043159 | IL1R2 | rs17649553 | MAPT |
| rs10878371 | LRRK2 | rs17523802 | PARK7 | rs2001350 | NFE2L2 | rs353116 | SCN3A | rs8118008 | DDRGK1 |
| rs199533 | NSF | rs356198 | SNCA | rs6739381 | 2q31.1 | rs28359172 | - | rs114138760 | GBA;SYT11 |
| rs242562 | MAPT | rs34016896 | NMD3 | rs17577094 | MAPT | rs193302927 | - | rs11931532 | BST1 |
| A hyphen (-) indicates that no gene(s) was (or were) specified in the original literature. | | | | | | | | | |

**Table S4.** List of literature-derived variants and genes in PD. (cont.)

| **Genetic variant** | **Corresp. genes** | **Genetic variant** | **Corresp. genes** | **Genetic variant** | **Corresp. genes** | **Genetic variant** | **Corresp. genes** | **Genetic variant** | **Corresp. genes** |
| --- | --- | --- | --- | --- | --- | --- | --- | --- | --- |
| rs10878405 | LRRK2 | rs3766606 | PARK7 | rs187978668 | ATG5 | rs4073221 | SATB1 | rs77522 | - |
| rs33962975 | LRRK2 | rs7517357 | PARK7 | rs1058808 | ERBB2 | rs12497850 | NCKIPSD | rs356225 | - |
| rs10847864 | HIP1R;CCDC62 | rs2280104 | SORBS3;PDLIM2;C8orf58;BIN3 | GRCh38.p12chr17:34252593G>C | CCL2 | rs143918452 | ALAS1;TLR9;DNAH1;BAP1;PHF7;NISCH;STAB1;ITIH3;ITIH4 | rs1775143 | intergenic region between RAB29 and SLC41A1 |
| rs34884856 | NR4A2 | ATXN2_polyQ | ATXN2 | rs199347 | GPNMB | rs78738012 | ANK2;CAMK2D | rs62637702 | PRKN |
| rs12069239 | FBXO42 | rs2010795 | PDXK | rs4240910 | PIK3CD | rs2694528 | ELOVL7 | rs4130047 | RIT2;SYT4 |
| rs35196193 | FBXO42 | rs7304279 | LRRK2 | rs12502586 | BST1 | rs2740594 | CTSB | rs193302980 | - |
| rs2390669 | STK39 | rs4795541 | SLC6A4 | rs28357980 | - | rs2641116 | PARK7 | rs147029798 | - |
| rs6599389 | GAK | rs10447854 | 7q32 | rs28358279 | - | rs13294100 | SH3GL2 | rs8180209 | - |
| rs3918242 | MMP9 | rs417968 | MAPT | rs28357681 | - | rs10906923 | FAM171A1 | rs1474055 | STK39 |
| rs34311866 | TMEM175;GAK;DGKQ | rs1519686 | HS3ST5;RPSAP43 | rs6430538 | ACMSD;TMEM163 | rs601999 | ATP6V0A1;PSMC3IP;TUBG2 | rs34884217 | TMEM175;GAK;DGKQ |
| rs1880669 | TF | rs202015799 | SYT11;RAB25 | rs733731 | PGLYRP2 | rs11343 | COQ7 | rs7681154 | SNCA |
| rs1049296 | TF | rs6710823 | ACMSD | rs892145 | PGLYRP2 | rs4784227 | TOX3 | rs13201101 | HLA-DQB1 |
| rs1800625 | AGER | rs113961104 | GAK | rs2987763 | PGLYRP3 | rs8005172 | GALC | rs3813135 | PGLYRP2 |
| rs3761863 | LRRK2 | rs75855844 | HLA-DRB5 | rs10888557 | PGLYRP4 | rs28358572 | - | rs199515 | MAPT |
| rs2273311 | FBXO42 | rs2942168 | MAPT | rs12063091 | PGLYRP4 | rs193303025 | - | rs4273468 | BST1 |
| rs2641117 | PARK7 | rs12185268 | MAPT | rs3006440 | PGLYRP4 | rs10513789 | MCCC1;LAMP3 | rs41349444 | MT-RNR2 |
| rs1801394 | MTRR | rs28357975 | - | rs3006448 | PGLYRP4 | rs878870695 | - | rs869096886 | MT-ND4 |
| rs1979277 | SHMT1 | rs1876828 | MAPT | rs3006458 | PGLYRP4 | rs2015062 | - | rs17563986 | MAPT |
| rs169201 | NSF | rs2395163 | HLA region |  |  |  |  |  |  |
| A hyphen (-) indicates that no gene(s) was (or were) specified in the original literature. | | | | | | | | | |

**Table S5.** List of literature-derived variants and genes in CD.

| **Genetic variant** | **Corresp. genes** | **Genetic variant** | **Corresp. genes** | **Genetic variant** | **Corresp. genes** | **Genetic variant** | **Corresp. genes** | **Genetic variant** | **Corresp. genes** |
| --- | --- | --- | --- | --- | --- | --- | --- | --- | --- |
| rs6738825 | PLCL1 | rs2302685 | LRP6 | rs12014762 | MORC4 | rs492602 | FUT2 | rs6822844 | IL2 |
| rs2797685 | PER3;VAMP3 | rs1050152 | SLC22A4 | rs6622126 | MORC4 | rs10063949 | SLC23A1 | rs2221903 | IL21 |
| rs1884613 | HNF4A | rs17622208 | SLC22A5 | rs743776 | IL2RB | rs231775 | CTLA4 | rs17005931 | IL21 |
| rs3863377 | NR1H4 | rs2631367 | SLC22A5 | rs1800470 | TGFB1 | rs1250550 | ZMIZ1 | rs2305764 | MYO9B |
| rs4820425 | RBX1;EP300 | rs2241879 | ATG16L1 | rs11130213 | APEH | rs5743289 | NOD2 | rs11747270 | IRGM |
| rs1487630 | TBC1D1 | rs1250559 | ZMIZ1 | rs12324931 | CYLD | rs10883365 | NKX2-3 | rs180802994 | IRGM |
| rs6478106 | TNFSF15 | rs1250560 | ZMIZ1 | rs12472244 | USP40 | rs11190140 | NKX2-3 | rs9637876 | IRGM |
| rs7329174 | ELF1 | rs181359 | YDJC | rs17314544 | CYLD | rs10045431 | IL12B | rs17313265 | NOD2 |
| rs7765379 | MHC | rs281379 | FUT2;RASIP1 | rs17765311 | USP3 | rs52812045 | CD24 | rs3212227 | IL12B |
| rs9891119 | STAT3 | rs4780355 | SOCS1 | rs2131109 | APEH;BSN | rs1800625 | AGER | rs315952 | IL1RN |
| rs4495224 | PTGER4 | rs2274910 | ITLN1 | rs2302759 | CYLD | rs1800624 | AGER | rs224143 | ZNF365 |
| rs13420683 | ZAP70 | rs2569190 | CD14 | rs4047198 | USP40 | rs2070600 | AGER | rs2823256 | NRIP1;USP25 |
| rs2910164 | MIR146A | rs3764022 | CLEC2D | rs7205423 | CYLD | rs1047781 | FUT2 | rs3749172 | GPR35 |
| rs919766 | IL12B | rs61752717 | MEFV | rs838548 | USP40 | rs4959235 | SLC22A23 | rs514000 | PTPN2 |
| rs2288831 | IL12B | rs28362679 | BTNL2 | rs7234029 | PTPN2 | rs9503518 | SLC22A23 | rs4503083 | IDO2 |
| rs4768261 | MUC19 | rs13015714 | IL1RL1 | rs2981804 | DMBT1 | rs207959 | SLCO3A1 | rs2043211 | CARD8 |
| rs7573065 | SLC11A1 | rs1128503 | ABCB1 | rs1801274 | FCGR2A | rs1523128 | NR1I2 | rs2927488 | BCL3 |
| rs13107325 | SLC39A8 | rs2032582 | ABCB1 | rs1801282 | PPARG | rs1551398 | LOC105375746 | rs3087243 | CTLA4 |
| rs962917 | MYO9B | rs11235604 | ATG16L2 | rs1883832 | CD40 | rs1793004 | NELL1 | rs3093158 | CYP4F2 |
| rs1800872 | IL10 | rs10758669 | JAK2 | rs104895447 | NOD2 | rs1800630 | TNF | rs3806265 | NLRP3 |
| rs1800871 | IL10 | rs744166 | STAT3 | rs2066843 | NOD2 | rs2352975 | TRAIP | rs3814055 | NR1I2 |
| rs3024505 | IL10 | rs4958847 | IRGM | rs2108622 | CYP4F2 | rs17598137 | TRAIP | rs3814057 | NR1I2 |
| rs2073617 | TNFRSF11B | rs1000113 | IRGM | rs2239774 | RAC2 | rs2271960 | TRAIP | rs3819025 | IL17A |
| rs2293152 | STAT3 | rs10954213 | IRF5 | rs2243639 | SFTPD | rs1865741 | USP4 | rs4274855 | TLR10 |
| rs957970 | STAT3 | rs10761659 | ZNF365 | rs4358188 | BPI | rs9881860 | USP4 | rs4283605 | BSN |
| rs16822581 | LY75 | rs1521868 | IGR2196 | rs4988235 | LCT | rs17080505 | USP4 | rs4809330 | ZGPAT |
| rs17860508 | IL12B | rs7517847 | IL23R | rs5743277 | NOD2 | rs9874474 | USP4 | rs4925648 | NLRP3 |
| rs6556412 | IL12B | rs2066845 | NOD2 | rs7574865 | STAT4 | rs1800668 | GPX1 | rs5743836 | TLR9 |
| rs1801198 | TCN2 | rs2066844 | NOD2 | rs34448891 | SLC11A1 | rs1972619 | CARD8 | rs5756564 | RAC2 |
| rs9606756 | TCN2 | rs2066847 | NOD2 | rs3731865 | SLC11A1 | rs2631372 | - | rs6446298 | - |
| rs4663402 | ATG16L1 | rs6871626 | IL12B | rs17235409 | SLC11A1 | rs1992660 | 5p13.1 | rs6559629 | TLE1 |
| rs4663421 | ATG16L1 | rs6887695 | IL12B | rs104895427 | NOD2 | rs1992662 | 5p13.1 | rs6785049 | NR1I2 |
| rs6737398 | ATG16L1 | rs1250569 | ZMIZ1 | rs104895431 | NOD2 | rs2074902 | CYP4F2 | rs7785088 | MAGI2 |
| rs10210302 | ATG16L1 | rs10114470 | TNFSF15 | rs104895444 | NOD2 | rs2907748 | NOD1 | rs8049439 | ATXN2L |
| rs6962966 | MAGI2 | rs7869487 | TNFSF15 | rs104895467 | NOD2 | rs2075818 | NOD1 | rs7745002 | HLA-DQA1 |
| rs10846086 | SLC2A14 | rs6478108 | TNFSF15 | rs1272 | CYP4F2 | rs2075820 | NOD1 | rs2858333 | HLA-DQA1 |
| rs12815313 | SLC2A14 | rs7848647 | TNFSF15 | rs6572 | RAC2 | rs2075822 | NOD1 | rs3129883 | HLA-DQA1 |
| rs2889504 | SLC2A14 | rs3810936 | TNFSF15 | rs272893 | SLC22A4 | rs2076756 | NOD2 | rs3844313 | HLA-DQA1 |
| rs144982232 | MST1 | rs6478109 | TNFSF15 | rs273900 | SLC22A4/5 | rs2160322 | MAGI2 | rs3807306 | IRF5 |
| rs478582 | PTPN2 | rs2476601 | PTPN22 | rs323149 | MAGI2 | rs11770589 | IRF5 | rs7454108 | HLA-DQB1 |
| rs11739135 | IBD5 | rs9501161 | NELFE | rs361525 | TNF | rs2222202 | IL10 | rs2071876 | HLA-DQB1 |
| rs12521868 | C5orf56 | rs1041983 | NAT2 | rs762421 | ICOSLG | rs2276707 | NR1I2 | rs2738786 | RTEL1 |
| rs2201841 | IL23R | rs1799931 | NAT2 | rs917997 | IL18RAP | rs2276886 | CXCL9 | rs2858880 | HLA-DRB1 |
| rs2542151 | PTPN2 | rs10512734 | - | rs991804 | CCL2 | rs2279002 | MYO9B | rs73440307 | GLT1D1 |
| rs2847288 | PTPN2 | rs11242115 | IRF1 | rs12948909 | CAVIN1 | DLG5_e26 | DLG5 | HLA-B*5201 | HLA-B |
| rs2542170 | PTPN2 | rs17166050 | RAD50 | rs3809758 | STAT3 | rs2289310 | DLG5 | rs72945092 | QRSL1 |
| rs484020 | PTPN2 | rs2067819 | PPARG | rs1026916 | STAT3 | rs2297322 | SLC15A1 | rs28362684 | BTNL2 |
| rs487273 | PTPN2 | rs2235035 | ABCB1 | rs1128535 | TRAIP | rs2298428 | UBE2L3 | HLA-Bw4 | HLA-B |
| rs11875687 | PTPN2 | rs1922242 | ABCB1 | rs1248696 | DLG5 | rs2305767 | MYO9B | 3DL1 | KIR3DL1 |
| rs774676466 | SLC6A4 | rs280519 | TYK2 | rs1042522 | TP53 | rs80244186 | AKAP11 | rs2975788 | GPR35 |
| A hyphen (-) indicates that no gene(s) was (or were) specified in the original literature. | | | | | | | | | |

**Table S5.** List of literature-derived variants and genes in CD. (cont.)

|  | **Genetic variant** | **Corresp. genes** | **Genetic variant** | **Corresp. genes** | **Genetic variant** | **Corresp. genes** | **Genetic variant** | **Corresp. genes** | **Genetic variant** | **Corresp. genes** |
| --- | --- | --- | --- | --- | --- | --- | --- | --- | --- | --- |
|  | rs1893217 | PTPN2 | rs2288877 | CARD8 | rs1363758 | NLRP11 | rs2306801 | KCNN4 | 2DS3 | KIR2DS3 |
|  | rs10065172 | IRGM | rs324015 | STAT6 | rs1373692 | PTGER4 | rs2297441 | TNFRSF6B | rs9607431 | RAC2 |
|  | rs1800896 | IL10 | rs3892175 | PPARG | rs1457092 | MYO9B | rs2315008 | ZGPAT | rs9858542 | BSN |
|  | rs2066842 | NOD2 | rs4778889 | IL16 | rs1523127 | NR1I2 | rs2872507 | ORMDL3 | rs9935563 | CDH1 |
|  | rs1728918 | UCN | rs4796793 | STAT3 | rs12721602 | NR1I2 | rs504963 | FUT2 | rs10734105 | TCERG1L |
|  | rs61300271 | ELF1 | rs5498 | ICAM1 | rs12721607 | NR1I2 | rs4077515 | CARD9;SNAPC4 | rs11229030 | SLC43A3;PRG2;AIFM2 |
|  | rs241427 | TAP2 | rs4986790 | TLR4 | rs13246026 | MAGI2 | rs12720356 | - | rs102275 | FADS1 |
|  | rs140068907 | HLA-DQB1;HLA-DQA2 | rs9271060 | HLA-DRB1;HLA-DQA1 | rs12677663 | C8orf84;TERF1;RPL7;RDH10;KCNB2 | rs6837335 | TXK;TEC;SLAIN2;SLC10A4;ZAR1;FRYL | rs13003464 | PUS10;PEX13;REL;KIAA1841;C2orf74;PAPOLG;USP34 |
|  | rs3764147 | LACC1;C13orf31 | rs1545620 | MYO9B | rs17234657 | PTGER4 | rs657555 | PTPN2 | rs864745 | JAZF1;CREB5 |
|  | rs3951715 | NUR77 | rs4979462 | TNFSF15 | rs35873774 | XBP1 | rs359457 | CPEB4 | rs2945412 | WSB1;LOC440419;KSR1;LGALS9;NOS2 |
|  | rs10489629 | IL23R | rs6074022 | CD40;MMP9 | rs60872763 | CTLA4 | rs415890 | CCR6 | rs3897478 | ADAM30 |
|  | rs11209032 | IL23R | rs6503695 | STAT3;STAT5B;STAT5A | rs2006996 | TNFSF15 | rs151181 | IL27;SH2B1;EIF3C;LAT;CD19 | rs694739 | ESRRA;PRDX5 |
|  | rs11465804 | IL23R | rs224090 | - | rs2542152 | PTPN2 | rs713875 | MTMR3 | rs7015630 | RIPK2 |
|  | rs1343151 | IL23R | rs13300483 | TNFSF15;TNFSF8 | rs3738447 | NLRP3 | rs736289 | - | rs10065637 | IL31RA;IL6ST;ANKRD55 |
|  | rs1495965 | IL23R | rs3197999 | MST1R | rs6841698 | TLR10 | rs740495 | GPX4;SBNO2 | rs12663356 | - |
|  | rs2201840 | IL23R | rs12994997 |  | rs7658893 | TLR10 | rs780093 | GCKR | rs13204742 | - |
|  | rs4833095 | TLR1 | rs6740462 | SPRED2 | rs8050910 | FAM92B | rs1819658 | UBE2D1 | rs16967103 | SPRED1;RASGRP1 |
|  | rs4343 | ACE | rs11195128 | SMNDC1;DUSP5 | rs16853571 | PHOX2B | rs1998598 | DENND1B | rs17695092 | CPEB4 |
|  | rs73243351 | TBC1D1 | rs4986791 | TLR4 | rs2301436 | CCR6;FGFR10P;RNASE2 | rs7076156 | ADO;ZNF365;ERG2 | rs6856616 | TBC1D1;KLF3 |
|  | rs394522 | CCR6 | rs224136 | ZNF365 | rs2836754 | - | rs2549794 | ERAP2;LRAP | rs10865331 | - |
|  | rs2062305 | TNFSF11 | rs10024216 | TLR10 | rs10077785 | IBD5 | rs3091315 | - | rs17391694 | - |
|  | rs7720838 | PTGER4 | rs10431923 | CDH1 | rs10801047 | - | rs4871611 | - | rs4625 | DAG1 |
| rs1045642 | | ABCB1 | rs10521209 | NOD2 | rs17221417 | NOD2 | rs4902642 | ZFP36L1 | rs602662 | FUT2 |
| rs10181042 | | C2orf74;REL | rs55646866 | NLRP3 | rs2064689 | IL23R | rs6568421 | PRDM1 | rs755374 | IL12B |
| rs1736020 | | - | rs4266924 | NLRP3 | rs9988642 | IL23R | rs7423615 | SP140 | rs7725052 | PTGER4 |
| rs2838519 | | ICOSLG | rs4353135 | NLRP3 | rs10889676 | IL23R | rs7714584 | IRGM | rs7731626 | ANKRD55 |
| rs4409764 | | NKX2-3 | rs10733113 | NLRP3 | rs11465802 | IL23R | rs7927997 | EMSY | rs1250563 | ZMIZ1 |
| rs4263839 | | TNFSF15 | rs3936503 | CCNY | rs2902440 | IL23R | rs11167764 | NDFIP1 | rs1332099 | NKX2-3 |
| rs1024611 | | CCL2 | rs10870077 | CARD9 | rs6669582 | IL23R | rs11564258 | MUC19;LRRK2 | rs2807264 | CD40LG |
| rs72553867 | | IRGM | rs10925019 | NLRP3 | rs10789230 | IL23R | rs11871801 | STAT3;MLX | rs2836882 | PSMG1 |
| rs13361189 | | IRGM | rs10975003 | JAK2 | rs11209002 | IL23R | rs12242110 | CREM | rs10822050 | ZNF365 |
| rs76418789 | | IL23R | rs10995271 | ZNF365 | rs11209003 | IL23R | rs12722489 | IL2RA | rs11145763 | CARD9 |
| rs1799724 | | TNF | rs11175593 | LRRK2 | rs12567232 | IL23R | rs13073817 | - | rs11580078 | IL23R |
| rs2241880 | | ATG16L1 | rs11465788 | IL23R | rs1456893 | - | rs13428812 | DNMT3A | rs11741255 | IL5 |
| rs1004819 | | IL23R | rs11571297 | CTLA4 | rs1736135 | - | rs17309827 | - | rs12598357 | SBK1 |
| rs10889677 | | IL23R | rs11571302 | CTLA4 | rs3828309 | ATG16L1 | rs3792109 | ATG16L1 | rs17466626 | LRRK2 |
| rs11209026 | | IL23R | rs11716445 | RHOA | rs114985235 | HLA-B;HLA-C | rs10486483 | C7orf71;SKAP2 | rs36001488 | ATG16L1 |
| rs851139 | | TLR5 | rs11938795 | IL2 | rs117372389 | NOD2 | rs187238 | IL18 | rs72743477 | SMAD3 |
| rs5744174 | | TLR5 | rs12035082 | - | rs7927894 | EMSY | rs7705924 | SLCO6A1 | rs77150043 | ADCY7 |
| rs2076753 | | NOD2 | rs104895469 | NOD2 | rs485186 | FUT2 | rs17525495 | LTA4H | rs2228145 | IL6R |
| rs3761547 | | FOXP3 | rs34637584 | LRRK2 | rs676388 | FUT2 | rs2236379 | PRKCQ | rs33995883 | LRRK2 |
| rs2232365 | | FOXP3 | rs2069717 | IFNG | rs28999107 | - | rs767455 | TNFRSF1A | rs104895438 | NOD2 |
| rs2294021 | | FOXP3 | rs2228570 | VDR | rs10884966 | - | rs1061624 | TNFRSF1B | rs104895421 | NOD2 |
| rs1583792 | | - | rs6808936 | - | rs11683692 | - | rs3397 | TNFRSF1B | rs104895423 | NOD2 |
|  | A hyphen (-) indicates that no gene(s) was (or were) specified in the original literature. | | | | | | | | | |

**Table S5.** List of literature-derived variants and genes in CD. (cont.)

|  | **Genetic variant** | **Corresp. genes** | **Genetic variant** | **Corresp. genes** | **Genetic variant** | **Corresp. genes** | **Genetic variant** | **Corresp. genes** | **Genetic variant** | **Corresp. genes** |
| --- | --- | --- | --- | --- | --- | --- | --- | --- | --- | --- |
|  | rs2241097 | TLR5 | rs11805303 | IL23R | rs4613763 | PTGER4 | rs12597188 | CDH1 | rs62324212 | IL21 |
|  | rs2675670 | - | rs4343432 | - | rs17582416 | - | rs17293632 | SMAD3 | rs4821544 | NCF4 |
|  | rs780204985 | NOD2 | rs3213119 | IL12B | rs763780 | IL17F | rs2850407 | DKK2 | rs1799946 | DEFB1 |
|  | rs755127265 | NOD2 | rs61729946 | NICN1 | rs3939286 | IL33 | rs1462278 | TNS3;ADCY1 | rs3785142 | CYLD |
|  | rs140716236 | NOD2 | rs200140527 | ARIH2 | rs6716753 | SP110;SP140 | rs153109 | IL27 | rs4656940 | CD244;ITLN1 |
|  | rs199475913 | NOD2 | rs138274580 | ATG4B | rs701109 | MME | rs6596 | SNX20 | rs139518863 | SLC2A13 |
|  | rs758485603 | NOD2 | rs7549308 | CHTOP | rs7308720 | LRRK2 | rs212388 | TAGAP | rs2284553 | IFNGR2 |
|  | rs770915641 | NOD2 | rs9264942 | - | rs141326733 | HEATR3 | rs6651252 | - | rs7170683 | LRRK1 |
|  | rs9286879 | FASLG;TNFSF18 | rs3091316 | CCL2;CCL7;CCL11 | rs17098094 | MIR4681;ACUL1 | rs2581828 | RFT1;RP11-894J14.5 | rs7279062 | KRTAP21-3;KRTAP25-1 |
|  | rs1251391668 | NOD2 | rs17855750 | IL27 | rs1946518 | IL18 | rs10492862 | CDH13 | rs10119910 | IFNA10 |
|  | rs202070332 | MDGA1 | rs181206 | IL27 | rs41310915 | PGLYRP4 | rs34687326 | SLAMF8 | rs113055208 | IFNA4 |
|  | rs1433620891 | ZNF366 | rs2304256 | TYK2 | rs360718 | IL18 | rs56116661 | LPP | STIN2-VNTR | SLC6A4 |
|  | rs488200 | RAP1A | rs1051992 | CAVIN3 | rs1800795 | IL6 | rs6908425 | CDKAL1 | rs11681525 | - |
| rs184950714 | | HLA-DQB1 | rs3838646 | CD24 | rs2234663 | IL1RN | rs146313066 | NOD2 | rs7746082 | - |
| rs56167332 | | IL12B | rs1748195 | USP1 | rs5744088 | TLR8 | rs1456896 | SPATA48;ZPBP | rs1800629 | TNF |
| rs2188958 | | - | rs724016 | - | rs2407992 | TLR8 | rs1847472 | BACH2 | rs9292777 | PTGER4 |
| rs12413565 | | NKX2-3 | rs9319943 | - | rs5744067 | TLR8 | rs7517810 | - | rs4796647 | STAT3 |
| rs382412 | | ZNF365 | rs3761548 | FOXP3 | rs7282490 | ICOSLG | rs8005161 | GALC;GPR65 | rs601338 | FUT2 |
| rs12786216 | | MIR4299;MRVI1-AS1 | rs7954567 | CD27;TNFRSF1A;LTBR | rs1292053 | TUBD1;RPS6KB1 | rs10495903 | THADA;ZFP36L2 | rs1142287 | SCAMP3;MUC1 |
| rs4366152 | | TNFSF15;TNFSF8 | rs9525625 | AKAP1;TFSF11 | rs12942547 | STAT3;STAT5B;STAT5A | rs4246905 | TNFSF8;TNFSF15;TNC | rs5761958 | MIR3199-2;MIR548J |
| rs9491697 | | RSPO3 | rs9459874 | CCR6 | rs13333062 | SALL1;ADCY7 | rs1250546 | - | rs11742570 | - |
| rs2187668 | | HLA-DQA1 | HLA-DRB1#37 | HLA-DRB1 | HLA-Cw*1202 | HLA-C | rs1906493 | - | rs11574514 | PSMB10 |
| rs3794433 | | LACC1;CCDC122 | rs12642902 | Intergenic (left IL21) | rs6852535 | Intergenic (left IL21) | rs7554511 | INAVA;KIF21B;CACNA1S | rs2069762 | Upstream IL2 (right side) |
| rs13126505 | | NFKB1;SLC39A8;BANK1 | rs2413583 | TAB1;SYNGR1;RPL3 | rs2058813 | SNX20;NOD2;CYLD;NKD1 | rs4807569 | STK11;SBNO2;729119 | rs2024092 | GPX4;SBNO2;STK11 |
| rs78898421 | | TNFSF15;TNFSF8 | rs11584383 | CACNA1S;KIF21B | rs2427870 | CD40LG;ARHGEF6 | rs1819333 | CCR6;RPS6KA2;RNASET2 | HLA-DQB1*02:02 | HLA-DQB1 |
| HLA-DPB1*0901 | | HLA-DPB1 | HLA-DRB1*0405 | HLA-DRB1 | rs12924003 | SNX20;NOD2;CYLD;NKD1 | rs11235667 | STARD10;ATG16L2;FCHSD2 | rs2072711 | NCF4;CSF2RB |
| rs7608910 | | REL;C2orf74;KIAA1841;AHSA2P | rs3091338 | IL3;ACSL6;P4HA2;PDLIM4;SLC22A4 | rs516246 | PUS10;PEX13;REL;KIAA1841;C2orf74;PAPOLG;USP34 | rs9258260 | UBD;HLA-A;HLA-G;HLA-F;MOG;GABBR1;HLA-H | rs6545946 | TMEM17;EHBP1;CPAMD8;AK3 |
| rs603439 | | TRIP4;SMAD6;SMAD3;MAP2K1;IGDCC4;IGDCC3 | rs2058660 | IL12RL2;IL18R1;IL1RL1;IL18RAP | rs727563 | TEF;NHP2L1;PMM1;L3MBTL2;CHADL | rs1799964 | TNF;LTA;HLA-DQA2;LST1;LTB | rs7702331 | FCHO2;TMEM171;TMEM174;FOXD1 |
| rs11190141 | | NKX2-3;SLC25A28;GOT1;ENTPD7;CNNM1;COX15;CUTC | rs9271366 | HLA-DRB5;HLA-DQA1;HLA-DRB1;HLA-DRA;BTNL2 | rs6927022 | HLA-DQB1;HLA-DRB1;HLA-DQA1;HLA-DRA | rs57275892 | BAZ1A;SRP54;FAM177A1;PPP2R3C;KIAA0391;PSMA6;NFKBIA | rs3731257 | CDKN2A;CDKN2A-DT;CDKN2B;CDKN2B-AS1 |
| rs10947261 | | HLA-DRB5;HLA-DQA1;HLA-DRB1;HLA-DRA;BTNL2 | rs1260326 | SNX17;ZNF513;KRTCAP3;FNDC4;PPM1G;IFT172;GCKR;NRBP1 | rs6679677 | MAGI3;PHTF1;RSBN1;PTPN22;BCL2L15;AP4B1;DCLRE1B;HIPK1;OLFML3 | rs4802307 | IGFL3;IGFL2;DKFZp434J0226;IGFL1;HIF3A;PPP5C;CCDC8;PNMAL1;PNMAL2 | rs2188962 | LOC441108;IL3;CSF2;HINT1;LYRM7;SLC22A4;C5orf56;IRF1;CDC42SE2;RAPGEF6;ACSL6;P4HA2;PDLIM4;SLC22A5;CTC-432M15.3;FNIP1 |
| rs2149085 | | RNASET2;FGFR1OP;CCR6;MIR3939 | rs7775228 | HLA-DQB1;HLA-DQA2 |  |  |  |  |  |  |
|  | A hyphen (-) indicates that no gene(s) was (or were) specified in the original literature. | | | | | | | | | |

| **Gene name** | **Gene type** | **Genes in Table S1?**  (Y / N) | **Literature evidence**  (PMID) | **AWmeta-blood**  (U / D / NS / NA) | **AWmeta-SN**  (U / D / NS / NA) |
| --- | --- | --- | --- | --- | --- |
| ADORA2A-AS1 | antisense lncRNA | N | NA | U \| NS | U |
| ATP6V0A1 | protein coding | Y | - | U | D |
| AUP1 | protein coding | N | 9806835 | D | U |
| BCL7C | protein coding | N | 31898332 | D | U |
| BDNF-AS | antisense lncRNA | N | 32057951 | D \| NS | U |
| BTNL2 | protein coding | N | NA | U \| NS | D \| NS |
| CASC16 | lincRNA | N | NA | U \| NS | D \| NS |
| CDIP1 | protein coding | N | NA | U \| NS | D \| NS |
| DBH-AS1 | antisense lncRNA | N | NA | D \| NS | D |
| FAM171A2 | protein coding | N | 33087363 | D \| NS | U \| NS |
| FAM47E-STBD1 | protein coding | N | 30957308 | D \| NS | D |
| FASTKD3 | protein coding | N | NA | U | D |
| FMR1-AS1 | bidirectional lncRNA | N | NA | U \| NS | D \| NS |
| GBF1 | protein coding | N | 32652860 | D | D |
| GTF3A | protein coding | N | NA | U \| NS | U |
| HDAC2-AS2 | antisense lncRNA | N | NA | U \| NS | D \| NS |
| HLA-DQB1-AS1 | antisense lncRNA | N | NA | U \| NS | D \| NS |
| HLA-DQB2 | protein coding | N | NA | U \| NS | U \| NS |
| HSPA1L | protein coding | N | 24887138; 24270810 | U | U |
| IL19 | protein coding | N | NA | U \| NS | D \| NS |
| INS-IGF2 | protein coding | N | NA | U \| NS | D \| NS |
| IP6K2 | protein coding | N | NA | U \| NS | U \| NS |
| ITGA8 | protein coding | Y | - | U \| NS | U \| NS |
| ITIH1 | protein coding | N | NA | D \| NS | U \| NS |
| KANSL1 | protein coding | Y | - | D \| NS | U \| NS |
| KLC1 | protein coding | N | 29991596; 24073418 | U | D |
| KLRG1 | protein coding | N | NA | D \| NS | U |
| LINC00243 | lincRNA | N | NA | U \| NS | NA |
| LINC02210-CRHR1 | protein coding | Y | - | D \| NS | D \| NS |
| LINC02456 | intronic lncRNA | N | NA | NA | D \| NS |
| LOC100287329 | antisense lncRNA | N | NA | NA | NA |
| LOC101927942 | lincRNA | N | NA | NA | NA |
| LOC105369501 | antisense lncRNA | N | NA | NA | NA |
| LOC105369736 | antisense lncRNA | N | NA | NA | NA |
| LOC105370032 | antisense lncRNA | N | NA | NA | NA |
| LOC105371114 | antisense lncRNA | N | NA | NA | NA |
| LOC105371702 | bidirectional lncRNA | N | NA | NA | NA |
| LOC105371449 | lincRNA | N | NA | NA | NA |
| SLC44A4 | protein coding | N | NA | U \| NS | U \| NS |
| SNCA-AS1 | antisense lncRNA | N | 24026176 | D \| NS | D \| NS |
| SPECC1L-ADORA2A | sense lncRNA | N | NA | D \| NS | U |
| SPPL2C | protein coding | Y | - | U \| NS | U \| NS |
| SYN1 | protein coding | N | 26269422 | U \| NS | D |

**Table S6.** Detailed information of dbSNP-predicted genes in PD, evidenced by literature and/or AWmeta.

Y stands for yes, N for no, PMID for PubMed ID, and NA for not available. “-” represents not applicable. AWmeta-blood and AWmeta-SN denote the AWmeta-derived gene differential quantification results in PD peripheral blood and substantia nigra, respectively. U refers to upregulated gene expression, D indicates downregulated gene expression, and NS represents “not statistically significant” (with the DEG threshold set as |log_2_FC| > log_2_1.5 and *P* < 0.05).

| **Gene name** | **Gene type** | **Genes in Table S1?**  (Y / N) | **Literature evidence**  (PMID) | **AWmeta-blood**  (U / D / NS / NA) | **AWmeta-SN**  (U / D / NS / NA) |
| --- | --- | --- | --- | --- | --- |
| LOC105371702 | bidirectional lncRNA | N | NA | NA | NA |
| LOC105371720 | lincRNA | N | NA | NA | NA |
| LOC105371744 | bidirectional lncRNA | N | NA | NA | NA |
| LOC105371800 | intronic lncRNA | N | NA | NA | NA |
| LOC105374339 | intronic lncRNA | N | NA | NA | NA |
| SLC2A9-AS1 | intronic lncRNA | N | NA | D \| NS | D \| NS |
| LOC105374925 | antisense lncRNA | N | NA | NA | NA |
| LOC105375056 | antisense lncRNA | N | 30538480 | NA | NA |
| LOC105375630 | bidirectional lncRNA | N | NA | NA | NA |
| LOC105377286 | intronic lncRNA | N | NA | NA | NA |
| LOC105377329 | lincRNA | N | NA | NA | NA |
| LOC105377374 | lincRNA | N | NA | NA | NA |
| LOC105379297 | lincRNA | N | NA | NA | NA |
| LOC105379396 | antisense lncRNA | N | NA | NA | NA |
| LOC107984284 | bidirectional lncRNA | N | NA | NA | NA |
| LOC107984782 | lincRNA | N | NA | NA | NA |
| LOC107985250 | lincRNA | N | NA | NA | NA |
| LOC107985948 | intronic lncRNA | N | NA | NA | NA |
| LOC107985959 | bidirectional lncRNA | Y | - | NA | NA |
| LOC107986115 | antisense lncRNA | N | NA | NA | NA |
| LOC107986119 | lincRNA | N | NA | NA | NA |
| HFE-AS1 | antisense lncRNA | N | NA | NA | NA |
| LOC112268089 | antisense lncRNA | N | NA | NA | NA |
| LOC112268095 | intronic lncRNA | N | NA | NA | NA |
| LOC339862 | lincRNA | N | NA | U \| NS | U \| NS |
| IL6-AS1 | antisense lncRNA | N | NA | D \| NS | D |
| LRRC37A2 | protein coding | Y | - | U \| NS | U |
| LTA | protein coding | N | 29323027; 9448320 | D \| NS | D \| NS |
| MAPT-AS1 | antisense lncRNA | Y | - | D \| NS | D \| NS |
| MAPT-IT1 | intronic lncRNA | N | NA | U \| NS | D \| NS |
| MIR4761 | miRNA | N | NA | NA | NA |
| MIR4769 | miRNA | N | NA | NA | NA |
| MIR6832 | miRNA | N | NA | NA | NA |
| MIR762HG | miRNA | N | NA | D \| NS | D \| NS |
| MT-ATP6 | protein coding | Y | - | U | D \| NS |
| MT-ATP8 | protein coding | Y | - | U | U \| NS |
| MT-CO1 | protein coding | Y | - | D | U \| NS |
| MT-CO2 | protein coding | Y | - | D \| NS | U \| NS |

**Table S6.** Detailed information of dbSNP-predicted genes in PD, evidenced by literature and/or AWmeta. (cont.)

Y stands for yes, N for no, PMID for PubMed ID, and NA for not available. “-” represents not applicable. AWmeta-blood and AWmeta-SN denote the AWmeta-derived gene differential quantification results in PD peripheral blood and substantia nigra, respectively. U refers to upregulated gene expression, D indicates downregulated gene expression, and NS represents “not statistically significant” (with the DEG threshold set as |log_2_FC| > log_2_1.5 and *P* < 0.05).

| **Gene name** | **Gene type** | **Genes in Table S1?**  (Y / N) | **Literature evidence**  (PMID) | **AWmeta-blood**  (U / D / NS / NA) | **AWmeta-SN**  (U / D / NS / NA) |
| --- | --- | --- | --- | --- | --- |
| MT-CO3 | protein coding | Y | - | U | D \| NS |
| MT-CYB | protein coding | Y | - | D | D \| NS |
| MT-ND1 | protein coding | Y | - | D | U \| NS |
| MT-ND2 | protein coding | Y | - | D | U \| NS |
| MT-ND3 | protein coding | Y | - | D | U |
| MT-ND4 | protein coding | Y | - | D | U \| NS |
| MT-ND4L | protein coding | Y | - | D | D \| NS |
| MT-ND5 | protein coding | Y | - | D \| NS | D \| NS |
| MT-ND6 | protein coding | Y | - | U | U \| NS |
| NDUFAF2 | protein coding | N | NA | U | D \| NS |
| NMRAL1 | protein coding | N | NA | U \| NS | U \| NS |
| NQO2-AS1 | antisense lncRNA | N | NA | U | D \| NS |
| PACERR | bidirectional lncRNA | N | NA | U \| NS | U \| NS |
| PACRG | protein coding | N | 19196541; 17590346 | D \| NS | D |
| PAH | protein coding | N | 30337205 | U \| NS | U \| NS |
| PBX2 | protein coding | N | NA | D \| NS | U \| NS |
| PINK1-AS | antisense lncRNA | N | 17362513 | D | D \| NS |
| PRRC2A | protein coding | N | NA | D | U |
| RINL | protein coding | N | NA | U | U \| NS |
| STH | protein coding | Y | - | D \| NS | U |
| SYT17 | protein coding | N | NA | D \| NS | D |
| THBS3 | protein coding | N | NA | U | U |
| TMCO6 | protein coding | N | NA | D \| NS | D |
| TMEM165 | protein coding | N | NA | U | U |
| TSBP1 | protein coding | N | 32827573; 32990896 | U \| NS | U |
| TSBP1-AS1 | antisense lncRNA | Y | - | D \| NS | D |
| TXNL4B | protein coding | N | NA | D \| NS | U \| NS |
| UCHL1-AS1 | bidirectional lncRNA | N | 23064229; 26029048 | NA | D \| NS |
| LOC112268294 | antisense lncRNA | N | NA | NA | NA |

**Table S6.** Detailed information of dbSNP-predicted genes in PD, evidenced by literature and/or AWmeta. (cont.)

Y stands for yes, N for no, PMID for PubMed ID, and NA for not available. “-” represents not applicable. AWmeta-blood and AWmeta-SN denote the AWmeta-derived gene differential quantification results in PD peripheral blood and substantia nigra, respectively. U refers to upregulated gene expression, D indicates downregulated gene expression, and NS represents “not statistically significant” (with the DEG threshold set as |log_2_FC| > log_2_1.5 and *P* < 0.05).

| **Gene name** | **Gene type** | **Genes in Table S2?**  (Y / N) | **Literature evidence**  (PMID) | **AWmeta-blood**  (U / D / NS / NA) | **AWmeta-IM**  (U / D / NS / NA) | **AWmeta-CM**  (U / D / NS / NA) |
| --- | --- | --- | --- | --- | --- | --- |
| ERAP1 | protein coding | N | 28651467; 28651467; 21102463 | U | D \| NS | D \| NS |
| COLEC10 | protein coding | N | NA | U \| NS | U | U |
| TLR8-AS1 | antisense lncRNA | N | NA | U \| NS | U \| NS | U \| NS |
| ICAM4 | protein coding | N | NA | D | U | U |
| LOC112267968 | protein coding | N | NA | D \| NS | U | NA |
| LTA | protein coding | Y | - | U \| NS | D | U \| NS |
| LY75-CD302 | protein coding | N | 27965521; 25557950; 18497880; 26732675; 25557950 | U | U | D |
| MAMSTR | protein coding | Y | - | D | D | D \| NS |
| PBX2 | protein coding | N | 11556964 | U | U | U |
| PSMG4 | protein coding | N | NA | D | D | D |
| RTEL1 | protein coding | Y | - | D | U | U |
| RTN4IP1 | protein coding | N | NA | D \| NS | U | U \| NS |
| TMCO6 | protein coding | N | NA | D \| NS | D \| NS | D |
| HLA-DRA | protein coding | Y | - | D \| NS | U \| NS | U |
| HLA-DQB1 | protein coding | Y | - | D \| NS | U | U |
| BRD2 | protein coding | N | 29293112; 28008999 | U \| NS | D \| NS | U |
| AP4B1-AS1 | antisense lncRNA | N | NA | D \| NS | D \| NS | U \| NS |
| CCDC148-AS1 | antisense lncRNA | N | NA | U \| NS | U | D \| NS |
| IL21-AS1 | antisense lncRNA | Y | - | U | D | NA |
| LINC00491 | lincRNA | N | NA | U \| NS | U \| NS | D \| NS |
| LINC00492 | lincRNA | N | NA | NA | D \| NS | NA |
| LINC01475 | lincRNA | Y | - | U \| NS | U | NA |
| LOC100287329 | antisense lncRNA | N | NA | NA | NA | NA |
| LOC101927825 | antisense lncRNA | N | NA | U \| NS | D \| NS | NA |
| TSBP1-AS1 | antisense lncRNA | N | NA | NA | D \| NS | D \| NS |
| LOC105369736 | antisense lncRNA | N | NA | NA | NA | NA |
| LOC105371082 | antisense lncRNA | N | NA | NA | NA | NA |
| LOC105371251 | antisense lncRNA | N | NA | NA | NA | NA |
| LOC105371720 | lincRNA | N | NA | NA | NA | NA |
| LOC105372272 | antisense lncRNA | N | NA | NA | NA | NA |
| LOC105372629 | antisense lncRNA | N | NA | NA | NA | NA |
| LOC105372988 | antisense lncRNA | N | NA | U \| NS | D \| NS | NA |
| LOC105374764 | antisense lncRNA | N | NA | NA | NA | NA |
| LOC105375024 | antisense lncRNA | N | NA | NA | NA | NA |
| LOC105377989 | lincRNA | N | NA | NA | NA | NA |
| TAGAP-AS1 | antisense lncRNA | N | NA | U \| NS | U | NA |
| LOC105378204 | lincRNA | N | NA | NA | NA | NA |
| LOC105378327 | lincRNA | N | NA | NA | NA | NA |
| LOC105447645 | antisense lncRNA | N | NA | NA | NA | NA |
| LOC107984576 | lincRNA | N | NA | NA | NA | NA |
| LOC107986469 | antisense lncRNA | N | NA | NA | NA | NA |
| LOC107986589 | antisense lncRNA | N | NA | NA | NA | NA |
| IL6-AS1 | antisense lncRNA | N | NA | D \| NS | U \| NS | U |

**Table S7.** Detailed information of dbSNP-predicted genes in CD, evidenced by literature and/or AWmeta.

Y stands for yes, N for no, PMID for PubMed ID, and NA for not available. “-” denotes not applicable. AWmeta-blood, AWmeta-IM and AWmeta-CM refer to the AWmeta-derived gene differential quantification results in CD peripheral blood, ileal and colonic mucosa, respectively. U represents upregulated gene expression, D denotes downregulated gene expression, and NS indicates “not statistically significant” (with the DEG threshold set as |log_2_FC| > log_2_1.5 and *P* < 0.05).

| **Gene name** | **Gene type** | **Genes in Table S2?**  (Y / N) | **Literature evidence**  (PMID) | **AWmeta-blood**  (U / D / NS / NA) | **AWmeta-IM**  (U / D / NS / NA) | **AWmeta-CM**  (U / D / NS / NA) |
| --- | --- | --- | --- | --- | --- | --- |
| LOC285626 | lincRNA | Y | - | D \| NS | D | D \| NS |
| MIR1236 | miRNA | N | NA | D \| NS | D | D \| NS |
| MIR3142HG | miRNA | N | NA | D | U | U |
| MIR3936HG | miRNA | Y | - | D \| NS | D | D |
| NCF4-AS1 | antisense lncRNA | N | NA | U | U \| NS | NA |
| RTEL1-TNFRSF6B | sense lncRNA | N | NA | D | U | U |
| SCARNA5 | intronic lncRNA | N | NA | U \| NS | D \| NS | U |

**Table S7.** Detailed information of dbSNP-predicted genes in CD, evidenced by literature and/or AWmeta. (cont.)

Y stands for yes, N for no, PMID for PubMed ID, and NA for not available. “-” denotes not applicable. AWmeta-blood, AWmeta-IM and AWmeta-CM refer to the AWmeta-derived gene differential quantification results in CD peripheral blood, ileal and colonic mucosa, respectively. U represents upregulated gene expression, D denotes downregulated gene expression, and NS indicates “not statistically significant” (with the DEG threshold set as |log_2_FC| > log_2_1.5 and *P* < 0.05).
